# Nature-based allied health: current practice, challenges and opportunities

**DOI:** 10.1101/2024.05.23.24307802

**Authors:** Jessica Stanhope, Kristen Foley, Mary Butler, Jennifer Boddy, Kelly Clanchy, Emma George, Rachel Roberts, Paul Rothmore, Amy Salter, Patricia Serocki, Abirami Thirumanickam, Philip Weinstein

## Abstract

**Purpose:** To guide the effective use of nature-based approaches, we aimed to determine current practice, challenges and proposed solutions concerning the use of these approaches by allied health professionals (AHPs). We also investigated the signs, symptoms and conditions AHPs believe nature-based approaches may prevent and/or manage, as well as the perceived impact of the COVID-19 pandemic.

**Materials and methods:** Allied health professionals who used or wanted to use in nature-based approaches in Australia were invited to complete an online questionnaire. Qualitative data were analysed through inductive coding and categorisation, while descriptive statistics were reported for the quantitative data.

**Results:** Allied health professionals indicated that exposure to nature could prevent and/or manage a range of physical and mental health, social and developmental outcomes. Perceived challenges were identified, related to the patients/clients, AHPs themselves, and external factors. Recommended solutions included increasing education for AHPs and the general public, advancing more research, implementing changes to governance, and legitimisation of nature-based approaches as part of allied health practice.

**Conclusions:** Increasing awareness and evidence of the use of nature-based approaches among the AHPs and across sectors – particularly with policy, education, funding, and health management– will support their legitimacy and potential benefit a range of populations.

**Implications for rehabilitation:**

- Allied health professionals reported that nature-based approaches may help to prevent and/or manage a range of physical, mental, social and developmental health outcomes.
- Challenges to implementing nature-based approaches included patient/ client, allied health professional, and external factors.
- To address these challenges, legitimisation of nature-based approaches is key, which may be driven predominantly by research and education.

## Background

There is increasing evidence that supports the health benefits of exposure to nature [1–13], and engaging in nature-based activities [14–28]. Allied health professionals (AHPs) can play an important role in improving people’s access to and use of nature, by working with individual patients/clients, communities and from a broader public health perspective, including advocacy for and input into the design of public natural environments. Yet, few studies to date have explored AHP perceptions of the use and benefits of nature-based approaches (NBAs); nor collected data around their experiences of doing so, including the challenges encountered and changes suggested to overcome these challenges. We fill this research gap in an Australian context.

Exposure to nature is important for human health and wellbeing [29,30]. Humans have evolved as part of natural ecosystems, yet urbanisation and industrialisation limit our exposure to nature, and may be contributing to the increases in a range of adverse health outcomes, including non-communicable conditions [31]. While healthcare was often provided in natural environments in Western countries in the 19^th^ and 20^th^ centuries – e.g. asylums and rehabilitation hospitals were often located within large green spaces [32–34] – newer healthcare settings are often in highly urbanised areas or there has been urban infill of the greenspaces surrounding these locations. People who are most vulnerable (e.g. people living with disabilities, older people) often cannot access natural spaces without assistance [35].

While there has been a shift away from healthcare settings in nature, there has been an increase in research into the benefits of exposure to nature (e.g. green [1–13] and blue spaces [2,10,13]), and nature-based activities to improve health and wellbeing [14–28]. Allied health professionals are well placed to integrate NBAs – by encompassing outdoor and indoor nature-based activities which may be recommended or directly engaged in with the patient/ client, and advocacy for or the provision of improvements to natural spaces – into their practice. Allied health professionals already use models that integrate the environment (e.g. the International Classification of Functioning, Disability and Health [36]) with the importance of the natural environment discussed as early as 2012 [37]), supporting people to engage in desired or necessary activities and environments, and working within interdisciplinary teams drawing upon the expertise of others to reduce barriers to exposure. Some of these activities may include those that enhance the environment (e.g. ecosystem conservation and restoration) potentially assisting in reducing eco-despair. Emerging evidence shows NBAs may be used to prevent and manage a range of health and wellbeing outcomes that AHPs are experts in (e.g. anxiety [2,15], depression [2,15], cardiometabolic [15], cardiovascular [5] and respiratory conditions [38], pain [39] and loneliness [8]).

Although research has been conducted regarding the use NBAs amongst by AHPs [17,27,40–53], to our knowledge only three studies have examined AHPs use of these NBAs outside of a research setting [34,54,55]. Those three studies examined occupational therapists and physiotherapists engaging in nature-based rehabilitation with people with acquired brain injuries [34], social workers using NBAs with children and parents living in shelters [54], and occupational therapists using gardening or garden-related activities [55]. Other studies have investigated broader groups of health professionals in mental health [56] and the use of outdoor talk therapies (not specific to natural settings) by psychologists [57,58]. No broad study of how AHPs use NBAs has been undertaken, which limits knowledge about their experiences and views – including which signs, symptoms, and conditions AHPs believe that can be prevented and/or managed through exposure to nature – as well as challenges and potential solutions to doing so.

## Objectives

Despite the potential role of NBAs in allied health practice, we know little about how and why AHPs integrate NBAs into their practice – and what the perceived benefits of doing so are. The objectives of this study were to determine 1) what signs, symptoms, and conditions AHPs believe NBAs may prevent/manage, 2) what types of NBAs AHPs engaged with and wanted to engage with, 3) the impact of the COVID-19 pandemic on AHPs engagement with NBAs, and 4) the challenges and solutions proposed by AHPs regarding NBAs. The AHPs involved in the study were exercise physiologists, occupational therapists, physiotherapists, psychologists, social workers, and speech pathologists registered or certified to work in Australia. The broader project also sought to determine what features AHPs think are important in outdoor natural environments; detail which is available elsewhere [59].

## Methods

The study had approval from The University of Adelaide Human Research Ethics Committee (HREC-2021-174). This study was conducted in two cross-sectional stages due to challenges with recruitment in Stage 1 leading to a reframing of the study (Stage 2).

## Stage 1 recruitment

This study was conducted in two cross-sectional stages due to challenges with recruitment in Stage 1 leading to study reframing (Stage 2). In addition to the aims outlined above, our original intention was also to understand the proportion of AHPs in Australia who were engaging in NBAs, and what factors were associated with such engagement (Stage 1). To be eligible for inclusion, AHPs had to be registered or certified to practice within Australia. To avoid sampling bias, Stage 1 was initially advertised as relating to contemporary approaches to allied health, rather than specifically nature-based allied health. Recruitment was conducted via advertisements (e.g. newsletter, social media, website) through Occupational Therapy Australia and Speech Pathology Australia, and was supplemented via advertisements on the investigators social media pages (e.g. LinkedIn, Twitter). After recruiting occupational therapists and speech pathologists (October 2021-January 2022), it became apparent that the sample size would be too low to determine the proportion of AHPs using NBAs and the factors associated with such engagement; hence these objectives were removed from the study. Consequently, questions were removed that were no longer relevant, and some additional questions were added (e.g. the age group the AHP predominantly works with). The project was relaunched following amendment approval from the HREC, and was advertised as relating to targeting those who currently use or want to use nature-based approaches. The data from Stage 1 was used where appropriate, as outlined below.

## Stage 2 recruitment

Stage 2 recruitment was conducted in May-December 2022. Recruitment was conducted through the same procedure as Stage 1, however we advertised to all six allied health disciplines, through their professional organisations (Exercise and Sports Science Australia, Occupational Therapy Australia, Australian Physiotherapy Association, Australian Psychological Society, Australian Association of Social Workers, and Speech Pathology Australia), and supplemented with advertisements on the social media pages of the investigators. Allied health professionals were eligible to participate if they were registered/ certified to practice in Australia, and were engaging with or interested in engaging with nature-based allied health. Respondents were asked if they had participated in the Stage 1 study and sent to an exit page if they indicated yes. Demographic data were cross-checked between respondents in Stages 1 and 2 to confirm whether any AHP participated in both stages – with no duplication detected.

## Data collection

Interested AHPs were invited to follow a web link to the online questionnaire (via Survey Monkey^TM^) used for data collection (Supplementary Material 1). In short, we asked AHPs for demographic information, the signs, symptoms or conditions they felt could be prevented and/or managed by spending time in outdoor natural environments and through indoor nature-based exposures (open response), the activities in outdoor natural environments and indoor nature-based exposures they had either recommended to patients/ clients or directly engaged in with patients/ clients in the last 12 months (from a list of activities, with an open response ‘other’ option), the spaces they recommended or directly engaged in for outdoor NBAs (from a list of spaces, with an open response ‘other’ option), the outdoor and indoor NBAs they would like to have engaged in, any services they referred patients/ clients to assist in engaging with NBAs, any advocacy or provision of additional natural spaces or improvements to these spaces in the last 12 months in their professional or personal lives (from a list of natural spaces), the impact COVID-19 had on their engagement in each type of activity (e.g. indoor nature-based exposure recommendations, by indicating increase, decrease or no change), and the challenges encountered in engaging with each type of activity (e.g. recommending indoor nature-based exposures) and solutions they would like to see implemented (open response). Examples provided of outdoor natural environments were parks, gardens, farms, lakes or the ocean, while the examples of indoor nature-based exposures were nature pictures, sounds, scents, virtual reality and driving through natural environments.

## Data analysis

Qualitative and quantitative approaches were used to analyse the data. Where appropriate, percentages of respondents were also reported. Owing to differences in the recruitment and inclusion criteria for the two stages of the project, all percentages reported are from to Stage 2, except where specifically indicated, as the Stage 1 respondents were not necessarily using or want to use NBAs. No discipline-specific percentages are reported, due to the small sample sizes from each discipline (n=1-22). Nonetheless, the discipline-specific percentages are reported in the Supplementary Material 2 for interested readers, as are results related to the type of practice the AHPs engaged in (e.g. aged care, mental health, rehabilitation).

Qualitative analysis was conducted through a general inductive approach [60], where categories were developed from the qualitative data with no *a priori* expectations [60]. Insights from this inductive analysis were discussed by the two data analysts, to refine potential interpretations and how it might augment or confer/contrast with findings from the quantitative data analysis. This process followed the logic of retroductive inference [61], borrowing from a critical realist approach where data and analysis from epistemologically distinct approaches can be mediated and synthesised to develop more robust understandings of a particular phenomenon [62].

The analyses were primarily conducted by an occupational therapist (KF – qualitative) and a physiotherapist (JS – quantitative). Input was sought from all co-authors (across the six allied health disciplines and public health), to refine the results and interpretations arising from the data collected.

## Results

There were 77 respondents across the two stages. In Stage 1, there were 16 respondents who were occupational therapists (n=4) or speech pathologists (n=12) who were not necessarily using or wanting to use NBAs, but responded to at least one of the survey items about nature-based therapies (Table 1 for demographics). A further 61 respondents were included from Stage 2 (22 psychologists, 15 social workers, 10 physiotherapists, nine occupational therapists, four speech pathologists, and one exercise physiologist). Stage 2 respondents were predominantly women (85%), with 38% aged 20-39 years (Table 1).

**Table 1:**
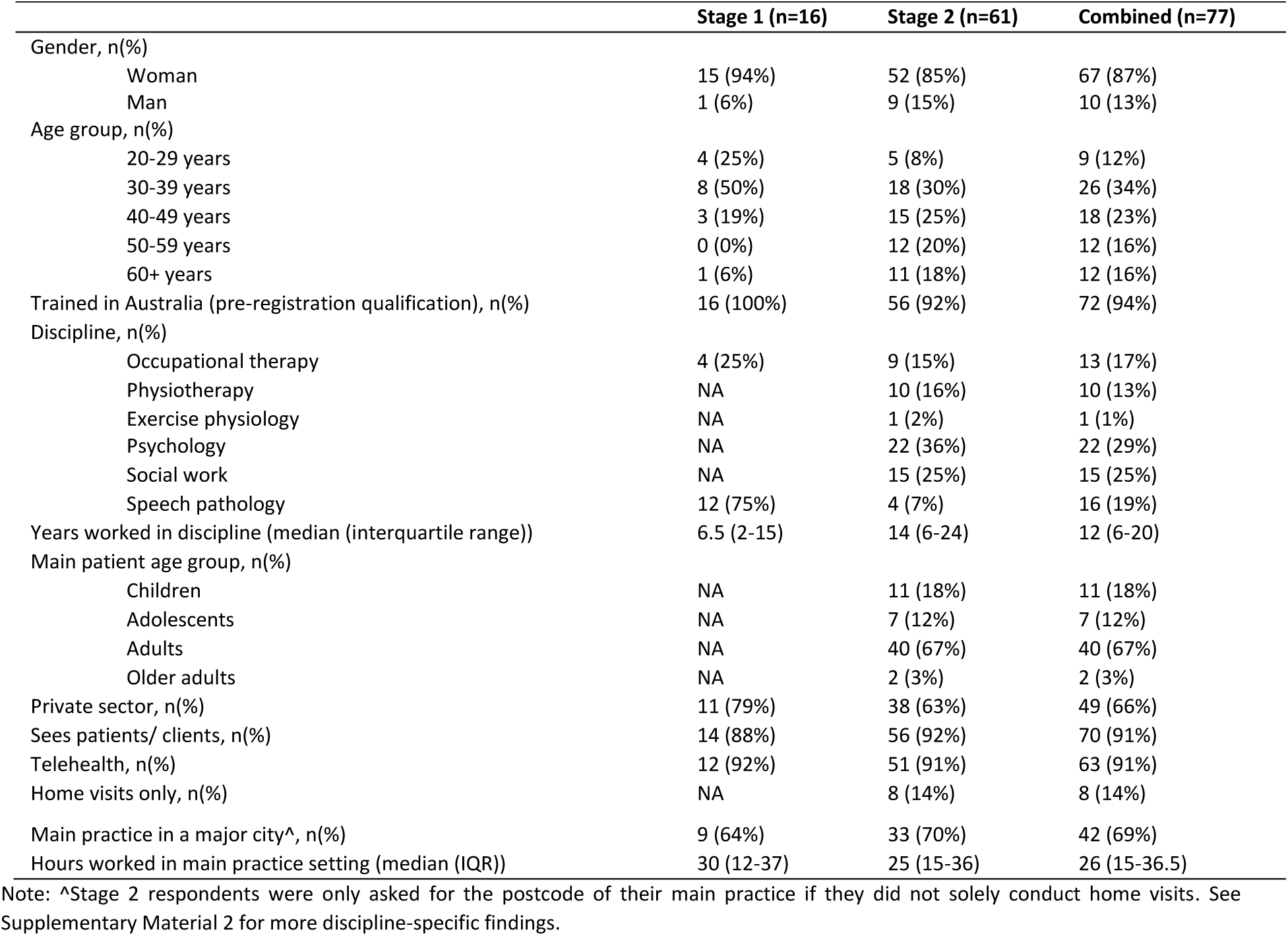
Respondent demographics.

The majority (91%) of respondents worked with patients/ clients, as opposed to those who worked in other allied health roles (e.g. public health). Of the 86% who worked with patients/ clients and did not solely conduct home visits, the majority of AHP had grass/lawn (75%), garden (75%) and trees (81%) in the 50m around their main practice setting, while 29% had a waterbody within the same radius.

### The signs, symptoms and conditions that may be prevented and/or managed through exposure to nature

Almost all AHPs respondents from Stage 2 identified people could benefit from spending time in outdoor natural environments (97%) and having indoor nature-based exposures (94%). Respondents reported a broad range of signs, symptoms, and conditions which they felt could be improved with NBAs in various contexts (e.g. rehabilitation) and settings (e.g. aged care) were further reported (see Table 2; Stages 1-2). The survey responses regarding signs, symptoms and conditions were classified as physical and mental health, and social and developmental outcomes, with a general category for broader statements such as chronic conditions and “most presentations”.

**Table 2:**
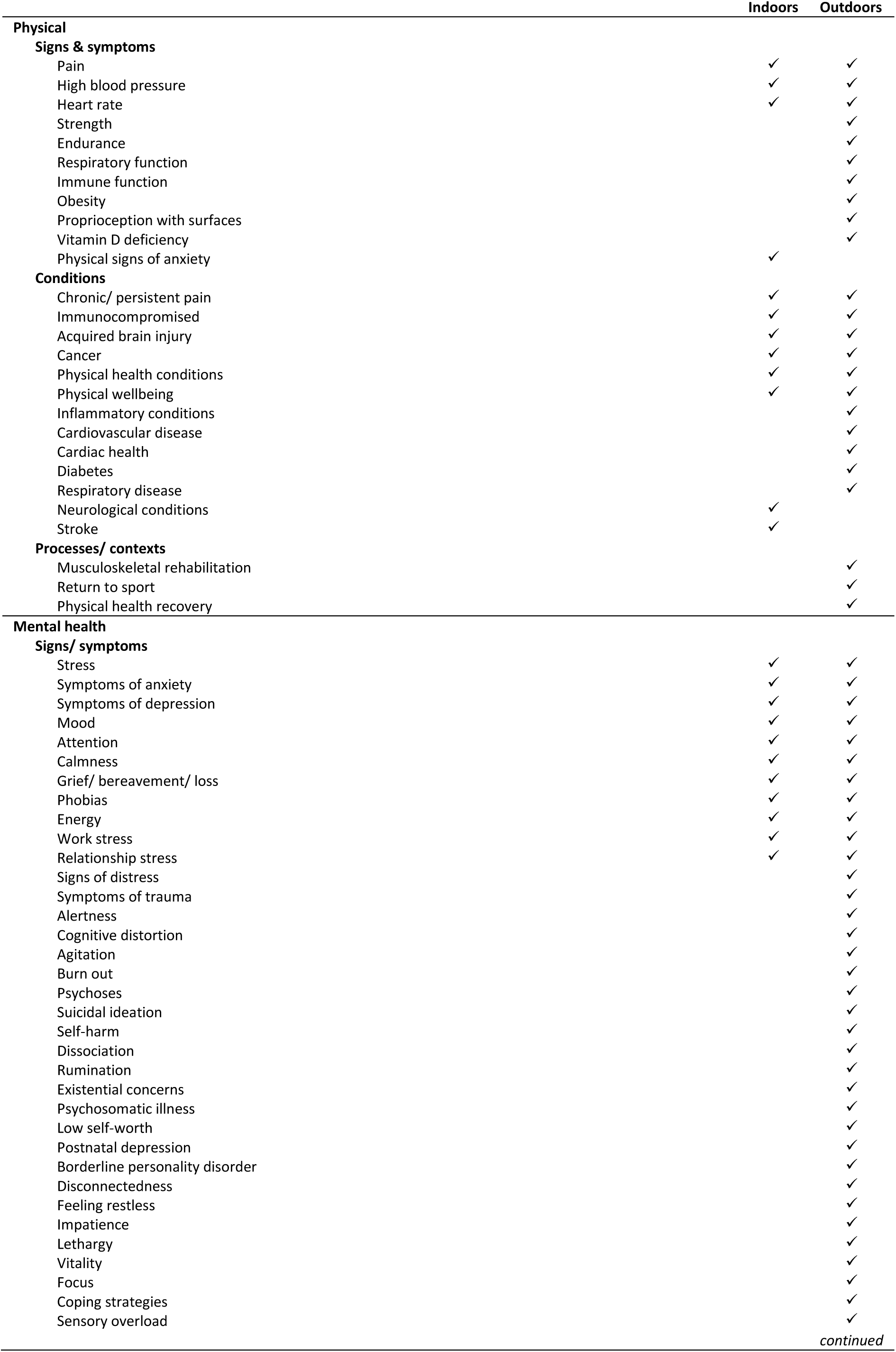
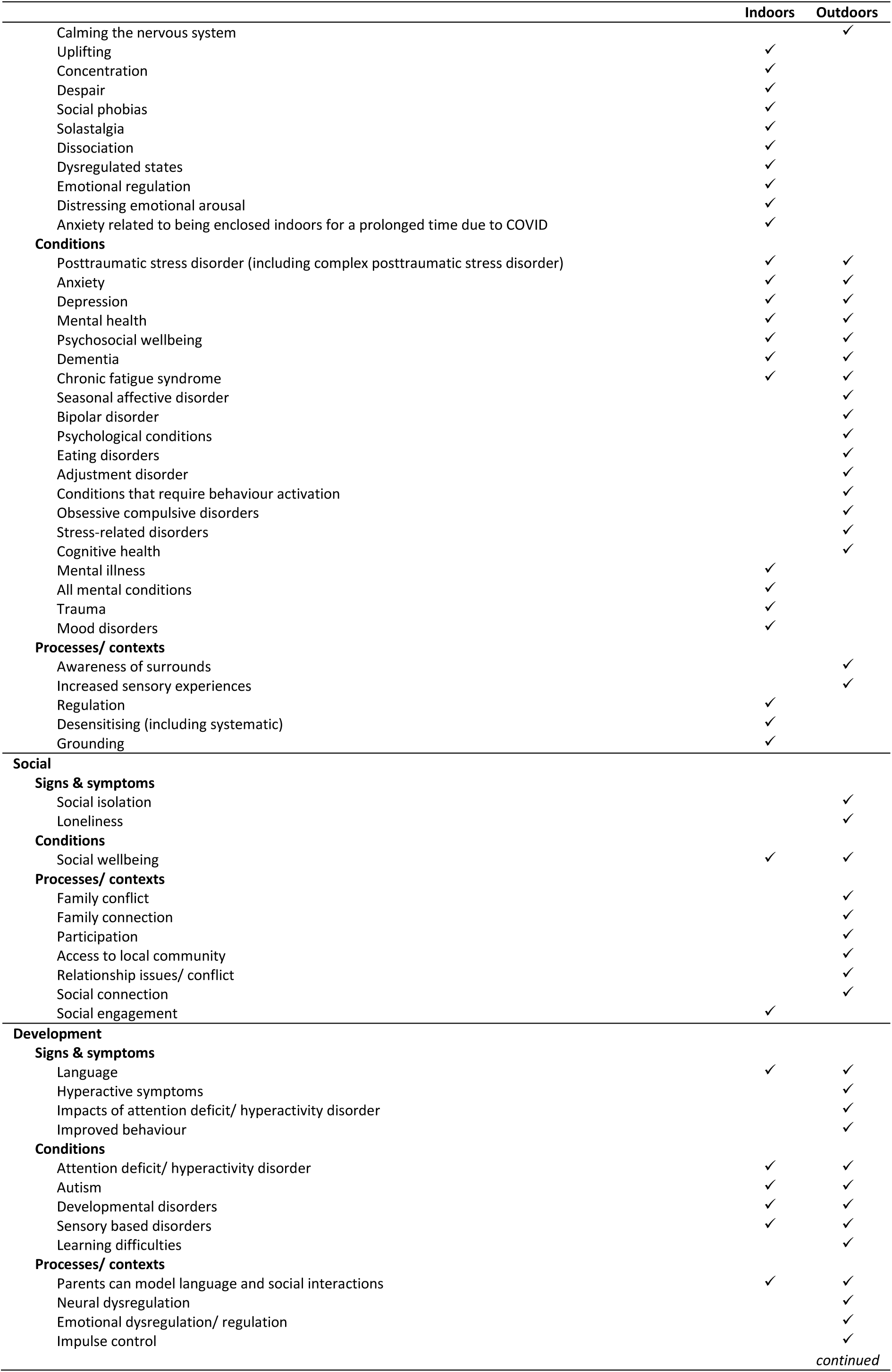
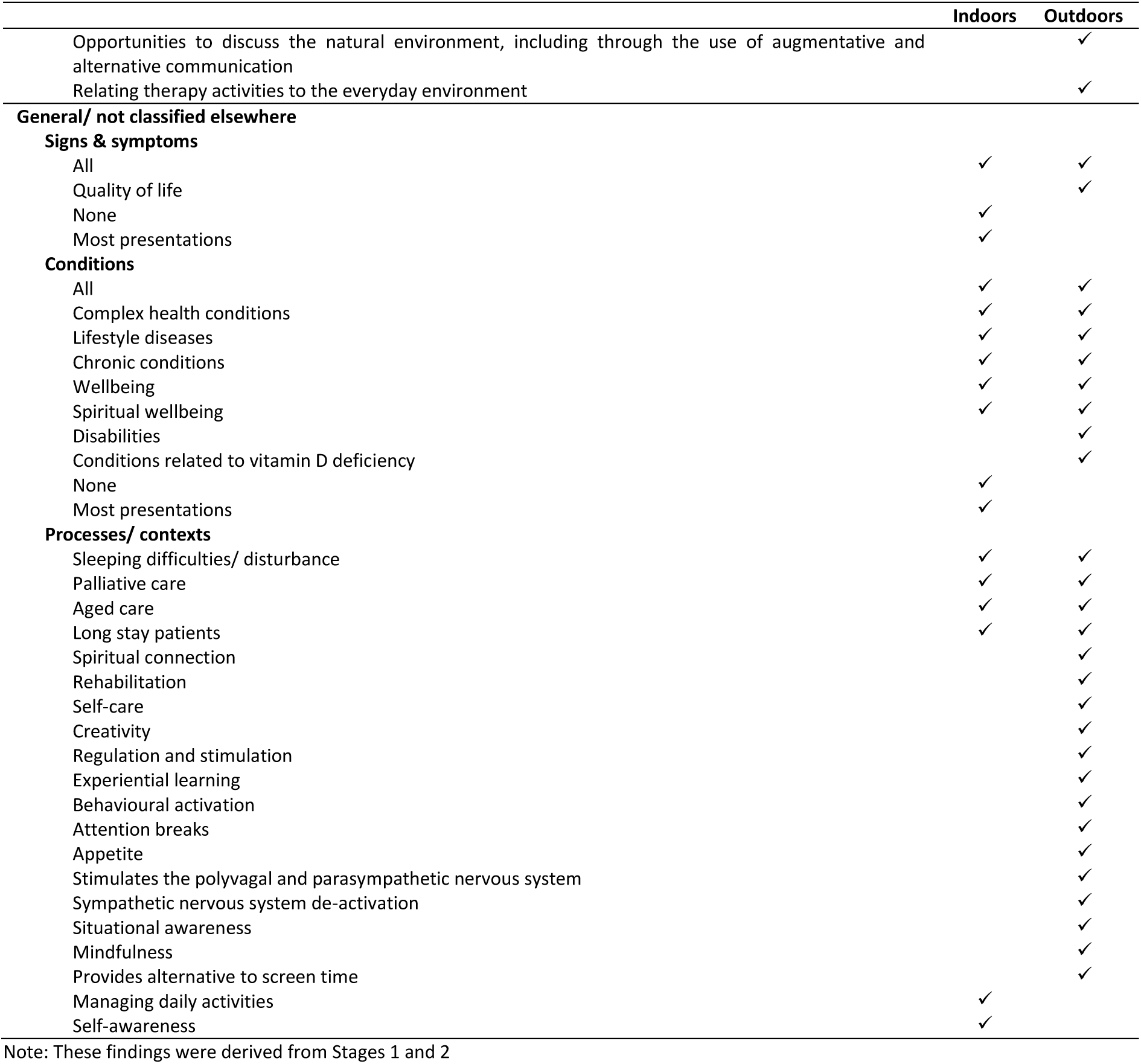
The signs, symptoms and conditions allied health professionals report may be improved with nature-based activities.

|  | Indoors | Outdoors |
| --- | --- | --- |
| **Physical** |  |  |
| **Signs & symptoms** |  |  |
| Pain | ✓ | ✓ |
| High blood pressure | ✓ | ✓ |
| Heart rate | ✓ | ✓ |
| Strength |  | ✓ |
| Endurance |  | ✓ |
| Respiratory function |  | ✓ |
| Immune function |  | ✓ |
| Obesity |  | ✓ |
| Proprioception with surfaces |  | ✓ |
| Vitamin D deficiency |  | ✓ |
| Physical signs of anxiety | ✓ |  |
| **Conditions** |  |  |
| Chronic/ persistent pain | ✓ | ✓ |
| Immunocompromised | ✓ | ✓ |
| Acquired brain injury | ✓ | ✓ |
| Cancer | ✓ | ✓ |
| Physical health conditions | ✓ | ✓ |
| Physical wellbeing | ✓ | ✓ |
| Inflammatory conditions |  | ✓ |
| Cardiovascular disease |  | ✓ |
| Cardiac health |  | ✓ |
| Diabetes |  | ✓ |
| Respiratory disease |  | ✓ |
| Neurological conditions | ✓ |  |
| Stroke | ✓ |  |
| **Processes/ contexts** |  |  |
| Musculoskeletal rehabilitation |  | ✓ |
| Return to sport |  | ✓ |
| Physical health recovery |  | ✓ |
| **Mental health** |  |  |
| **Signs/ symptoms** |  |  |
| Stress | ✓ | ✓ |
| Symptoms of anxiety | ✓ | ✓ |
| Symptoms of depression | ✓ | ✓ |
| Mood | ✓ | ✓ |
| Attention | ✓ | ✓ |
| Calmness | ✓ | ✓ |
| Grief/ bereavement/ loss | ✓ | ✓ |
| Phobias | ✓ | ✓ |
| Energy | ✓ | ✓ |
| Work stress | ✓ | ✓ |
| Relationship stress | ✓ | ✓ |
| Signs of distress |  | ✓ |
| Symptoms of trauma |  | ✓ |
| Alertness |  | ✓ |
| Cognitive distortion |  | ✓ |
| Agitation |  | ✓ |
| Burn out |  | ✓ |
| Psychoses |  | ✓ |
| Suicidal ideation |  | ✓ |
| Self-harm |  | ✓ |
| Dissociation |  | ✓ |
| Rumination |  | ✓ |
| Existential concerns |  | ✓ |
| Psychosomatic illness |  | ✓ |
| Low self-worth |  | ✓ |
| Postnatal depression |  | ✓ |
| Borderline personality disorder |  | ✓ |
| Disconnectedness |  | ✓ |
| Feeling restless |  | ✓ |
| Impatience |  | ✓ |
| Lethargy |  | ✓ |
| Vitality |  | ✓ |
| Focus |  | ✓ |
| Coping strategies |  | ✓ |
| Sensory overload |  | ✓ |
| *continued* |  |  |
| Calming the nervous system |  | ✓ |
| Uplifting | ✓ |  |
| Concentration | ✓ |  |
| Despair | ✓ |  |
| Social phobias | ✓ |  |
| Solastalgia | ✓ |  |
| Dissociation | ✓ |  |
| Dysregulated states | ✓ |  |
| Emotional regulation | ✓ |  |
| Distressing emotional arousal | ✓ |  |
| Anxiety related to being enclosed indoors for a prolonged time due to COVID | ✓ |  |
| **Conditions** |  |  |
| Posttraumatic stress disorder (including complex posttraumatic stress disorder) | ✓ | ✓ |
| Anxiety | ✓ | ✓ |
| Depression | ✓ | ✓ |
| Mental health | ✓ | ✓ |
| Psychosocial wellbeing | ✓ | ✓ |
| Dementia | ✓ | ✓ |
| Chronic fatigue syndrome | ✓ | ✓ |
| Seasonal affective disorder |  | ✓ |
| Bipolar disorder |  | ✓ |
| Psychological conditions |  | ✓ |
| Eating disorders |  | ✓ |
| Adjustment disorder |  | ✓ |
| Conditions that require behaviour activation |  | ✓ |
| Obsessive compulsive disorders |  | ✓ |
| Stress-related disorders |  | ✓ |
| Cognitive health |  | ✓ |
| Mental illness | ✓ |  |
| All mental conditions | ✓ |  |
| Trauma | ✓ |  |
| Mood disorders | ✓ |  |
| **Processes/ contexts** |  |  |
| Awareness of surrounds |  | ✓ |
| Increased sensory experiences |  | ✓ |
| Regulation | ✓ |  |
| Desensitising (including systematic) | ✓ |  |
| Grounding | ✓ |  |
| **Social** |  |  |
| **Signs & symptoms** |  |  |
| Social isolation |  | ✓ |
| Loneliness |  | ✓ |
| **Conditions** |  |  |
| Social wellbeing | ✓ | ✓ |
| **Processes/ contexts** |  |  |
| Family conflict |  | ✓ |
| Family connection |  | ✓ |
| Participation |  | ✓ |
| Access to local community |  | ✓ |
| Relationship issues/ conflict |  | ✓ |
| Social connection |  | ✓ |
| Social engagement | ✓ |  |
| **Development** |  |  |
| **Signs & symptoms** |  |  |
| Language | ✓ | ✓ |
| Hyperactive symptoms |  | ✓ |
| Impacts of attention deficit/ hyperactivity disorder |  | ✓ |
| Improved behaviour |  | ✓ |
| **Conditions** |  |  |
| Attention deficit/ hyperactivity disorder | ✓ | ✓ |
| Autism | ✓ | ✓ |
| Developmental disorders | ✓ | ✓ |
| Sensory based disorders | ✓ | ✓ |
| Learning difficulties |  | ✓ |
| **Processes/ contexts** |  |  |
| Parents can model language and social interactions | ✓ | ✓ |
| Neural dysregulation |  | ✓ |
| Emotional dysregulation/ regulation |  | ✓ |
| Impulse control |  | ✓ |
| Opportunities to discuss the natural environment, including through the use of augmentative and alternative communication |  | ✓ |
| Relating therapy activities to the everyday environment |  | ✓ |
| **General/ not classified elsewhere** |  |  |
| **Signs & symptoms** |  |  |
| All | ✓ | ✓ |
| Quality of life |  | ✓ |
| None | ✓ |  |
| Most presentations | ✓ |  |
| **Conditions** |  |  |
| All | ✓ | ✓ |
| Complex health conditions | ✓ | ✓ |
| Lifestyle diseases | ✓ | ✓ |
| Chronic conditions | ✓ | ✓ |
| Wellbeing | ✓ | ✓ |
| Spiritual wellbeing | ✓ | ✓ |
| Disabilities |  | ✓ |
| Conditions related to vitamin D deficiency |  | ✓ |
| None | ✓ |  |
| Most presentations | ✓ |  |
| **Processes/ contexts** |  |  |
| Sleeping difficulties/ disturbance | ✓ | ✓ |
| Palliative care | ✓ | ✓ |
| Aged care | ✓ | ✓ |
| Long stay patients | ✓ | ✓ |
| Spiritual connection |  | ✓ |
| Rehabilitation |  | ✓ |
| Self-care |  | ✓ |
| Creativity |  | ✓ |
| Regulation and stimulation |  | ✓ |
| Experiential learning |  | ✓ |
| Behavioural activation |  | ✓ |
| Attention breaks |  | ✓ |
| Appetite |  | ✓ |
| Stimulates the polyvagal and parasympathetic nervous system |  | ✓ |
| Sympathetic nervous system de-activation |  | ✓ |
| Situational awareness |  | ✓ |
| Mindfulness |  | ✓ |
| Provides alternative to screen time |  | ✓ |
| Managing daily activities | ✓ |  |
| Self-awareness | ✓ |  |

Note: These findings were derived from Stages 1 and 2

Several respondents reported there would be greater benefits related to outdoor nature exposures, compared with indoors. One respondent reported no expectation of benefits from indoor exposures, while another noted that indoors was safer and could still provide some of the nature exposure. Notable differences in the signs, symptoms and conditions reported for both indoor and outdoor NBAs are shown within Table 2 – suggesting that AHPs perceive outdoor NBAs to have a greater effect.

### The nature-based activities allied health professionals are engaging with

Overall, 94% of respondents engaging with patients/clients reported having recommended outdoor NBAs in the 12 months prior to data collection, with most respondents reporting land-based gross motor activities, gardening, engaging with animals, mindfulness/ meditation/ relaxation, and social activities in outdoor natural spaces. Most respondents (83%) reported that they had recommended time in outdoor natural spaces with no specific activity recommended (Table 3). In considering the responses for ‘other’ outdoor NBAs, respondents from Stages 1-2 also reported they had directly engaged with patients/clients in reading, art, community cultural development and speech/ language/ communication therapy. The majority of respondents (Stage 2) reported recommending these activities be performed in private gardens (75%), public parks (75%), spaces near water (65%), and in national parks (52%; Table 3).

**Table 3:** Nature-based activities used and recommended by allied health professionals.

|  |  | Recommendations by allied health professionals (% Stage 2 only) | Direct engagement by allied health professionals (% Stage 2 only) | Physiotherapists/ exercise physiologist | Occupational therapists | Speech pathologists | Social workers | Psychologists |
| --- | --- | --- | --- | --- | --- | --- | --- | --- |
| **Outdoor nature-based activities** | Any | 94 | 82 | ^# | ^# | ^# | ^# | ^# |
|  | Land-based gross motor activities | 60 | 33 | ^# | ^# | ^# | ^# | ^# |
|  | Land-based fine motor activities | 44 | 24 | ^# | ^# |  | ^# | ^# |
|  | Water-based physical activities | 46 | 14 | ^# | ^# | ^ | ^ | ^# |
|  | Active transport | 31 | 8 | ^# | ^# | ^ | ^# | ^ |
|  | Powered transport | 15 | 8 | ^# | ^# |  | ^ | ^# |
|  | Gardening | 67 | 20 | ^# | ^# | ^ | ^# | ^# |
|  | Ecological conservation or restoration | 23 | 4 | ^ | ^# |  | ^# | ^# |
|  | Engaging with/ tending to animals | 54 | 14 | ^# | ^# | ^# | ^# | ^# |
|  | Wilderness activities | 33 | 10 | ^# | ^# | ^# | ^ | ^# |
|  | Nature play | 42 | 18 | ^# | ^# | ^# | ^# | ^# |
|  | Nature observation activities | 50 | 24 | ^# | ^# | ^# | ^# | ^# |
|  | Mindfulness/ meditation/ relaxation | 83 | 55 | ^# | ^# | ^# | ^# | ^# |
|  | Forest bathing | 21 | 8 | ^ | # |  | ^ | ^# |
|  | Social activities | 56 | 29 | ^# | ^# | ^# | ^# | ^# |
|  | No specific activity | 83 | 53 | ^# | ^# | ^# | ^# | ^# |
|  | Other activities | 4 | 2 |  | # | # | # | ^# |
| **Outdoor nature-based activity locations** | Private garden | 75 | 33 | ^# | ^# | ^# | ^# | ^# |
|  | Community/ school garden | 40 | 27 | ^# | ^# | # | ^# | ^# |
|  | Farm | 25 | 6 | ^# | ^ | ^ | ^ | ^# |
|  | Public park | 75 | 53 | ^# | ^# | ^# | ^# | ^# |
|  | National park | 52 | 12 | ^ | ^# | ^# | ^ | ^# |
|  | Forest | 40 | 12 | ^ | ^# | ^# | ^ | ^# |
|  | Oval/ sports field | 25 | 24 | ^# | ^# |  | ^# | ^# |
|  | On the water (lake, river or ocean) | 38 | 6 | ^ | ^# |  | ^ | ^# |
|  | Near water (lake, river or ocean) | 65 | 31 | ^# | ^# | ^# | ^# | ^# |
|  | Other | 0 | 2 |  |  |  |  | # |
| **Indoor nature-based activities** | Any | 75 | 74 | ^# | ^# | ^# | ^# | ^# |
|  | Listening to natural sounds | 44 | 38 | ^# | ^# | # | ^# | ^# |
|  | Having indoor pot plants | 42 | 55 | ^# | ^ | # | ^# | ^# |
|  | Pictures of natural scenes | 25 | 38 | ^# | # | ^# | ^# | ^# |
|  | Natural scents | 35 | 28 | ^# | ^ |  | ^# | ^# |
|  | Travelling in a car or on public transport through natural environments | 25 | 9 |  | ^# |  | ^# | ^# |
|  | Views of nature outside the window | 42 | 43 | ^# | ^# | ^# | ^# | ^# |
|  | Open windows for fresh air and the smells and sounds of nature | 42 | 34 | ^# | ^# | ^# | ^# | ^# |
|  | Nature-based virtual reality | 6 | 11 | ^# | ^# |  | # | ^# |
|  | Other | 13 | 11 | ^# | ^# |  | ^# | ^# |

^ (blue) indicates recommendation, # (orange) indicates direct engagement, ^# (blue) indicates both recommendation and direct engagement by at least one respondent (either Stage 1 or 2). See Supplementary Material 2 for more discipline-specific findings.

Respondents were also asked whether they had referred patients/ clients to other services (including other health professionals) in enacting their recommendations for spending time in outdoor natural spaces, with 40% respondents indicating that they had. The services reported (from Stages 1 and 2) were equine therapy and “equine mentoring”, community gardens, walking groups, sporting groups, support groups, and events, nature play, nature craft, beautification projects, tai chi, meditation, neighbourhood centres, the council, neighbourhood houses, activities in the park program, workplaces, occupational therapy, other allied health professionals, personal trainers, general practitioners, mentors, and support workers. This elucidates the broad scope of NBAs utilised by AHPs.

Direct engagement in outdoor nature-based activities with patients/ clients was less common than recommendations (82%). The most reported activity was mindfulness/ meditation/ relaxation (55%), while no specific activity was also frequent (53%). Again, the dominant spaces were public parks (53%), private gardens (33%), and spaces near water (31%). Two respondents (one each from Stages 1 and 2) reported additional activities they had directly engaged in with patients/ clients: wildlife cruises and conducting speech/ language/ communication therapy in parks. One respondent (Stage 2) reported using a carpark with garden supplies (presumably to engage in gardening activities), which could not be classified into the existing categories.

For indoor NBAs, the percentage of respondents reporting recommendations (75%) was similar to that for direct engagement (74%). The most commonly reported activities were nature sounds (44% recommended, 38% directly engaged), indoor plants (42% recommended, 55% directly engaged), views of nature (42% recommended, 43% directly engaged), and opening windows (42% recommended, 34% directly engaged). Other activities recommended by respondents (Stage 1 n=1, Stage 2 n=6) that did not fit into the existing categories were videos/ documentaries of nature, sharing photos, discussions about nature, using natural items, sand trays, cut flowers, sharing pot plants with the local café, “sleep stories”, and visiting animals, public libraries, museums, galleries and archives. Respondents (Stage 1 n=1, Stage 2 n=5) reported directly engaging with respondents in watching nature videos/ documentaries, using salt lamps, cut flowers, discussions about nature, metaphors related to nature, guided imagery, visiting animals, using natural items, making art, gardening, community cultural development, and visiting public libraries, archives, galleries, and museums.

Overall, 73% of respondents (Stage 2) were involved in advocacy for the provision or improvement to natural environments; most commonly care gardens (37%), nature elements in non-care environments (35%), and community gardens (33%) (Table 4), with others also mentioning national/coastal parks and street trees. These advocacy activities included those in both their professional and personal lives. We did not ask specifically which improvements were advocated for (rather than which spaces) however one respondent reported table tennis and another bush regeneration (Stage 2). A respondent from Stage 1 also reported advocating for their organisation to move to a building with outdoor spaces, and another reported blogging about the therapeutic aspects of such spaces as well as the installation of disability playgrounds as a form of advocacy for who should be able to access, use and benefit from public spaces.

**Table 4:**
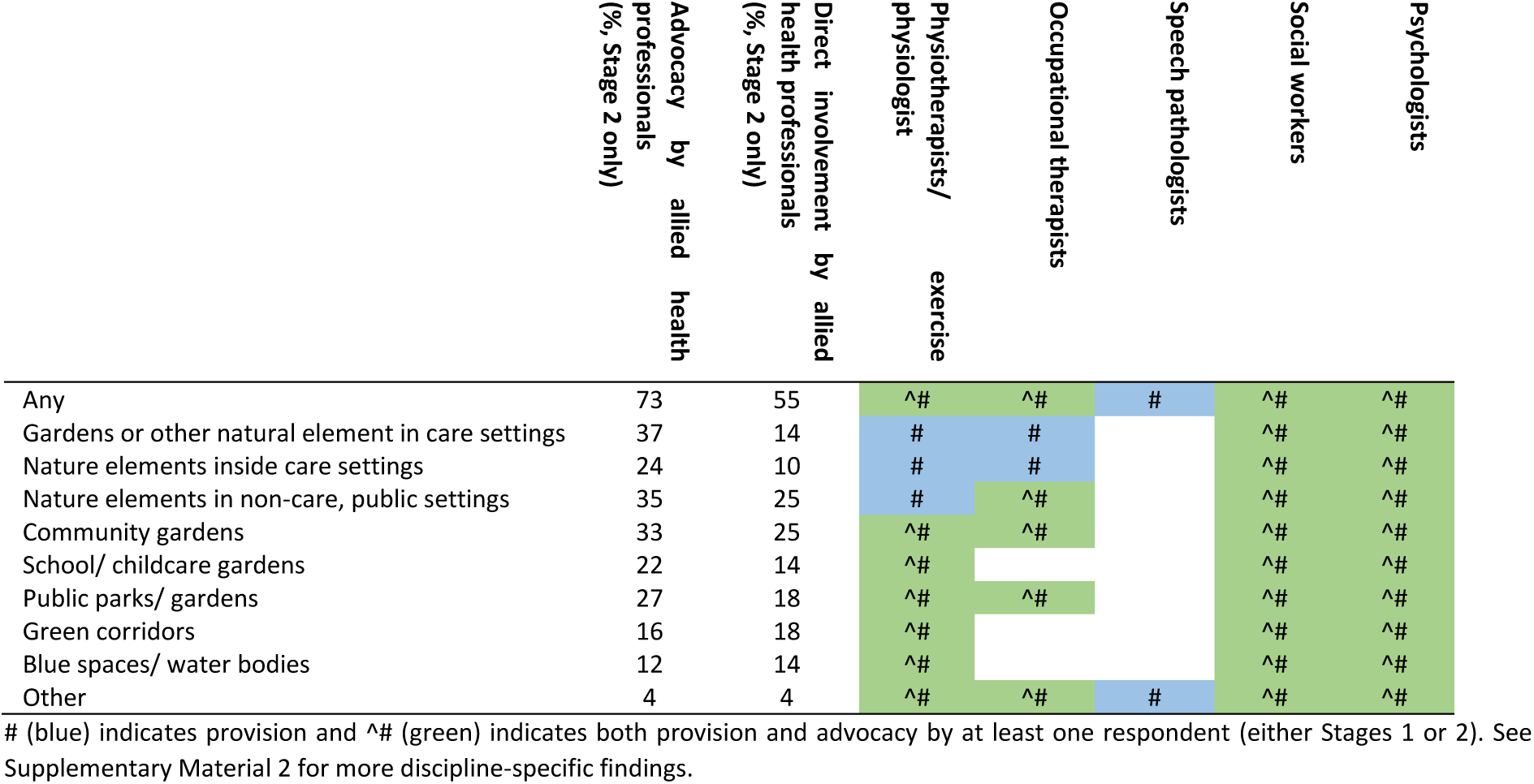
Outdoor natural environments where allied health professionals advocated for or were directly involved in the provision of improvements in their professional and/or personal lives.

|  | Advocacy by allied health professionals (% Stage 2 only) | Direct involvement by allied health professionals (% Stage 2 only) | Physiotherapists/physiologist | Occupational therapists | Speech pathologists | Social workers | Psychologists |
| --- | --- | --- | --- | --- | --- | --- | --- |
| Any | 73 | 55 | ^# | ^# | # | ^# | ^# |
| Gardens or other natural element in care settings | 37 | 14 | # | # |  | ^# | ^# |
| Nature elements inside care settings | 24 | 10 | # | # |  | ^# | ^# |
| Nature elements in non-care, public settings | 35 | 25 | # | ^# |  | ^# | ^# |
| Community gardens | 33 | 25 | ^# | ^# |  | ^# | ^# |
| School/ childcare gardens | 22 | 14 | ^# |  |  | ^# | ^# |
| Public parks/ gardens | 27 | 18 | ^# | ^# |  | ^# | ^# |
| Green corridors | 16 | 18 | ^# |  |  | ^# | ^# |
| Blue spaces/ water bodies | 12 | 14 | ^# |  |  | ^# | ^# |
| Other | 4 | 4 | ^# | ^# | # | ^# | ^# |

Just over half of respondents (55%) were also engaged directly in the provision of natural, public spaces, with community gardens and nature elements in non-care environments the most commonly reported (25% for both), in their personal or professional lives (Table 4). One respondent (Stage 2) reported consulting on an Aboriginal outdoor space in a hospital, and another involved in an urban tree canopy campaign. Respondents reported direct engagement with collecting rubbish, land care and planting activities (Stage 2), while another reported assisting others in engaging with the community garden by providing transport (Stage 1).

### The impact of COVID-19 pandemic

Decreased engagement with NBAs was reported during the pandemic, particularly direct engagement in outdoor NBAs (18%). However, overall there was a net increase in recommendations for outdoor nature-based approaches (see Figure 1). One respondent reported how “Covid-19 has highlighted the success of flexible working from home arrangements, reducing workers [sic] commute/travel time and allowing people to spend more time in outdoor natural environment [sic]. However, in recent months there has been push/pressure from organisations to get workers back into the workplace setting, reducing workers [sic] ability to spend time in natural environment [sic] to support work-life balance” indicating that there may have been an increase in engagement earlier in the COVID-19 pandemic, which has since waned.

**Figure 1:**
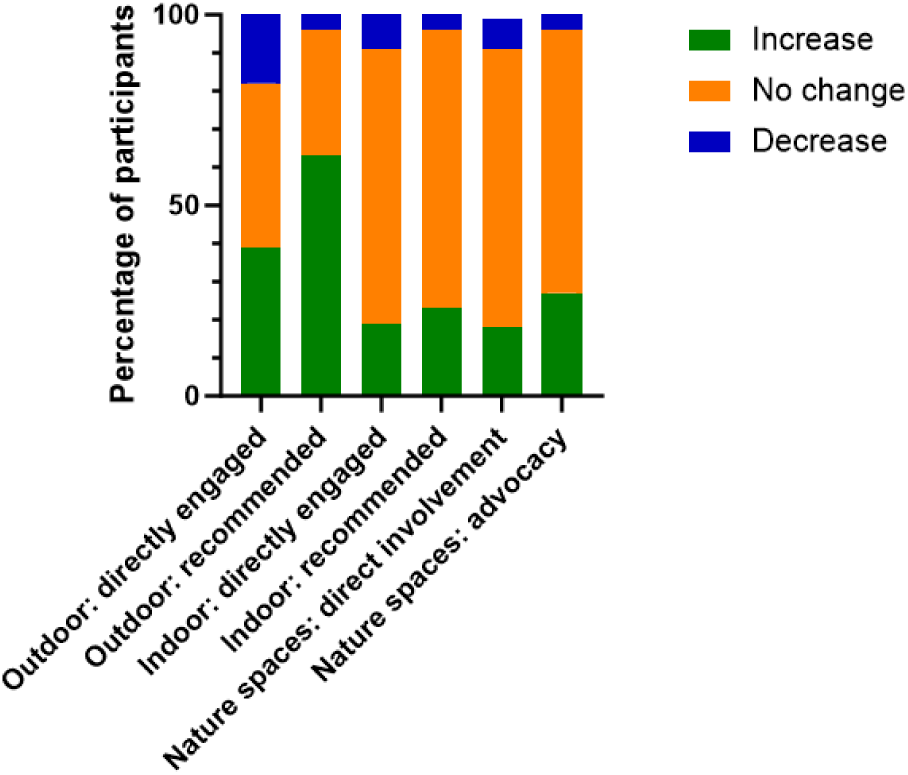
Respondent-reported change in engagement in nature-based approaches due to COVID-19. Note: Findings were taken from Stage 2 only.

### The nature-based approaches allied health professionals would like to have engaged in but did not

Several respondents reported NBAs they would have liked to engage with in the 12 months prior, with respect to recommendations for both outdoor (14%) and indoor (15%) activities, and directly engaging in these activities with patients/ clients (36% and 21% for outdoors and indoors, respectively), with specific activities reported in Table 5. Respondents also reported wanting to do more, particularly with regards to nature play, walking, and time doing sessions outdoors. There appeared to be a drive to do more, with one respondent indicating that this was a “new era”, and they would like to learn more about what they could be doing, while another stated that they would “like to have a specified work role associated with nature-based therapy”, indicating the value they place on these forms of therapy.

**Table 5:** Nature-based activities allied health professionals would have liked to engage in in the 12 months prior but did not.

|  | Outdoor nature-based activities | Indoor nature-based activities |
| --- | --- | --- |
| **Recommendation** | - Park Run<br>- Hiking<br>- Sports<br>- Fitness classes<br>- Activities that incorporate wellness<br>- Land care activities<br>- Volunteer work with animals<br>- Youth-based adventure camps (including on Country)<br>- Travel | - Nature posters in homes<br>- Nature sounds in homes<br>- Gardening based activities<br>- Animal assisted therapy<br>- Virtual reality<br>- Social media activities (e.g. following nature restoration activities)<br>- “All of the above” (likely indicating all the activities listed in the questionnaire) |
| **Direct engagement** | - Physical activity<br>- Walking (including walk and talk therapy)<br>- Sitting down for therapy<br>- Using playgrounds<br>- Gardening<br>- Hippotherapy<br>- Kayaking<br>- Bouldering<br>- Camping<br>- Forest bathing<br>- Meditation<br>- Mindfulness<br>- Beach activities<br>- Forest activities<br>- Park activities<br>- Structured activities<br>- Assisted patients/ clients in visiting local outdoor spaces and facilitated access | - Nature-based activities in homes and clinics<br>- Nature sounds in homes<br>- Nature smells in homes<br>- Opening windows<br>- Pot plants<br>- Mindfully attending to different textures of plants<br>- Gardening based activities<br>- Travelling through natural environments<br>- Virtual reality<br>- Social media activities (e.g. following nature restoration activities)<br>- Zoom events for nature-based activities |

Note: in the advocacy question related to what they would have liked to have engaged with but did not also reported they would have liked to have had clients visit a local seed back, and to have an office garden club for clients. Findings were taken from Stages 1 and 2.

Several respondents also reported activities related to advocating for improvements to outdoor natural environments (31%) and for engaging in the provision of such improvements (24%; Table 6). These included advocacy for and the provision of nature spaces (e.g. therapeutic gardens, community gardens), and enhancements to spaces (e.g. increased biodiversity, native planting, mindfulness and meditation spaces, disability playgrounds, transport to the community garden, therapeutic landscapes). Respondents made general statements like “lots”, “any of the above” and “all of them”. In addition to the results listed in Table 6, respondents reported the desire to have been involved more in decision-making regarding the improvement to or provision of natural environments. Two respondents also reported advocacy activities they would have liked to have engaged with to increase the use of outdoor nature spaces; one reported a desire to ban electric scooters on university campuses to increase active transport, the other a wish to offer outdoor classes at university. Finally, one respondent reported blogging regarding the benefits of exposure to nature.

**Table 6:** Advocacy and provision for improvements to outdoor natural environments that allied health professionals would have liked to engage in in the 12 months prior but did not.

|  | Advocacy | Provision of improvements |
| --- | --- | --- |
| **Types of spaces** | - Greater variety of parks (including in childcare, school and public settings)<br>- Therapeutic garden at their practice<br>- School/ childcare gardens<br>- Community gardens<br>- More natural spaces | - School/ childcare gardens<br>- Gardens and therapeutic horticulture<br>- Community gardens (including those specific to children, and on campus)<br>- Community gardens specifically for children |
| **Features in spaces** | - Raised garden beds in community gardens<br>- Soundscapes<br>- Increased biodiversity in the local park<br>- Native plantings that bring birds, native bees and insects<br>- Outdoor exercise equipment on the university campus<br>- More places for people to stop and sit<br>- Seats near trees<br>- More group activities<br>- More natural trails around the city<br>- Accessibility to outdoor environments<br>- Footpath restoration<br>- Creative play materials<br>- Natural sculptures | - Development and design of outdoor workout and movement spaces<br>- Designing outdoor mindfulness and meditation spaces<br>- Nature elements insight care settings<br>- Modifying local recreation grounds to encourage children to engage with nature not just sport<br>- "Wild space conservation work" |

Note: Findings were taken from Stages 1 and 2

### Challenges related to nature-based activities

The challenges related NBAs were broadly categorised as challenges associated with the patients/ clients, with the AHPs, and external factors (e.g. weather, governance; Table 7). Patient/ client factors related to time, motivation, understanding the value to their health and wellbeing, concerns about judgement from others, COVID, safety and privacy, and patient/ client capacity. The challenges relating to the AHP included time, funding, expectations of others (e.g. does it align with AHP roles), confidence, and concerns regarding risk management and ethics. Finally, external factors included a lack of appropriate space, funding, administrative barriers, logistics and the weather.

**Table 7:**
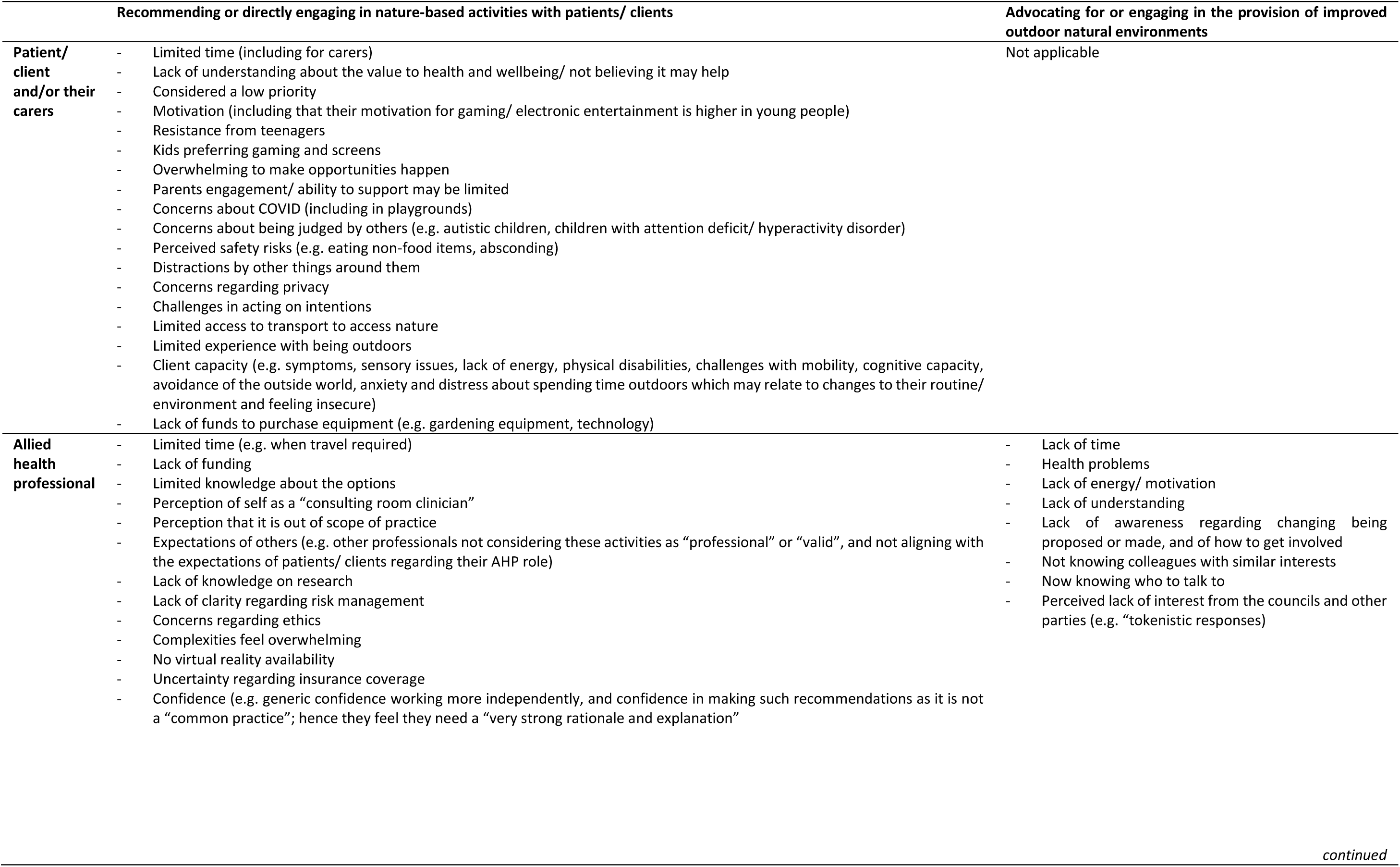
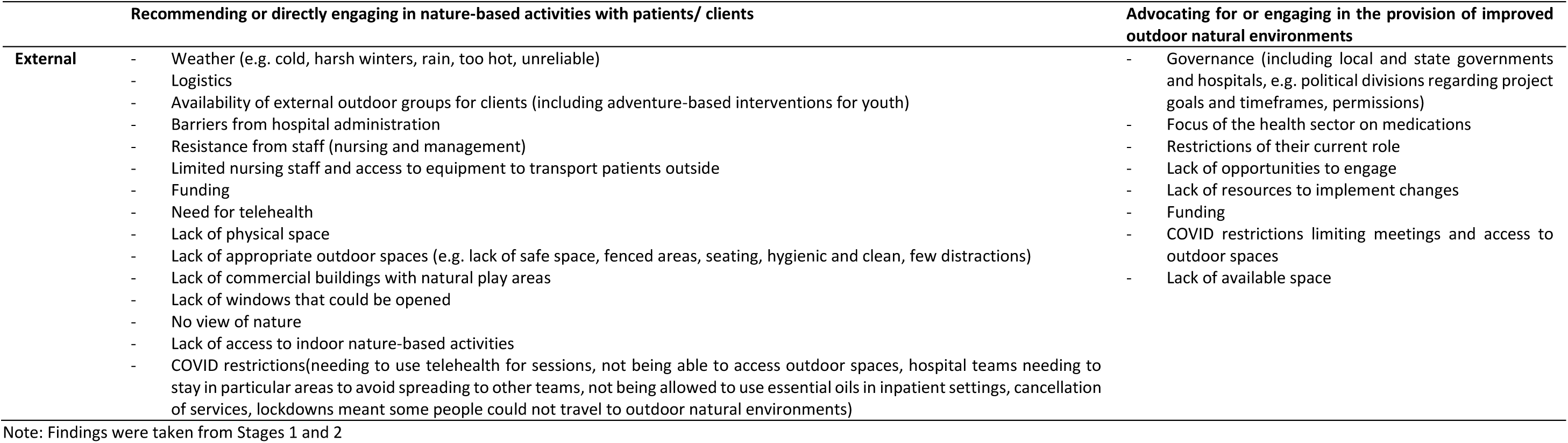
Challenges reported by allied health professionals for recommending or directly engaging in nature-based activities with patients/ clients.

Concerns regarding NBA as a scope of practice within AHP were exemplified by one of the respondents from Stage 1 who, in response to being asked about the challenges related to recommending indoor nature-based exposures stated “This is not a speech pathologist’s responsibility nor area of professional practice. Your questioning is misguided. The challenge in doing it would be any SP [speech pathologist] would then be over-stepping the mark”, and went on to write (in response to the question about solutions to the challenge) “Get back within your professional boundaries as a SP, and refresh your ethical responsibilities”. Despite this strong opinion, the respondent recommended to patients/ clients that they should be opening windows and having views of nature from indoors, as well as engaging directly in “speech/ language/ communication therapy in parks”. The same respondent stated that both indoor and outdoor nature exposures could benefit general health and wellbeing.

In addition to the responses outlined in Table 7, several respondents indicated elsewhere in the questionnaire other challenges related to NBAs. The need for a patient-centred approach was indicated by one respondent who stated recommendations for NBAs depend “on the client’s individual needs and preferences and capacity”, while another respondent indicated that clinicians may have different views regarding the use of NBAs. The latter stated that when working with social work students, they are “sometimes restrained in recommending [the] value of nature. It’s my lens but I need to let others explore/ develop their own worldviews”. Other challenges reported elsewhere in the questionnaire related to advocacy and provision of improvements to natural spaces, including: such advocacy being of low priority traditionally and not emphasised in training; local community gardens having a wait list; and finding out about relevant community events on social media following the fact.

### Proposed solutions to challenges of engaging with nature-based activities

Solutions to the abovementioned challenges proposed by the AHPs, broadly related to the research evidence, education, governance, support from professional bodies, access to nature-based activities, access to nature and COVID-19 restrictions, and the need for legitimisation of NBAs as part of AHP practice (Table 8). Some respondents did not propose solutions, with one highlighting the complexity of integrating NBAs, while maintaining a patient-centred approach, and stating “I want to be able to meet people where they are at and I don’t want to impact engagement by having them feel like they are being forced into an environment that makes them uncomfortable”. The statement indicates that a NBA approach might not always be appropriate.

**Table 8:**
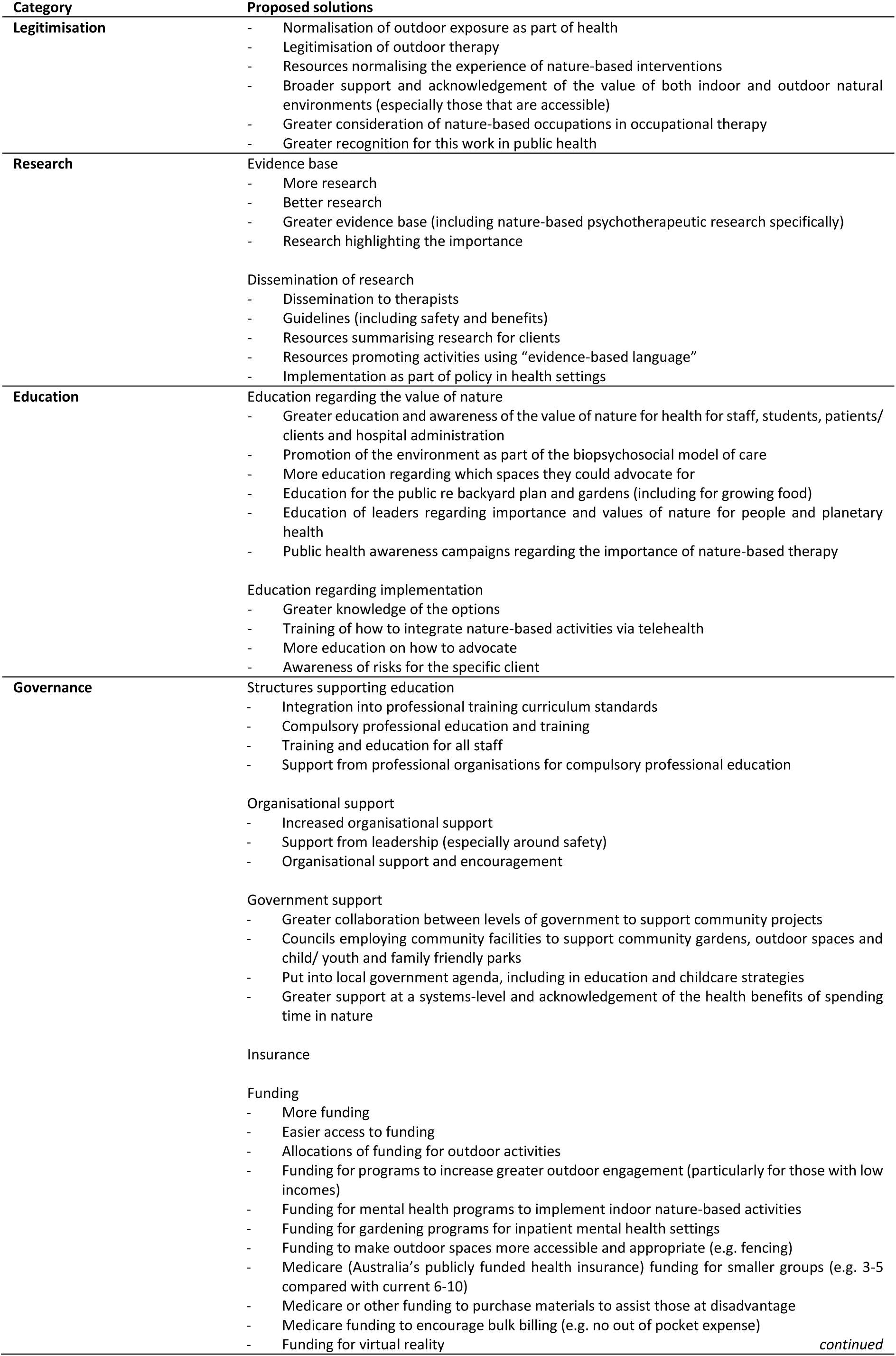
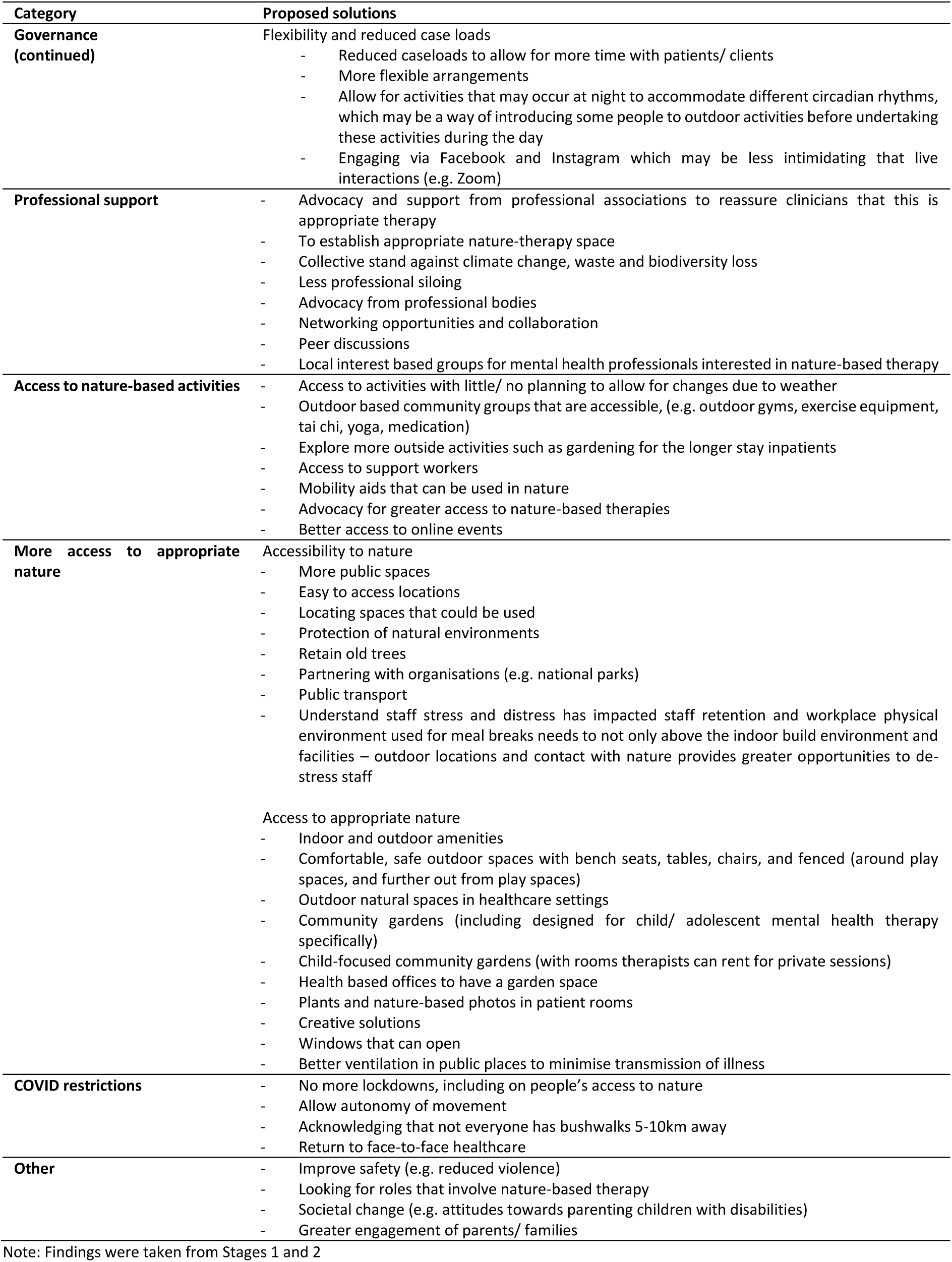
Suggested solutions combined, for recommendations, direct engagement, and advocacy and provision.

| Category | Proposed solutions |
| --- | --- |
| **Legitimation** | - Normalisation of outdoor exposure as part of health<br>- Legitimation of outdoor therapy<br>- Resources normalising the experience of nature-based interventions<br>- Broader support and acknowledgement of the value of both indoor and outdoor natural environments (especially those that are accessible)<br>- Greater consideration of nature-based occupations in occupational therapy<br>- Greater recognition for this work in public health |
| **Research** | Evidence base<br>- More research<br>- Better research<br>- Greater evidence base (including nature-based psychotherapeutic research specifically)<br>- Research highlighting the importance<br>Dissemination of research<br>- Dissemination to therapists<br>- Guidelines (including safety and benefits)<br>- Resources summarising research for clients<br>- Resources promoting activities using “evidence-based language”<br>- Implementation as part of policy in health settings |
| **Education** | Education regarding the value of nature<br>- Greater education and awareness of the value of nature for health for staff, students, patients/clients and hospital administration<br>- Promotion of the environment as part of the biopsychosocial model of care<br>- More education regarding which spaces they could advocate for<br>- Education for the public re backyard plan and gardens (including for growing food)<br>- Education of leaders regarding importance and values of nature for people and planetary health<br>- Public health awareness campaigns regarding the importance of nature-based therapy<br>Education regarding implementation<br>- Greater knowledge of the options<br>- Training of how to integrate nature-based activities via telehealth<br>- More education on how to advocate<br>- Awareness of risks for the specific client |
| **Governance** | Structures supporting education<br>- Integration into professional training curriculum standards<br>- Compulsory professional education and training<br>- Training and education for all staff<br>- Support from professional organisations for compulsory professional education<br>Organisational support<br>- Increased organisational support<br>- Support from leadership (especially around safety)<br>- Organisational support and encouragement<br>Government support<br>- Greater collaboration between levels of government to support community projects<br>- Councils employing community facilities to support community gardens, outdoor spaces and child/ youth and family friendly parks<br>- Put into local government agenda, including in education and childcare strategies<br>- Greater support at a systems-level and acknowledgement of the health benefits of spending time in nature<br>Insurance<br>Funding<br>- More funding<br>- Easier access to funding<br>- Allocations of funding for outdoor activities<br>- Funding for programs to increase greater outdoor engagement (particularly for those with low incomes)<br>- Funding for mental health programs to implement indoor nature-based activities<br>- Funding for gardening programs for inpatient mental health settings<br>- Funding to make outdoor spaces more accessible and appropriate (e.g. fencing)<br>- Medicare (Australia’s publicly funded health insurance) funding for smaller groups (e.g. 3-5 compared with current 6-10)<br>- Medicare or other funding to purchase materials to assist those at disadvantage<br>- Medicare funding to encourage bulk billing (e.g. no out of pocket expense)<br>- Funding for virtual reality |
| **Governance (continued)** | Flexibility and reduced case loads<br>- Reduced caseloads to allow for more time with patients/ clients<br>- More flexible arrangements<br>- Allow for activities that may occur at night to accommodate different circadian rhythms, which may be a way of introducing some people to outdoor activities before undertaking these activities during the day<br>- Engaging via Facebook and Instagram which may be less intimidating than live interactions (e.g. Zoom) |
| **Professional support** | - Advocacy and support from professional associations to reassure clinicians that this is appropriate therapy<br>- To establish appropriate nature-therapy space<br>- Collective stand against climate change, waste and biodiversity loss<br>- Less professional siloing<br>- Advocacy from professional bodies<br>- Networking opportunities and collaboration<br>- Peer discussions<br>- Local interest based groups for mental health professionals interested in nature-based therapy |
| **Access to nature-based activities** | - Access to activities with little/ no planning to allow for changes due to weather<br>- Outdoor based community groups that are accessible, (e.g. outdoor gyms, exercise equipment, tai chi, yoga, medication)<br>- Explore more outside activities such as gardening for the longer stay inpatients<br>- Access to support workers<br>- Mobility aids that can be used in nature<br>- Advocacy for greater access to nature-based therapies<br>- Better access to online events |
| **More access to appropriate nature** | Accessibility to nature<br>- More public spaces<br>- Easy to access locations<br>- Locating spaces that could be used<br>- Protection of natural environments<br>- Retain old trees<br>- Partnering with organisations (e.g. national parks)<br>- Public transport<br>- Understand staff stress and distress has impacted staff retention and workplace physical environment used for meal breaks needs to not only be above the indoor built environment and facilities – outdoor locations and contact with nature provides greater opportunities to de-stress staff<br>Access to appropriate nature<br>- Indoor and outdoor amenities<br>- Comfortable, safe outdoor spaces with bench seats, tables, chairs, and fenced (around play spaces, and further out from play spaces)<br>- Outdoor natural spaces in healthcare settings<br>- Community gardens (including designed for child/ adolescent mental health therapy specifically)<br>- Child-focused community gardens (with rooms therapists can rent for private sessions)<br>- Health based offices to have a garden space<br>- Plants and nature-based photos in patient rooms<br>- Creative solutions<br>- Windows that can open<br>- Better ventilation in public places to minimise transmission of illness |
| **COVID restrictions** | - No more lockdowns, including on people's access to nature<br>- Allow autonomy of movement<br>- Acknowledging that not everyone has bushwalks 5-10km away<br>- Return to face-to-face healthcare |
| **Other** | - Improve safety (e.g. reduced violence)<br>- Looking for roles that involve nature-based therapy<br>- Societal change (e.g. attitudes towards parenting children with disabilities)<br>- Greater engagement of parents/ families |

Note: Findings were taken from Stages 1 and 2

Awareness of possible approaches appeared important. It was reported in response to other questions that completing the survey had given the AHPs additional ideas about what they would now like to engage with, and another stating that they had not really thought about the benefit of recommending indoor nature-based activities. Additionally, one respondent indicated they will add information regarding the value of nature to their wellbeing kits. Another respondent stated they have lots of ideas for “future” work, including visiting botanic gardens, a small wildlife sanctuary, pet friendly venues and overnight stays, however it is unclear whether these were also activities they had wanted to engage with in the previous 12 months.

## Discussion

Our study is the first to explore the perspectives of AHPs on how nature exposure and NBAs can address various health concerns, alongside the challenges and proposed solutions in implementing NBAs. Crucially, the findings substantiate the legitimacy of NBAs within allied health practices, demonstrating that AHPs not only employ a diverse array of NBAs but also recognize their efficacy in managing and enhancing physical and mental, and social and developmental outcomes. This substantiation of NBAs by AHPs underscores the necessity for further research to deepen the evidence base regarding NBAs’ effectiveness. Such research could reinforce the established legitimacy of NBAs, addressing ancillary issues like funding and accessibility. Our study thus marks a significant step towards integrating NBAs into standard allied health practices, backed by AHPs’ firsthand experiences and the positive health outcomes observed through NBA interventions.

### Benefits of exposure to nature

Allied health professionals highlighted the important role that exposure to nature may prevent and/or manage a range of physical and mental health, and social and developmental outcomes, with mental health outcomes dominating the responses. The emphasis on mental health, as reflected by our findings, underscores AHPs recognition of nature’s benefits to humans. This focus aligns with a substantial body of evidence supporting nature exposure [1–3] and NBAs [14–16,19–24,26,27] in the prevention and/or management of mental health concerns. Our sample comprised predominantly of psychologists (29%), social workers (25%) and occupational therapists (17%) who often work in mental health settings; hence, the bias in our sample may have led to the dominance of mental health outcomes. Although research on physical health [5,11,13–15,18,20,23,38,63,64], social [8,23] and developmental outcomes [7,10] is still emerging, it shows promising growth.

Our study also revealed a preference among AHPs for outdoor nature exposures over indoor alternatives. These greater benefits may relate to the diverse and sensory-rich experiences associated with outdoor nature environments. Outdoor settings have been described previously as offering a unique, “invigorating” experience that can enhance health outcomes [65], contrasting with the more controlled and less varied indoor settings. While indoor nature exposure can serve as an accessible introduction to nature’s benefits, especially for vulnerable groups with limited access to outdoor nature [35], the dynamic and immersive outdoor environment may hold the greatest value for health.

### Nature-based activities used by allied health professionals

Our findings reveal that AHPs are involved in a broad spectrum of both outdoor and indoor NBAs, encompassing traditional clinical activities adapted to outdoor natural settings (e.g. motor activities, mindfulness), initiatives promoting direct nature engagement (e.g. ecological conservation), and modifications to indoor environments for nature exposure (e.g. indoor plants, nature sounds). There are multiple mechanisms through which exposure to nature may influence health outcomes, including exposure to environmental microbiota, biogenic volatile organic compounds and negative air ions, the sights and sounds of nature and sunlight exposure [39]. However, the integration of allied health approaches with nature exposures provides additional opportunities. For example, the use of nature provides a naturalistic milieu to promote speech development using incidental teaching [66], and building functional skills and confidence in real-world settings, including outdoor environments may assist in overcoming disparities in exposure to nature, where those who are more vulnerable likely have less access to nature [35]. Some of these activities could be integrated into prevocational training, with for example, prevocational farm activities [67] and horticulture programs [68], improving health and wellbeing, and a potential role for environmental enhancement (e.g. ecological restoration) activities having been recently highlighted [69]. The diverse engagement in NBAs not only showcases the versatility of NBAs, but also underscores their recognised legitimacy within the scope of AHP practice.

A large percentage of our respondents engaged in advocacy (73%) and the provision (55%) of or improvements to outdoor natural spaces, reflecting an inherent understanding of public health’s integral role within AHP responsibilities. This advocacy, whether professional or personal, is a new finding and illustrates the commitment of AHPs to integrate NBAs more fully into health practices.

Engaging in environmental enhancement activities, like gardening, ecological conservation and restoration, offers AHPs a unique opportunity to adopt a relational approach to the environment; one that transcends conventional AHP training and emphasises healing interactions with nature. This relational approach is deeply rooted in Indigenous practice, where the focus in not merely on the utility of nature, but on fostering a reciprocal relationship that nurtures both human and ecosystem health. Timler and Sandy’s [70] work with Indigenous communities in Canada highlights this perspective, revealing how gardening activities can shift from a focus on harvesting to a deeper engagement with caring for plants, understanding the intricate dynamics of ecosystems, and advocating for the preservation of nature for future generations’ health and wellbeing. Similarly, in Australia, such engagements offer pathways to (re)connect with the land, or ‘Country’, and embrace Indigenous insights into living in harmony with nature, thereby enriching AHP’s practices with a holistic and healing approach to environmental interactions. Through such a lens, the environment is seen not just as a setting but as an integral part of health and wellbeing, where caring for the land reciprocally supports human health, and nurtures social, cultural and spiritual connections [71].

Activities related to environmental enhancement (e.g. gardening, ecological conservation and restoration) as well as animal care, which are typically beyond the realm of AHP education, may provide an opportunity for AHPs to learn alongside and potentially from their patients/ clients. This learning may also relate to the sharing of Indigenous knowledge and approaches to environmental enhancement. For example, Timler and Sandy [70] explored working with Indigenous people in Canada in gardening activities, where different perspectives were elucidated, around shifting the focus away from harvesting towards caring for the plant, understanding the complexity of the environment (e.g. seasonality, weather, plants and animals), and the need to protect nature for the health and wellbeing of future generations. In Australia, such activities may include opportunities to reconnect with Country and related Indigenous knowledge. Engaging in NBAs or facilitating such engagement may allow people of all backgrounds to reconnect with places and activities of significance to themselves, their families and cultures, but AHPs need to be mindful of understanding and integrating different perspectives.

### Impact of COVID-19

During the COVID-19 pandemic, our respondents largely increased their engagement with NBAs, including heightened advocacy for and enhancements to natural outdoor spaces. This trend aligns with findings from other studies, indicating a rise in outdoor therapeutic practices, including psychologists, conducting more outdoor talk therapy sessions [57], and a general increase in time spent by Australian in green and blue spaces [72]. The pandemic-induced restrictions likely played a role in this shift, compelling AHPs to explore alternative, outdoor settings for therapy and activities to navigate the constraints posed by lockdown and indoor gathering restrictions. Additionally, the increased indoor confinement may have heightened the collective awareness of nature’s intrinsic value and its impact on health, potentially influencing AHPs decisions to incorporate more NBAs into their practice. Our findings suggest that the pandemic may have catalysed an expansion on the way in which AHPs utilise NBAs, recognising their potential as fundamental components of health promotion and care.

### Recommendations to overcome challenges in using nature-based activities

Regarding the use of NBAs, we identified that challenges pertaining to the AHP themselves, the patient/ client, and external factors (e.g. related to the patient/ client and/or their family, organisational governance, insurance). We argue there is a need to establish a framework for legitimising NBAs, which entails building research evidence and providing education (including entry level and professional development for AHPs, as well the general public). This framework should garner support from professional bodies, AHP networks, funders, insurers and employers, thereby mitigating the obstacles to integrating NBAs into allied health practice (Figure 2).

**Figure 2:**
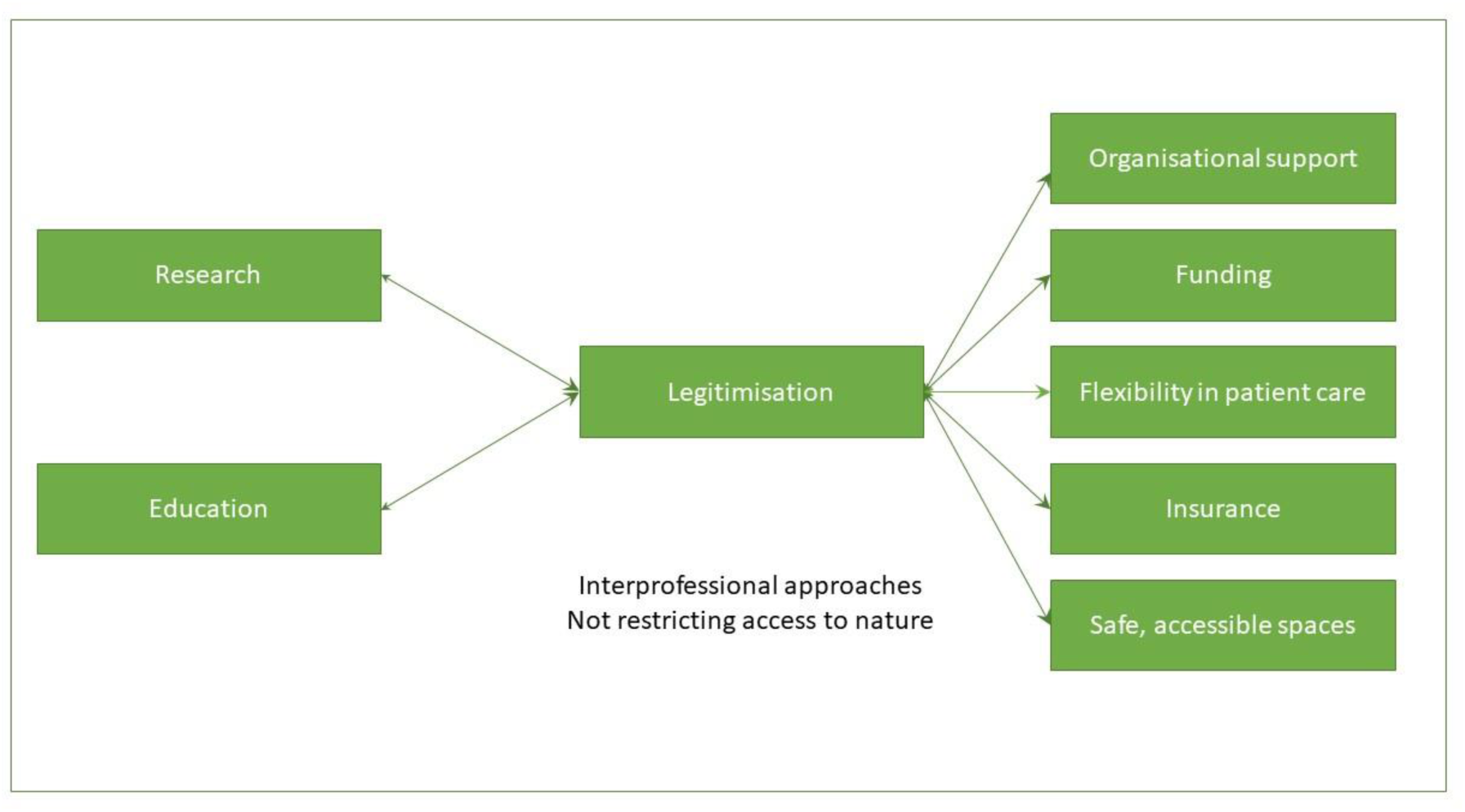
C**o**nceptual **framework for overcoming the challenges of using nature-based approaches** The connections are bidirectional, with the left-to-right direction being dominant

To effectively integrate NBAs into allied health practice, a key challenge pertains to translating research into practical, evidence-grounded strategies for AHPs. The rich body of existing research [1–28] highlights the health and wellbeing benefits of engaging with nature. However, the gap between this knowledge and its application in day-to-date practice remains. Bridging this gap requires targeted translational research that not only reaffirms the value of NBAs but also provides APHs with clear, actionable guidance on incorporating these practices in a manner that is rooted in evidence.

Education of health professionals and the general public was also highlighted as a solution to overcoming some of the current challenges. Strategies to increase the accessibility of evidence and to education clinicians and the general public may include the use of non-traditional dissemination with links to the full publications (e.g. blog posts), having collections of research evidence, providing professional development opportunities for integrating NBAs into rehabilitation practice, and support for educators to integrate this content into their teaching. These strategies are starting to be integrated in allied health, for example through the Environmental Physiotherapy Association [73,74], as well as through university programs [74,75].

Furthermore, addressing the accessibility of natural spaces – both in terms of physical accessibility and the perception of these spaces as suitable therapeutic settings – is imperative. As highlighted in previous research [57,58], suggested enhancements include facilitating easy access, promoting public transport options, establishing community gardens, providing seating and incorporating fenced areas [59]. Given the anticipated challenges posed by climate change [76]; it is crucial to design these spaces to remain cool and shaded. Improving the accessibility of natural spaces involves not only infrastructural improvements and the provision of appropriate mobility aids, but also to normalise the use of natural spaces for therapeutic purposes. By making nature-based approaches commonplace for AHPs and their patients/ clients, we can ensure that these spaces are readily accessible and beneficial to all.

Ensuring natural spaces are welcoming and accessible is crucial in extending an open invitation for NBAs. To truly integrate NBAs into allied health practice, environments must be adapted to be inclusive of all individuals, including those with disabilities and older adults. This adaptation involves not just physical modifications for accessibility, but also a broader cultural shift towards embracing nature as a therapeutic ally. Key to this transformation is the design of natural spaces with features such as easy access paths, ample and appropriate seating, sheltered areas, fencing, and clear signage, ensuring they cater to diverse needs [59]. Additionally, the provision of ‘grab and go’ therapeutic kits and adaptive equipment like the TrailRider (a mobility device designed for adaptive hiking) [77] can enable AHPs to conduct sessions seamlessly in these settings, regardless of weather conditions.

Beyond physical adaptations, fostering a societal mindset that values and prioritises accessibility in nature is essential. This mindset involves educational initiatives for the public, and support for AHPs in articulating the benefits of NBAs, thereby enhancing motivation for outdoor activities over screen time. Through such comprehensive strategies, we can create an environment that not only supports, but actively encourages NBAs for everyone.

Lastly, the broader societal perspective on NBAs and technology use needs to evolve to accommodate and value NBAs. By advocating for a more balanced approach to technology use and nature engagement, we can create an environment that supports and validates the use of NBAs in allied health practice. Indeed, targeted technology use can be used to encourage people to engage with nature [78,79]. In essence, the journey toward fully integrating NBAs into allied health practice is not just about accumulating more evidence, but about translating the existing evidence into practical, tangible practices that AHPs can confidently adopt, knowing their work is consistent with evidence-based practice.

### Recommendations for future research

Future research should prioritise the co-design of NBAs with AHPs and their patients/ clients, collaboration based on the insights provided in our study, including the proposed solutions to overcome the challenges experienced by AHPs. This collaborative approach could serve as a cornerstone for developing robust, evidence-based practices that resonate with both AHPs and those they work with. Additionally, filling the current evidence gaps regarding the impact of exposure to diverse natural environments and elements on health outcomes is essential. These gaps include exploring the effects of various types of nature (e.g. different levels of biodiversity), natural elements (e.g. environmental microbiota, natural sounds), and specific NBAs (e.g. gardening, ecological conservation and restoration) on different demographic groups, and health outcomes. Understanding the mechanisms behind nature’s health benefits, and how to optimise these interactions is crucial for refining NBAs.

Furthermore, extending this research internationally and exploring the motivations driving AHPs’ use of NBAs can provide valuable insights into global practices and identify potential barriers to NBA integration, along with potential solutions to address challenges and improve practice. This deeper understanding, acquired through co-design methodologies, can guide the development of strategies aimed at enhancing the adoption and effectiveness of NBAs in allied health disciplines worldwide.

## Conclusion

A wide variety of NBAs are being used by AHPs in Australia. While NBAs are being embraced for their multifaceted benefits across physical and mental health, social and developmental health outcomes, challenges persist. These challenges include issues related to patient/ client engagement, AHP readiness, and external factors, like the weather and institutional constraints. Addressing these challenges calls for a collective effort to further legitimise NBAs, linking closely with the need for more targeted research, improved education and training, enhancing access to natural spaces and supportive governance structures.

Future research should aim to deepen the evidence base on NBAs’ safety and efficacy, explore their use beyond Australian borders, and engage more in nuanced investigations on how to enact innovative approaches suggested by the respondents in this study through methodologies like co-design. By tackling these challenges and building on the strong foundation of existing practices, we can expand the research and impact of NBAs, benefiting a wider array of patients/ clients, and the broader community.

## Data Availability

All data produced in the present study are available upon reasonable request to the authors

## Acknowledgements

The authors would like to thank Filip Maric, Adam Toner, Abby Tabor and Kexun Kenneth Chen for their feedback on the questionnaire, and the Australian Physiotherapy Association, Occupational Therapy Australia, Speech Pathology Australia, Australian Psychological Society, Exercise & Sports Science Australia, and the Australian Association of Social Workers for their assistance with participant recruitment.

## References

[1] Núñez MBF, Suzman LC, Maneja R, Bach A, Marquet O, Anguelovski I, Knobel P. Gender and sex differences in urban greenness’ mental health benefits: A systematic review. Health & Place 2022;76:102864.

[2] Bray I, Reece R, Sinnett D, Martin F, Hayward R. Exploring the role of exposure to green and blue spaces in preventing anxiety and depression among young people aged 14–24 years living in urban settings: A systematic review and conceptual framework. Environmental Research 2022;214:114081.

[3] Liu Z, Chen X, Cui H, Ma Y, Gao N, Li X, Meng X, Lin H, Abudou H, Guo L and others. Green space exposure on depression and anxiety outcomes: A meta-analysis. Environmental Research 2023;231:116303.

[4] Meo SA, Al-Khlaiwi T, Aqil M. Impact of the residential green space environment on the prevalence and mortality of Type 2 diabetes mellitus. European Review for Medical and Pharmacological Sciences 2022;26:3599–606.

[5] Liu XX, Ma XL, Huang WZ, Luo YN, He CJ, Zhong XM, Dadvand P, Browning MHEM, Li L, Zou XG and others. Green space and cardiovascular disease: A systematic review with meta-analysis. Environmental Pollution 2022;301:118990.

[6] Sillman D, Rigolon A, Browning MHEM, Yoon HV, McAnirlin O. Do sex and gender modify the association between green space and physical health? A systematic review. Environmental Research 2022;209:112869.

[7] Sprague NL, Bancalari P, Karim W, Siddiq S. Growing up green: a systematic review of the influence of greenspace on youth development and health outcomes. Journal of Exposure Science and Environmental Epidemiology 2022;32:660–81.

[8] Astell-Burt T, Hartig T, Putra IGNE, Walsan R, Dendup T, Feng X. Green space and loneliness: A systematic review with theoretical and methodological guidance for future research. Science of the Total Environment 2022;847:157521.

[9] Song Y, Li H, Yu H. Effects of green space on physical activity and body weight status among Chinese adults: a systematic review. Frontiers in Public Health 2023;11:1198439.

[10] Díaz-Martínez F, Sánchez-Sauco MF, Cabrera-Rivera LT, Sánchez CO, Hidalgo-Albadalejo MD, Claudio L, Ortega-García JA. Systematic review: Neurodevelopmental benefits of active/passive school exposure to green and/or blue spaces in children and adolescents. International Journal of Environmental Research and Public Health 2023;20:3958.

[11] Ccami-Bernal F, Soriano-Moreno DR, Fernandez-Guzman D, Tuco KG, Castro-Díaz SD, Esparza-Varas AL, Medina-Ramirez SA, Caira-Chuquineyra B, Cortez-Soto AG, Yovera-Aldana M and others. Green space exposure and type 2 diabetes mellitus incidence: A systematic review. Health & Place 2023;82:103045.

[12] Patwary MM, Bardhan M, Browning MHEM, Astell-Burt T, van der Bosch M, Dong J, Dzhambov AM, Dadvand P, Fasolino T, Markevych I and others. The economics of nature’s healing touch: A systematic review and conceptual framework of green space, pharmaceutical prescriptions, and healthcare expenditure associations. Science of the Total Environment 2024;914:169635.

[13] Luque-García L, Muxika-Legorburu J, Mendia-Berasategui O, Lertxundi A, García-Baquero G, Ibarluzea J. Green and blue space exposure and non-communicable disease related hospitalizations: A systematic review. Environmental Research 2023;245:118059.

[14] Marini S, Mauro M, Grigoletto A, Toselli S, Latessa PM. The effect of physical activity interventions carried out in outdoor natural blue and green spaces on health outcomes: A systematic review. International Journal of Environmental Research and Public Health 2022;19:12482.

[15] Nguyen P-Y, Astell-Burt T, Rahimi-Ardabili H, Feng X. Effect of nature prescriptions on cardiometabolic and mental health, and physical activity: a systematic review. Lancet Planetary Health 2023;7:313–28.

[16] Briggs R, Morris PG, Rees K. The effectiveness of group-based gardening interventions for improving wellbeing and reducing symptoms of mental ill-health in adults: a systematic review and meta-analysis Journal of Mental Health 2023;32:787–804.

[17] Obeng JK, Kangas K, Stamm I, Tolvanen A. Promoting sustainable well-being through nature-based interventions for young people in precarious situations: Implications for social work. A systematic review. Journal of Happiness Studies 2023;24:2881–911.

[18] Bikomeye JC, Balza JS, Kwarteng JL, Beyer AM, Beyer KMM. The impact of greenspace or nature-based interventions on cardiovascular health or cancer-related outcomes: A systematic review of experimental studies. PLoS One 2022;17:e0276517.

[19] Taylor EM, Robertson N, Lightfoot CJ, Smith AC, Jones CR. Nature-based interventions for psychological wellbeing in long-term conditions: A systematic review. International Journal of Environmental Research and Public Health 2022;19:3214.

[20] Coventry PA, Brown JVE, Pervin J, Brabyn S, Pateman R, Breedvelt J, Gilbody S, Stancliffe R, McEachan R, White PCL. Nature-based outdoor activities for mental and physical health: Systematic review and meta-analysis. SSM – Population Health 2021;16:100934.

[21] Ma J, Lin P, Williams J. Effectiveness of nature-based walking interventions in improving mental health in adults: a systematic review. Current Psychology 2024;43:9521–39.

[22] Iancu SC, Hoogendoorn AW, Zweekhorst MBM, Veltman DJ, Bunders JFG, van Balkom AJLM. Farm-based interventions for people with mental disorders: a systematic review of literature. Disability and Rehabilitation 2015;37:379–88.

[23] Mygind L, Kjeldsted E, Hartmeyer R, Mygind E, Bølling M, Bentsen P. Mental, physical and social health benefits of immersive nature-experience for children and adolescents: A systematic review and quality assessment of the evidence. Health & Place 2019;58:102136.

[24] Panțiru I, Ronaldson A, Sima N, Dregan A, Sima R. The impact of gardening on well-being, mental health, and quality of life: an umbrella review and meta-analysis. Systematic Reviews 2024;13:45.

[25] Young A, Boddy J, O’Leary P, Mazerolle P. Trialling a nature-based intervention with men who perpetrate domestic and family violence. Trends & Issues in Crime and Criminal Justic 2023:676.

[26] Pringle G, Boddy J, Slattery M, Harris P. Adventure therapy for adolescents with complex trauma: A scoping review and analysis. Journal of Experiential Education 2022;46.

[27] Boddy J, Slattery M, Liang J, Gallagher H, Smith A, Agllias K. Psychosocial interventions situated within the natural environment with young people who have experienced trauma: A scoping review. The British Journal of Social Work 2021;51:1018–40.

[28] Fan MSN, Li WHC, Ho LLK, Phiri L, Choi KC. Nature-based interventions for autistic children. A systematic review and meta-analysis. JAMA Network Open 2023;6:e2346715.

[29] Millennium Ecosystem Assessment. Ecosystems and human well-being: Synthesis. Washington, DC: Island Press; 2005.

[30] Stanhope J, Maric F, Rothmore P, Weinstein P. Physiotherapy and ecosystem services: Improving the health of our patients, the population, and the environment. Physiotherapy Therapy and Practice 2023;39:227–40.

[31] Wilson EO. Biophilia. The human bond with other species. Cambridge (MA): Harvard University Press; 1986.

[32] Hickman C. Cheerful prospects and tranquil restoration: the visual experience of landscape as part of the therapeutic regime of the British asylum, 1800-60. History of Psychiatry 2009;20:425–41.

[33] Hickman C. ‘To brighten the aspect of our streets and increase the health and enjoyment of our city’: The National Health Society and urban green space in late-nineteenth century London. Landscape and Urban Planning 2013;118:112–9.

[34] Vibholm AP, Christensen JR, Pallesen H. Occupational therapists and physiotherapists experiences of using nature-based rehabilitation. Physiotherapy Theory and Practice 2023;39:529–39.

[35] Stanhope J, Weinstein P. Public health lessons from the COVID-19 pandemic: the importance of green spaces for vulnerable populations. Perspectives in Public Health 2022;142:145–6.

[36] World Health Organization. Geneva: World Health Organization –]; Available from: https://www.who.int/standards/classifications/international-classification-of-functioning-disability-and-health.

[37] Day AMB, Theurer JA, Dykstra AD, Doyle PC. Nature and the natural environment as health facilitators: the need to reconceptualize the ICF environmental factors. Disability and Rehabilitation 2012;34:2281–90.

[38] Mueller W, Milner J, Loh M, Vardoulakis S, Wilkinson P. Exposure to urban greenspace and pathways to respiratory health: An exploratory systematic review. Science of the Total Environment 2022;829:154447.

[39] Stanhope J, Breed MF, Weinstein P. Exposure to greenspaces could reduce the high global burden of pain. Environmental Research 2020;187:109641.

[40] McCluskey A, Ada L, Kelly PJ, Middleton S, Goodall S, Grimshaw JM, Logan P, Longworth M, Karageorge A. Compliance with Australian stroke guideline recommendations for outdoor mobility and transport training by post-inpatient rehabilitation services: An observational cohort study. BMC Health Services Research 2015;15:296.

[41] Kucher N, Larson JM, Fischer G, Mertaugh M, Peterson L, Gershan LA. 3-Dimensional nature-based therapeutics in pediatric patients with total pancreatectomy and islet auto-transplant. Complementary Therapies in Medicine 2020;48:102249.

[42] Moore L, Koger D, Blomberg S, Legg L, McConahy R, Wit S, Gatmaitan M. Making best practice our practice reflections on our journey into natural environments. Infants and Young Children 2012;25:95–105.

[43] James G, Kidd K, Cooley SJ, Fenton K. The feasibility of outdoor psychology sessions in an adult mental health inpatient rehabilitation unit: Service user and psychologist perspectives. Frontiers in Psychology 2021;12.

[44] Ayaz EY, Dincer B, Mete E, Benli RK, Cinbaz G, Karacan E, Cankül A, Mesci B. Evaluating the impact of aerobic and resistance green exercises on the fitness, aerobic and intrinsic capacity of older individuals. Archives of Gerontology and Geriatrics 2024;118:105281.

[45] Pijpker R, Veen EJ, Vaandrager L, Koelen M, Bauer GF. Developing an intervention and evaluation model of outdoor therapy for employee burnout: Unraveling the interplay between context, processes, and outcomes. Frontiers in Psychology 2022;13:785697.

[46] McCluskey A, Middleton S. Increasing delivery of an outdoor journey intervention to people with stroke: A feasibility study involving five community rehabilitation teams. Implementation Science 2010;5:59.

[47] Uldall SW, Poulsen DV, Christensen SI, Wilson L, Carlsson J. Mixing job training with nature-based therapy shows promise for increasing labor market affiliation among newly arrived refugees: Results from a danish case series study. International Journal of Environ Research and Public Health 2022;19:4850.

[48] Poulsen DV, Lygum VL, Djernis HG, Stigsdotter UK. Nature is just around us! Development of an educational program for implementation of nature-based activities at a crisis shelter for women and children exposed to domestic violence. Journal of Social Work Practice 2021;35:159–75.

[49] Overbey TA, Diekmann F, Lekies KS. Nature-based interventions for vulnerable youth: a scoping review. International Journal of Environmental Health Research 2023;33:15–53.

[50] Wang Y, Gao Q, Pei F, Wang Y, Cheng Z, Zhang J, Wu Y. Innovative technology-enhanced social work service during COVID-19: How ‘Garden on the Balcony’ promoted resilience, community bonds and a green lifestyle. Qualitative Social Work 2023;22:321–39.

[51] Hauge ÅL, Lindheim M, Røtting K, Johnsen SÅK. The meaning of the physical environment in child and adolescent therapy: A qualitative study of the outdoor care retreat. Ecopsychology 2023;15:244–58.

[52] Yıldırım Ayaz E, Dincer B, Mete E, Kaygusuz Benli R, Cinbaz G, Karacan E, Cankül A, Mesci B. Evaluating the impact of aerobic and resistance green exercises on the fitness, aerobic and intrinsic capacity of older individuals. Archives of Gerontology and Geriatrics 2024;118:105281.

[53] Firby H, Raine R. Engaging with nature and the outdoors: A scoping review of therapeutic applications in contemporary occupational therapy. British Journal of Occupational Therapy 2022;86.

[54] Peters E, Hovinga D, Maas J, Schuengel C. Social workers’ choice making in supporting nature activities by parents and children in shelters. Frontiers in Psychology 2022;13:891419.

[55] Wagenfeld A, Atchison B. “Putting the occupation back in occupational therapy:” a survey of occupational therapy practitioners’ use of gardening as an intervention. The Open Journal of Occupational Therapy 2014;2.

[56] Tambyah R, Olcoń K, Allan J, Destry P, Astell-Burt T. Mental health clinicians’ perceptions of nature-based interventions within community mental health services: evidence from Australia. BMC Health Services Research 2022;22:841.

[57] Cooley SJ, Taylor E, Ceslikauskaite K, Robertson N. Outdoor therapy: Maverick or mainstream? A survey of clinical psychologists. Clinical Psychology Forum 2023;1:21–31.

[58] Cooley SJ, Jones CR, Moss D, Robertson N. Organizational perspectives on outdoor talking therapy: Towards a position of ‘environmental safe uncertainty’. British Journal of Clinical Psychology 2022;61:132–56.

[59] Stanhope J, Foley K, Butler M, Boddy J, Clanchy K, George E, Roberts R, Rothmore P, Salter A, Serocki P and others. “A variety of green spaces available to all”: knowledge of patient and community needs in natural spaces as understood by allied health professionals medRxiv 2024.

[60] Thomas DR. A general inductive approach for analysing qualitative evaluation data. American Journal of Evaluation 2006;27:237–46.

[61] Meyer SB, Lunnay B. The application of abductive and retroductive inference for the design and anaysis of theory-driven sociological research. Sociological Research Online 2013;18:86–96.

[62] Fletcher AJ. Applying critical realism in qualitative research: methodology meets method. International Journal of Social Research Methodology 2017;20:181–94.

[63] Ye T, Yu P, Wen B, Yang Z, Huang W, Yuming G, Abramson MJ, Li S. Greenspace and health outcomes in children and adolescents: A systematic review. Environmental Pollution 2022;314:120193.

[64] Twohig-Bennett C, Jones A. The health benefits of the great outdoors: A systematic review and meta-analysis of greenspace exposure and health outcomes Environmental Research 2018;166:628–37.

[65] Browning MHEM, Shipley N, McAnirlin O, Becker D, Yu CP, Hartig T, Dzhambov AM. An actual natural setting improves mood better than its virtual counterpart: a meta-analysis of experimental data. Frontiers in Psychology 2020;11:2200.

[66] Lundgren K. Nature-based therapy: its potential as a complementary approach to treating communication disorders. Semin Speech Lang 2004;25:121–31.

[67] Ellingsen-Dalskau LH, Berget B, Pedersen I, Tellnes G, Ihlebæk C. Understanding how prevocational training on care farms can lead to functioning, motivation and well-being Disability and Rehabilitation 2016;38:2504–13.

[68] Joy YS, Lee A-Y, Park S-A. A horticultural therapy program focused on succulent cultivation for the vocational rehabilitation training of individuals with intellectual disabilities. International Journal of Environmental Research and Public Health 2020;17:1303.

[69] Robinson JM, Aronson J, Dnaiels CB, Goodwin N, Liddicoat C, Orlando L, Phillips D, Stanhope J, Weinstein P, Cross AT and others. Ecosystem restoration is integral to humanity’s recovery from COVID-19. The Lancet Planetary Health 2022;6:769–73.

[70] Timler K, Sandy DW. Gardening in ashes: the possibilities and limitations of gardening to support indigenous health and well-being in the context of wildfires and colonialism. International Journal of Environmental Research and Public Health 2020;17:3273.

[71] Venn TJ. Economic implications of inalienable and communal native title: The case of Wik forestry in Australia. Ecological Economics 2007;64.

[72] Astell-Burt T, Feng X. Time for ‘green’ during COVID-19? Inequities in green and blue space access, visitation and felt benefits. International Journal of Environmental Research and Public Health 2021;18:2757.

[73] Environmental Physiotherapy Association. 2024 –]; Available from: https://environmentalphysio.com/.

[74] Maric F, Chance-Larsen K, Chevan J, Jameson S, Nicholls D, Opsommer E, Perveen W, Richter R, Stanhope J, Stone O and others. A progress report on planetary health, environmental and sustainability education in physiotherapy – Editorial. European Journal of Physiotherapy 2021;23:201–2.

[75] Environmental Physiotherapy Association. Environmental Physiotherapy Association; 2024 –]; Available from: https://environmentalphysio.com/education/inspirational-case-reports/.

[76] Weinstein P, Bi P, Stanhope J. Climate change adaptation must not replicate lockdown scenarios. Perspectives in Public Health 2024.

[77] James L, Shing J, Mortenson WB, Mattie J, Borisoff J. Experiences with and perceptions of an adaptive hiking program. Disability and Rehabilitation 2017;40:1584–90.

[78] Torrado JC, Jaccheri L, Pelagatti S, Wold I. HikePal: A mobile exergame to motivate people with intellectual disabilities to do outdoor physical activities. Entertainment Computing 2022;42:100477.

[79] Buettel JC, Brook BW. Egress! How technophilia can reinforce biophilia to improve ecological restoration. Restoration Ecology 2016;24:843–7.

