## Supplementary Material for "Nature-based allied health: current practice, challenges and opportunities"

**Supplementary Material 1: Nature-based allied health questionnaire** **2**
**Supplementary Material 2: Discipline-specific findings** **35**
Occupational therapy 35
Physiotherapy/ exercise physiology 44
Psychology 53
Social work 62
Speech pathology 71

### Supplementary Material 1: Nature-based allied health questionnaire

*Note: This is the questionnaire used for the Stage 2 data collection. The Stage 1 questionnaire had some differences, however all aspects reported in the paper relate to questions that were the same across Stages 1 and 2*

#### Demographic questions

***We will ask you some questions related to yourself. Please note, you can choose to skip any question if you would prefer not to respond.***

1. What is your age? (in years)

2. What is your gender?

3. For you first health qualification:

    What is the name of the qualification?

    In which year did you obtain the qualification?

    In which country did you do this training?

4. Have you obtained another health qualification?

    ☐ Yes

    ☐ No (skip to Question 13)

5. For you second health qualification:

    What is the name of the qualification?

    In which year did you obtain the qualification?

    In which country did you do this training?

6. Have you obtained another health qualification?

    ☐ Yes

    ☐ No (skip to Question 13)

7. For you third health qualification:

What is the name of the qualification?

In which year did you obtain the qualification?

In which country did you do this training?

8. Have you obtained another health qualification?

☐ Yes

☐ No (skip to Question 13)

9. For you fourth health qualification:

What is the name of the qualification?

In which year did you obtain the qualification?

In which country did you do this training?

10. Have you obtained another health qualification?

☐ Yes

☐ No (skip to Question 13)

11. For you fifth health qualification:

What is the name of the qualification?

In which year did you obtain the qualification?

In which country did you do this training?

12. Have you obtained another health qualification?

☐ Yes

☐ No

### Discipline

13. Which is your main discipline?

☐ Exercise physiology (skip to Question 49)

☐ Occupational therapy (skip to Question 25)

☐ Physiotherapy

☐ Psychology (skip to Question 57)

☐ Social work (skip to Question 41)

☐ Speech pathology (skip to Question 33)

### Physiotherapy

14. Are you registered as another type of health professional?

☐ No

☐ Yes (please specify):

15. For how many years (full time equivalent) have you worked in any capacity as a physiotherapist? *This includes physiotherapy roles which do not involve working with patients/ clients*

16. In this last 12 months, what has been your principal physiotherapy role?

☐ Clinician

☐ Administration

☐ Teacher/ education

☐ Researcher

☐ Other (please specify)

17. Which ONE age group of patients/ clients have you worked with the most in the last 12 months?

☐ Children (0-12 years)

☐ Adolescents (13-17 years)

☐ Adults (18-74 years)

☐ Older adults (75 years or older)

18. In the last 12 months, in which ONE physiotherapy scope of practice have you worked the most?

☐ Musculoskeletal

☐ Aged care

☐ Neurological

☐ Cardiorespiratory

☐ Paediatrics

☐ Sports

☐ Women's health

☐ Other (please specify):

19. In the last 12 months, which ONE physiotherapy stream have you practiced in the most?

☐ Rehabilitation

☐ Acute care

☐ Community based care

☐ Aged care

☐ Chronic disease management

☐ Cancer care

☐ Palliative care

☐ Mental health

☐ Other (please specify):

20. Are you an Australian Physiotherapy Association (APA) titled physiotherapist?

☐ No

☐ Yes (please specify the title):

21. Are you a specialist physiotherapist (as awarded by the Australian College of Physiotherapists)?

☐ No

☐ Yes (please specify your area of specialty):

22. In the last 12 months, which sector have you worked in the most?

☐ Public

☐ Private

23. In the last 12 months which ONE setting have you worked in the most as a physiotherapist?

☐ Group private practice

☐ Solo private practice

☐ Hospital

☐ Outpatient service

☐ Residential aged care facility

☐ Rehabilitation/ physical development service

☐ Educational facility

☐ Sports centre/ clinic

☐ Domiciliary service

☐ Other community health care service

☐ Other (please specify):

24. In the last 12 months have you worked directly with patients/ clients as a physiotherapist (including tele-health)?

☐ Yes (skip to Question 66)

☐ No (skip to Question 71)

**Occupational therapy**

25. Are you registered as another type of health professional?

☐ No

☐ Yes (please specify):

26. For how many years (full time equivalent) have you worked in any capacity as an occupational therapist? *This includes occupational therapy roles which do no involve working with patients/ clients*

27. In the last 12 months, what has been your principal occupational therapy role?

☐ Clinician

☐ Administration

☐ Teacher/ education

☐ Researcher

☐ Other (please specify):

28. Which ONE age group of patients/ clients have you worked with the most in the last 12 months?

☐ Children (0-12 years)

☐ Adolescents (13-17 years)

☐ Adult (18-74 years)

☐ Older adults (75 years or older)

29. In the last 12 months, which ONE occupational therapy scope have you worked in the most?

☐ Rehabilitation

☐ Paediatrics

☐ Aged care

☐ Mental health

☐ Disability

☐ Occupational health

☐ Hand therapy

☐ Neurological

☐ Driving assessment

☐ Other (please specify):

30. In the last 12 months, which sector have you worked in the most?

☐ Public

☐ Private

31. In the last 12 months which ONE setting have you worked in the most as an occupational therapist?

☐ Group private practice

☐ Solo private practice

☐ Hospital

☐ Outpatient service

☐ Residential aged care facility

☐ Rehabilitation/ physical development service

☐ Educational facility

☐ Disability service

☐ Other government department or agency

☐ Other community health care service

☐ Other (please specify):

32. In the last 12 months have you worked directly with patients/ clients as an occupational therapist (including tele-health)?

☐ Yes (skip to Question 66)

☐ No (skip to Question 71)

**Speech pathology**

33. Are you registered as another type of health professional?

☐ No

☐ Yes (please specify):

34. For how many years (full time equivalent) have you worked in any capacity as a speech pathologist? *This includes speech pathology roles which do not involve working with patients/ clients*

35. In the last 12 months, what has been your principal speech pathology role?

☐ Clinician

☐ Administration

☐ Teacher/ education

☐ Researcher

☐ Other (please specify):

36. Which ONE age group of patients/ clients have you worked with the most in the last 12 months?

☐ Children (0-12 years)

☐ Adolescents (13-17 years)

☐ Adults (18-74 years)

☐ Older adults (75 years or older)

37. In the last 12 months, in which ONE area of speech pathology practice have you worked the most?

☐ Speech and language

☐ Literacy

☐ Fluency

☐ Voice

☐ Communication and multimodal communication

☐ Swallowing and mealtimes

☐ Other (please specify):

38. In the last 12 months, which sector have you worked in the most?

☐ Public

☐ Private

39. In the last 12 months, which ONE setting have you worked in the most as a speech pathologist?

☐ Group private practice

☐ Solo private practice

☐ Hospital

☐ Outpatient service

☐ Residential aged care facility

☐ Rehabilitation/ physical development service

☐ Educational facility

☐ Disability service

☐ Other government department or agency

☐ Other community health care service

☐ Other (please specify):

40. In the last 12 months, have you worked directly with patients/ clients as a speech pathologist (including tele-health)?

☐ Yes (skip to Question 66)

☐ No (skip to Question 71)

**Social work**

41. Are you registered as another type of health professional?

☐ No

☐ Yes (please specify):

42. For how many years (full time equivalent) have you worked in any capacity as a social worker? *This includes social work roles which do not involve working with patients/ clients*

43. In the last 12 months, what has been your principal social work role?

☐ Clinician

☐ Administration

☐ Teacher/ education

☐ Researcher

☐ Other (please specify):

44. Which ONE age group of patients/ clients have you worked with the most in the last 12 months?

☐ Children (0-12 years)

☐ Adolescents (13-17 years)

☐ Adults (18-74 years)

☐ Older adults (75 years or older)

45. In the last 12 months, in which ONE social work field of practice have you worked in the most?

☐ Mental health

☐ Health

☐ Disability

☐ Child protection

☐ Education

☐ Family support

☐ Youth

☐ Family violence

☐ Aged care

☐ Alcohol, tobacco and other drugs

☐ Sexual violence

☐ Housing

☐ Income support

☐ Management

☐ Refugees/ asylum seekers

☐ Culturally and linguistically diverse communities

☐ Palliative care

☐ Veteran affairs

☐ Other (please specify):

46. In the last 12 months, which sector have you worked in the most?

☐ Public

☐ Private

47. In the last 12 months, which ONE setting have you worked in the most as a social worker?

- ☐ Group private practice
- ☐ Solo private practice
- ☐ Hospital
- ☐ Outpatient service
- ☐ Residential aged care facility
- ☐ Rehabilitation/ physical development service
- ☐ Educational facility
- ☐ Disability service
- ☐ Other government department or agency
- ☐ Other community health care service (including non-government agency)
- ☐ Other (please specify):

48. In the last 12 months, have you worked directly with patients/ clients as a social worker (including tele-health)?

- ☐ Yes (skip to Question 66)
- ☐ No (skip to Question 71)

### Exercise physiology

49. Are you registered as another type of health professional?

- ☐ No
- ☐ Yes (please specify):

50. For how many years (full time equivalent) have you worked in any capacity as an exercise physiologist? *This includes exercise physiology roles which do not involve working with patients/ clients*

51. In the last 12 months, what has been your principal exercise physiology role?

☐ Clinician

☐ Administration

☐ Teacher/ education

☐ Researcher

☐ Other (please specify):

52. Which ONE age group of patients/ clients have you worked with the most in the last 12 months?

☐ Children (0-12 years)

☐ Adolescents (13-17 years)

☐ Adults (18-74 years)

☐ Older adults (75 years or older)

53. Which ONE age below best describes your work?

☐ Musculoskeletal

☐ Metabolic

☐ Neurological

☐ Cardiovascular

☐ Respiratory

☐ Cancer

☐ Sports

☐ Mental health

☐ Occupational health

☐ Mixed

☐ Other (please specify):

54. In the last 12 months, which sector have you worked in the most?

☐ Public

☐ Private

55. In the last 12 months, which ONE work sector best describes you work as an exercise physiologist?

☐ Federal government organisation

☐ State government organisation

☐ Workers' compensation agency

☐ Fitness club or institution

☐ Sporting club or institution

☐ Health care organisation

☐ Hospital

☐ Mining

☐ Private company

☐ Research or education institution

☐ Other (please specify):

56. In the last 12 months, have you worked directly with patients/ clients as an exercise physiologist (including tele-health)?

☐ Yes (skip to Question 66)

☐ No (skip to Question 71)

### Psychology

57. Are you registered as another type of health professional?

☐ No

☐ Yes (please specify):

58. Which area(s) of practice do you have endorsement for? *(tick all that apply)*

☐ Clinical psychology

☐ Counselling psychology

☐ Clinical neuropsychology

☐ Education and developmental psychology

☐ Forensic psychology

☐ Organisational psychology

☐ Health psychology

☐ Sport and exercise psychology

☐ Community psychology

☐ No areas of practice endorsement

59. For how many years (full time equivalent) have you worked in any capacity as a psychologist? *This includes psychology roles which do not involve working with patients/ clients*

60. In the last 12 months, what has been your principal psychologist role?

☐ Clinician

☐ Administration

☐ Teacher/ education

☐ Research

☐ Other (please specify):

61. Which ONE age group of patients/ clients have you worked with the most in the last 12 months?

☐ Children (0-12 years)

☐ Adolescents (13-17 years)

☐ Adults (18-74 years)

☐ Older adults (75 years or older)

62. In the last 12 months, in which ONE psychology job area have you worked the most?

☐ Counselling

☐ Mental health intervention

☐ Neuropsychology/ cognitive assessment

☐ Psychology management/ administration

☐ Consulting/ advising for work

☐ Behavioural assessments

☐ Physical health/ rehabilitation

☐ Community engagement

☐ Health promotion

☐ Personal development/ coaching

☐ Organisational practices

☐ Recruitment

☐ Training for work purposes

☐ Medico-legal assessment

☐ Teaching/ supervision

☐ Research projects

☐ Other (please specify):

63. In the last 12 months, which sector have you worked in the most?

☐ Public

☐ Private

64. In the last 12 months, which ONE setting have you worked in the most as a psychologist?

- ☐ Solo private practice
- ☐ Group private practice
- ☐ GP practice
- ☐ Hospital
- ☐ Community mental health service
- ☐ Tertiary educational facility
- ☐ School
- ☐ Commercial business/ service
- ☐ Other government department
- ☐ Other private practice
- ☐ Remaining work settings
- ☐ Other (please specify):

65. In the last 12 months, have you worked directly with patients/ clients as a psychologist (including tele-health)?

- ☐ Yes
- ☐ No (skip to Question 71)

### Practice location

***The remaining questions refer to your work in your main practice setting (e.g. hospital, private practice)***

66. Do you solely do home visits?

- ☐ Yes (skip to Question 69)
- ☐ No

**Setting details**

67. For this main practice setting, which of the following are within 50m of the building? *(tick all that apply)*

- ☐ Grass/ lawn
- ☐ Garden
- ☐ Tree
- ☐ Water bodies (e.g. lake, dam, river, creek, ocean)
- ☐ Other open space (please describe):

68. What is the postcode of your main practice setting?

**Work details**

69. In the last 12 months, how many hours per week have you worked on average in your main practice setting (e.g. private practice)?

70. In the last 12 months, have you used any form of telehealth?

- ☐ Yes
- ☐ No

### Nature-based exposures (for those who do not work directly with patients/ clients)

***We will now ask you some questions about whether you think people could benefit from spending time in outdoor natural environments, and/or indoor nature-based exposures***

71. Do you think people could benefit from spending time in outdoor natural environment? *Outdoor natural environments may include parks, gardens, farms, lakes, or the ocean.*

☐ No

☐ Yes. Which signs, symptoms, or conditions do you think might be prevented and/or managed by spending time in outdoor natural environments?

72. Do you think people could benefit from indoor nature-based exposures? *For example, nature pictures, sounds, scents, virtual reality, driving through natural environments*

☐ No (skip to Question 98)

☐ Yes. Which signs, symptoms, or conditions do you think might be prevented and/or managed through indoor nature-based exposures? (skip to Question 98)

### Outdoor natural environments

***Now, we will ask you some questions about using outdoor natural environments in therapy, in your main practice setting. Outdoor natural environments may include parks, gardens, farms, lacks, or the ocean***

73. Do you think people could benefit from spending time in outdoor natural environment? *Outdoor natural environments may include parks, gardens, farms, lakes, or the ocean.*

☐ No

☐ Yes. Which signs, symptoms, or conditions do you think might be prevented and/or managed by spending time in outdoor natural environments?

### Outdoor natural environments: recommended activities

***First, we are going to ask you specifically about the activities you have recommended be performed in outdoor natural environments outside of the therapy session***

74. In the last 12 months, which activities have you **recommended** your patients/ clients perform in outdoor natural environments outside of their therapy sessions? *(tick all that apply)*

☐ Land-based gross motor activities (e.g. aerobic, resistance, balance)

☐ Land-based fine motor activities (e.g. finger exercises, craft)

☐ Water-based physical activities (i.e. in or on water)

☐ Active transport (e.g. e-bikes, e-scooters)

☐ Powered transport (e.g. electric wheelchair)

☐ Gardening

☐ Ecological conservation or restoration

☐ Engaging with/ tending to animals (e.g. care, leisure or hobby farming)

☐ Wilderness activities (e.g. camping, rock climbing)

☐ Nature play

☐ Nature observation (e.g. bird watching, photography)

☐ Mindfulness/ meditation/ relaxation

☐ Forest bathing

☐ Social activities

☐ Spending time in outdoor natural environments (no specific activity)

☐ Other (please describe)

☐ No activities in outdoor natural environments

75. In the last 12 months, which outdoor natural environments have you recommended your patients/ clients spend time in outside of their therapy sessions? *(tick all that apply)*

☐ Private garden

☐ Community/ school garden

☐ Farm (including hobby, leisure and care farms)

☐ Public park

☐ National park

☐ Forest

☐ Oval/ sportsfield

☐ On/ in the lake

☐ Next to a lack

☐ On/ in the river

☐ Next to a river

☐ On/ in the ocean

☐ Next to the ocean

☐ Other (please describe):

76. In the last 12 months, have you recommended other health professionals or services (e.g. council, community groups) to assist your patients/ clients in spending time in outdoor natural environments?

☐ No

☐ Yes (please specify):

77. Has the COVID-19 pandemic influenced your recommendations to your patients/ clients to spend time in outdoor natural environments outside of their therapy sessions?

☐ Yes, increased recommendations

☐ Yes, decreased recommendations

☐ No change

78. Are there any outdoor activities in nature you would have liked to have recommended to patients/ clients outside of their therapy sessions in the last 12 months, but didn't?

☐ No

☐ Yes (please describe):

79. In the last 12 months, have you experienced any challenges related to recommending that your patients/ clients spend time in outdoor natural environments?

☐ No (skip to Question 81)

☐ Yes. Please describe the challenges.

80. What solutions would you like to see implemented to overcome these challenges?

**Outdoor natural environments: activities with your patients/ clients**

81. In the last 12 months, which activities have you **worked directly with your patients/ clients** to perform in outdoor natural environments (excluding tele-health)? *(tick all that apply)*

☐ Land-based gross motor activities (e.g. aerobic, resistance, balance)

☐ Land-based fine motor activities (e.g. finger exercises, craft)

☐ Water-based physical activities (i.e. in or on water)

☐ Active transport (e.g. e-bikes, e-scooters)

☐ Powered transport (e.g. electric wheelchair)

☐ Gardening

☐ Ecological conservation or restoration

☐ Engaging with/ tending to animals (e.g. care, leisure or hobby farming)

☐ Wilderness activities (e.g. camping, rock climbing)

☐ Nature play

☐ Nature observation (e.g. bird watching, photography)

☐ Mindfulness/ meditation/ relaxation

☐ Forest bathing

☐ Social activities

☐ Spending time in outdoor natural environments (no specific activity)

☐ Other (please describe):

☐ None

82. In the last 12 months, in which outdoor natural environments have you worked directly with your patients/ clients (excluding tele-health)? *(tick all that apply)*

☐ Private garden

☐ Community/ school garden

☐ Farm (including hobby, leisure and care farms)

☐ Public park

☐ National park

☐ Forest

☐ Oval/ sportsfield

☐ On/ in the lake

☐ Next to a lack

☐ On/ in the river

☐ Next to a river

☐ On/ in the ocean

☐ Next to the ocean

☐ Other (please describe):

83. Has the COVID-19 pandemic influenced the amount of time you have spent working in outdoor natural environments with your patients/ clients?

☐ Yes, increased time

☐ Yes, decreased time

☐ No change

84. Are there any outdoor activities in nature you would have liked to have incorporated into therapy sessions with patients/ clients in the last 12 months, but didn't?

☐ No

☐ Yes (please specify):

85. In the last 12 months, have you experienced any challenges related to engaging directly with your patients/ clients in outdoor natural environments?

☐ No (skip to Question 87)

☐ Yes. Please describe the challenges.

86. What solutions would you like to see implemented to overcome these challenges?

**Indoor nature-based exposures: recommended activities**

***Now, we will ask you some questions about your use of indoor nature-based exposures in therapy.***

87. Do you think people could benefit from indoor nature-based exposures? *For example, nature pictures, sounds, scents, virtual reality, driving through natural environments*

☐ No

☐ Yes. Which signs, symptoms or conditions do you think might be prevented and/ or managed through indoor nature-based exposures? (please describe):

88. In the last 12 months, which of the following indoor nature-based exposures have you **recommended** to your patients/ clients? *(tick all that apply)*

☐ Listening to natural sounds

☐ Having indoor pot plants

☐ Pictures of natural scenes

☐ Natural scents (e.g. infusers, tea bags)

☐ Traveling in care or on public transport through natural environments

☐ Views of nature outside the window

☐ Opening windows for fresh air and the smells and sounds of nature

☐ Nature-based virtual reality

☐ Other (please describe):

☐ None

89. Has the COVID-19 pandemic influenced your recommendations of indoor nature-based exposure to your patients/ clients?

☐ Yes, increased recommendations

☐ Yes, decreased recommendations

☐ No change

90. Are there any indoor nature-based activities you would have liked to have recommended to patients/ clients outside of their therapy sessions in the last 12 months, but didn't?

☐ No

☐ Yes (please describe):

91. In the last 12 months, have you experienced any challenges related to recommending that your patients/ clients spend time in indoor nature-based exposures?

☐ No (skip to Question 93)

☐ Yes. Please describe the challenges.

92. What solutions would you like to see implemented to overcome these challenges?

**Indoor nature-based exposures: activities with your patients or clients**

93. In the last 12 months, which of the following indoor nature-based exposures have you used ***with your patients/ clients*** in therapy sessions (excluding tele-health)? *(tick all that apply)*

☐ Listening to natural sounds

☐ Having indoor pot plants

☐ Pictures of natural scenes

☐ Natural scents (e.g. infusers, tea bags)

☐ Traveling in car or on public transport through natural environments

☐ Views of nature outside the window

☐ Opening winds for fresh air and the smalls and sounds of nature

☐ Nature-based virtual reality

☐ Other (please describe):

☐ None

94. Has the COVID-19 pandemic influences the time you have spent using indoor nature-based exposures with your patients/ clients in therapy sessions (excluding tele-health)?

☐ Yes, increased time

☐ Yes, decreased time

☐ No change

95. Are there any indoor nature-based activities you would have liked to have incorporated into therapy sessions with your patients/ clients in the last 12 months, but didn't?

☐ No

☐ Yes (please describe):

96. In the last 12 months, have you experienced any challenges in using indoor nature-based exposures with your patients/ clients in therapy sessions?

☐ No (skip to Question 98)

☐ Yes. Please describe the challenges.

97. What solutions would you like to see implemented to overcome these challenges?

### Public outdoor natural spaces

***We will now ask you some questions about the provision and/or optimisation of outdoor natural environments***

98. What features do you think are important in outdoor natural environments for your patients/ clients and the broader community?

### Public outdoor natural spaces: advocacy

99. In the last 12 months, which of the following outdoor natural environments have you personally or professionally **advocated** to have provided or improved? *Improvements may include improvements to access, activities available, amenities, and biodiversity (tick all that apply)*

- ☐ Gardens or other outdoor natural elements in care settings (e.g. hospital, aged care, clinics)
- ☐ Natural elements inside care settings
- ☐ Natural elements inside non-care, public settings (e.g. workplaces)
- ☐ Community gardens
- ☐ School/ childcare gardens
- ☐ Public parks/ gardens
- ☐ Green corridors (e.g. around streets, paths)
- ☐ Blue spaces/ water bodies
- ☐ Other (please describe):

- ☐ None

100. Are there any improvements to outdoor natural spaces you would have liked to have advocated for in the last 12 months, but didn't?

☐ No

☐ Yes (please describe):

101. Has the COVID-19 pandemic influenced your advocacy for the provision of or improvement to outdoor natural environments?

☐ Yes, increased advocacy

☐ Yes, decreased advocacy

☐ No change

102. In the last 12 months, have you experienced any challenges related to advocating for the provision of or improvement to outdoor natural environments?

☐ No (skip to Question 104)

☐ Yes. Please describe the challenges.

103. What solutions would you like to see implemented to overcome these challenges?

### Public outdoor natural spaces: direct involvement

104. In the last 12 months, which of the following outdoor natural environments have you personally or professional been **directly involved in** the provision of and/or improvement to? *Improvements may include improvements to access, activities available, amenities, and biodiversity (tick all that apply)*

- ☐ Gardens or other outdoor natural elements in care settings (e.g. hospitals, aged care, clinics)
- ☐ Natural elements inside care settings
- ☐ Natural elements inside non-care, public settings (e.g. workplaces)
- ☐ Community gardens
- ☐ School/ childcare gardens
- ☐ Public parks/ gardens
- ☐ Green corridors (e.g. around streets, paths)
- ☐ Blue spaces/ water bodies
- ☐ Other (please describe):
- ☐ None

105. Are there any improvements to outdoor natural spaces you would have liked to have been directly involved in during the last 12 months, but didn't?

- ☐ No
- ☐ Yes (please describe)

106. Has the COVID-19 pandemic influences the amount of time you have spent directly involved in the provision of or improvement to outdoor natural environments?

☐ Yes, increased time

☐ Yes, decreased time

☐ No change

107. In the last 12 months, have you experienced challenges in your direct involvement in the provision of or improvement to outdoor natural environments?

☐ No (end of survey)

☐ Yes. Please describe the challenges.

108. What solutions would you like to see implemented to overcome these challenges?

End of survey

### Supplementary Material 2: Discipline-specific results

The results reported here are discipline specific. Ticks indicate that at least one participant engaged in the activity. Given the relatively small sample sizes, absence of evidence from our sample that health professionals have engaged in the specific activity should not be interpreted as evidence that these activities have not been undertaken by allied health professionals.

#### Occupational therapy

**Table S2.1: Occupational therapy respondent demographics**

| | Stage 1 (n=4) | Stage 2 (n=9) |
|---|---|---|
| Gender | | |
| Woman | 4/4 | 8/9 |
| Man | 0/4 | 1/9 |
| Age in years, median (interquartile range) | | |
| 20-29 years | 2/4 | 2/9 |
| 30-39 years | 1/4 | 1/9 |
| 40-49 years | 0/4 | 2/9 |
| 50-59 years | 0/4 | 2/9 |
| 60+ years | 1/4 | 2/9 |
| Trained in Australia (pre-registration qualification) | 4/4 | 7/9 |
| Years worked in discipline, median (interquartile range) | 10 (2-23) | 10 (4-19) |
| Main patient age group | | |
| Children | NA | 4/9 |
| Adolescents | NA | 1/9 |
| Adults | NA | 4/9 |
| Older adults | NA | 0/9 |
| Role | | |
| Teacher/ educator | NA | 2/9 |
| Clinician | NA | 5/9 |
| Administration | NA | 1/9 |
| Discharge planning | NA | 1/9 |
| Scope | | |
| Disability | 0/4 | 1/9 |
| Occupational health | 0/4 | 2/9 |
| Acute care | 0/4 | 1/9 |
| Mental health | 0/4 | 1/9 |
| Paediatrics | 2/4 | 4/9 |
| Community | 1/4 | 0/9 |
| Education | 1/4 | 0/9 |
| Setting | | |
| Private practice | 2/4 | 5/9 |
| Hospital | 0/4 | 2/9 |
| Disability services | 0/4 | 1/9 |
| Community pharmacy | 0/4 | 1/9 |
| Community | 2/4 | 0/9 |
| Private sector | 3/3 | 3/9 |
| Sees patients/ clients | 3/4 | 7/9 |
| Telehealth | 2/3 | 5/7 |
| Home visits only | NA | 3/7 |
| Main practice in a major city^ | 3/3 | 4/4 |
| Hours worked in main practice setting, median (interquartile range) | 30 (30-34) | 30 (25-38) |

Note: NA: Not asked/ ^Stage 2 respondents were only asked for the postcode of their main practice if they did not solely conduct home visits.

### Recommendations

**Table S2.2: Recommendations for outdoor nature-based activities from the occupational therapists in our sample**

| Activity | Number from Stage 1 | Number from Stage 2 | Age groups^ | | | Scope | | | | Sector | | Setting | | | |
|---|---|---|---|---|---|---|---|---|---|---|---|---|---|---|---|
| | | | Children | Adolescents | Adults | Disability | Occupational health | Paediatrics | Community | Private | Public | Private practice | Disability services | Community pharmacy | Community setting |
| Any | 3/3 | 6/6 | ✓ | ✓ | ✓ | ✓ | ✓ | ✓ | ✓ | ✓ | ✓ | ✓ | ✓ | ✓ | ✓ |
| Land-based gross motor activities | 3/3 | 5/6 | ✓ | ✓ | ✓ | ✓ | ✓ | ✓ | ✓ | ✓ | ✓ | ✓ | | ✓ | ✓ |
| Land-based fine motor activities | 1/3 | 3/6 | ✓ | | | | | ✓ | ✓ | ✓ | ✓ | ✓ | ✓ | | |
| Water-based physical activities | 2/3 | 4/6 | ✓ | ✓ | | ✓ | | ✓ | ✓ | ✓ | ✓ | ✓ | ✓ | | ✓ |
| Active transport | 1/3 | 1/6 | | ✓ | | ✓ | | | ✓ | ✓ | | ✓ | | | |
| Powered transport | 1/3 | 0/6 | | | | | | | ✓ | ✓ | | ✓ | | | |
| Gardening | 2/3 | 2/6 | ✓ | ✓ | | ✓ | | ✓ | ✓ | ✓ | ✓ | ✓ | | | ✓ |
| Ecological conservation or restoration | 1/3 | 0/6 | | | | | | | ✓ | ✓ | | ✓ | | | |
| Engaging with/ tending to animals | 2/3 | 1/6 | | ✓ | | ✓ | | ✓ | ✓ | ✓ | | ✓ | | | ✓ |
| Wilderness activities | 2/3 | 0/6 | | | | | | ✓ | ✓ | ✓ | | ✓ | | | ✓ |
| Nature play | 2/3 | 2/6 | ✓ | | | | | ✓ | ✓ | ✓ | ✓ | ✓ | | | ✓ |
| Nature observation activities | 2/3 | 3/6 | ✓ | ✓ | | ✓ | | ✓ | ✓ | ✓ | ✓ | ✓ | | | ✓ |
| Mindfulness/ meditation/ relaxation | 1/3 | 2/6 | ✓ | | | | | ✓ | | ✓ | ✓ | ✓ | | | ✓ |
| Forest bathing | 0/3 | 0/6 | | | | | | | | | | | | | |
| Social activities | 3/3 | 3/6 | ✓ | | | | | ✓ | ✓ | ✓ | ✓ | ✓ | | | ✓ |
| No specific activity | 3/3 | 4/6 | ✓ | ✓ | ✓ | ✓ | ✓ | ✓ | ✓ | ✓ | ✓ | ✓ | ✓ | ✓ | ✓ |
| Other activities: community cultural development activities | 1/3 | 0/6 | | | | | | | ✓ | ✓ | | ✓ | | | |

Notes: Only headings relevant to our sample have been reported, e.g. no occupational therapist included in our study mainly worked with older adults. ^the main age groups worked with were not asked about in Stage 1.

**Table S2.3: Locations in which outdoor nature-based activities were recommended by the occupational therapists in our sample**

| | | | Age groups^ | | | Scope | | | | Sector | | Setting | | | |
|---|---|---|---|---|---|---|---|---|---|---|---|---|---|---|---|
| **Location** | **Number from Stage 1** | **Number from Stage 2** | **Children** | **Adolescents** | **Adults** | **Disability** | **Occupational health** | **Paediatrics** | **Community** | **Private** | **Public** | **Private practice** | **Disability services** | **Community pharmacy** | **Community setting** |
| Private garden | 3/3 | 2/6 | ✓ | | | | | ✓ | ✓ | ✓ | ✓ | ✓ | | | ✓ |
| Community/ school garden | 2/3 | 2/6 | ✓ | ✓ | | ✓ | | ✓ | ✓ | ✓ | ✓ | ✓ | | | ✓ |
| Farm | 2/3 | 0/6 | | | | | | ✓ | ✓ | ✓ | | ✓ | | | ✓ |
| Public park | 3/3 | 4/6 | ✓ | | ✓ | | ✓ | ✓ | ✓ | ✓ | ✓ | ✓ | ✓ | ✓ | ✓ |
| National park | 0/3 | 2/6 | ✓ | | | | | ✓ | | | ✓ | ✓ | | | |
| Forest | 1/3 | 1/6 | | ✓ | | ✓ | | | ✓ | ✓ | | ✓ | | | |
| Oval/ sports field | 2/3 | 0/6 | | | | | | ✓ | ✓ | ✓ | | ✓ | | | ✓ |
| On the lake | 1/3 | 0/6 | | | | | | | ✓ | ✓ | | ✓ | | | |
| Near the lake | 1/3 | 0/6 | | | | | | | ✓ | ✓ | | ✓ | | | |
| On the river | 2/3 | 0/6 | | | | | | ✓ | ✓ | ✓ | | ✓ | | | ✓ |
| Near the river | 2/3 | 0/6 | | | | | | ✓ | ✓ | ✓ | | ✓ | | | ✓ |
| On the ocean | 2/3 | 1/6 | | ✓ | | ✓ | | ✓ | ✓ | ✓ | | ✓ | | | ✓ |
| Near the ocean | 1/3 | 1/6 | | ✓ | | ✓ | | ✓ | | ✓ | | ✓ | | | ✓ |
| Other | 0/3 | 0/6 | | | | | | | | | | | | | |

Notes: Only headings relevant to our sample have been reported. ^the main age groups worked with were not asked about in Stage 1.

**Table S2.4: Recommendations for indoor nature-based activities from the occupational therapists in our sample**

| | | | Age groups^ | | Scope | | | Sector | | Setting | |
|---|---|---|---|---|---|---|---|---|---|---|---|
| Activity | Number from Stage 1 | Number from Stage 2 | Children | Adolescents | Disability | Paediatrics | Community | Private | Public | Private practice | Community setting |
| Any | 2/3 | 2/5 | ✓ | ✓ | ✓ | ✓ | ✓ | ✓ | ✓ | ✓ | ✓ |
| Listening to natural sounds | 2/3 | 1/5 | | ✓ | ✓ | ✓ | ✓ | ✓ | | ✓ | ✓ |
| Having indoor pot plants | 1/3 | 0/5 | | | | ✓ | | ✓ | | | ✓ |
| Pictures of natural scenes | 0/3 | 0/5 | | | | | | | | | |
| Natural scents | 1/3 | 2/5 | ✓ | ✓ | ✓ | ✓ | | ✓ | ✓ | ✓ | ✓ |
| Travelling in a car or on public transport through natural environments | 1/3 | 1/5 | | ✓ | ✓ | ✓ | | ✓ | | ✓ | ✓ |
| Views of nature outside the window | 1/3 | 0/5 | | | | ✓ | | ✓ | | | ✓ |
| Open windows for fresh air and the smells and sounds of nature | 2/3 | 1/5 | | ✓ | ✓ | ✓ | ✓ | ✓ | | ✓ | ✓ |
| Nature-based virtual reality | 1/3 | 0/5 | | | | | ✓ | ✓ | | ✓ | |
| Other: visits to public libraries, museums, galleries, archives | 1/3 | 0/5 | | | | | ✓ | ✓ | | ✓ | |

Notes: Only headings relevant to our sample have been reported. ^the main age groups worked with were not asked about in Stage 1.

### Direct engagement

**Table S2.5: Direct engagement in outdoor nature-based activities by occupational therapists in our sample with their patients/ clients**

| | | | Age groups^ | | Scope | | | Sector | | Setting | | |
|---|---|---|---|---|---|---|---|---|---|---|---|---|
| **Activity** | **Number from Stage 1** | **Number from Stage 2** | **Children** | **Adolescents** | **Disability** | **Paediatrics** | **Community** | **Private** | **Public** | **Private practice** | **Disability services** | **Community setting** |
| Any | 3/3 | 5/5 | ✓ | ✓ | ✓ | ✓ | ✓ | ✓ | ✓ | ✓ | ✓ | ✓ |
| Land-based gross motor activities | 2/3 | 4/5 | ✓ | ✓ | ✓ | ✓ | | ✓ | ✓ | ✓ | | ✓ |
| Land-based fine motor activities | 1/3 | 3/5 | ✓ | | | ✓ | ✓ | ✓ | ✓ | ✓ | ✓ | |
| Water-based physical activities | 1/3 | 3/5 | ✓ | | | ✓ | ✓ | ✓ | ✓ | ✓ | ✓ | |
| Active transport | 1/3 | 1/5 | | ✓ | ✓ | | ✓ | ✓ | | ✓ | | |
| Powered transport | 1/3 | 0/5 | | | | | ✓ | ✓ | | ✓ | | |
| Gardening | 1/3 | 2/5 | ✓ | ✓ | ✓ | ✓ | ✓ | ✓ | ✓ | ✓ | | |
| Ecological conservation or restoration | 1/3 | 0/5 | | | | | ✓ | ✓ | | ✓ | | |
| Engaging with/ tending to animals | 1/3 | 2/5 | ✓ | ✓ | ✓ | ✓ | ✓ | ✓ | ✓ | ✓ | | |
| Wilderness activities | 1/3 | 0/5 | | | | | ✓ | ✓ | | ✓ | | |
| Nature play | 2/3 | 4/5 | ✓ | ✓ | ✓ | ✓ | ✓ | ✓ | ✓ | ✓ | | ✓ |
| Nature observation activities | 2/3 | 2/5 | ✓ | | | ✓ | ✓ | ✓ | ✓ | ✓ | | ✓ |
| Mindfulness/ meditation/ relaxation | 1/3 | 1/5 | ✓ | | | ✓ | | | ✓ | ✓ | | |
| Forest bathing | 0/3 | 1/5 | ✓ | | | ✓ | | | ✓ | ✓ | | |
| Social activities | 3/3 | 2/5 | ✓ | | | ✓ | ✓ | ✓ | ✓ | ✓ | | ✓ |
| No specific activity | 3/3 | 2/5 | ✓ | | | ✓ | ✓ | ✓ | ✓ | ✓ | | ✓ |
| Other activities | 0/3 | 0/5 | | | | | | | | | | |

Notes: Only headings relevant to our sample have been reported. ^the main age groups worked with were not asked about in Stage 1.

**Table S2.6: Locations in which the occupational therapists in our sample engaged directly with their patients/ clients in outdoor nature-based activities**

| Location | Number from Stage 1 | Number from Stage 2 | Age groups^ | | Scope | | | Sector | | Setting | | |
|---|---|---|---|---|---|---|---|---|---|---|---|---|
| | | | Children | Adolescents | Disability | Paediatrics | Community | Private | Public | Private practice | Disability services | Community setting |
| Private garden | 3/3 | 2/5 | ✓ | | | ✓ | ✓ | ✓ | ✓ | ✓ | | ✓ |
| Community/ school garden | 3/3 | 3/5 | ✓ | ✓ | ✓ | ✓ | ✓ | ✓ | ✓ | ✓ | | ✓ |
| Farm | 1/3 | 0/5 | | | | | ✓ | ✓ | | ✓ | | |
| Public park | 3/3 | 4/5 | ✓ | | | ✓ | ✓ | ✓ | ✓ | ✓ | ✓ | ✓ |
| National park | 0/3 | 2/5 | ✓ | | | ✓ | | | ✓ | ✓ | | |
| Forest | 1/3 | 1/5 | ✓ | | | ✓ | ✓ | ✓ | ✓ | ✓ | | |
| Oval/ sports field | 2/3 | 2/5 | ✓ | ✓ | ✓ | ✓ | ✓ | ✓ | ✓ | ✓ | | ✓ |
| On the lake | 1/3 | 1/5 | ✓ | | | ✓ | ✓ | ✓ | ✓ | ✓ | | |
| Near the lake | 1/3 | 0/5 | | | | | ✓ | ✓ | | ✓ | | |
| On the river | 1/3 | 0/5 | | | | | ✓ | ✓ | | ✓ | | |
| Near the river | 1/3 | 0/5 | | | | | ✓ | ✓ | | ✓ | | |
| On the ocean | 1/3 | 0/5 | | | | | ✓ | ✓ | | ✓ | | |
| Near the ocean | 1/3 | 2/5 | ✓ | ✓ | ✓ | ✓ | ✓ | ✓ | ✓ | ✓ | | |
| Other | 0/3 | 0/5 | | | | | | | | | | |

Notes: Only headings relevant to our sample have been reported. ^the main age groups worked with were not asked about in Stage 1.

**Table S2.7: Direct engagement in indoor nature-based activities by occupational therapists in our sample with their patients/ clients**

| | | | Age groups^ | Scope | | | Sector | Setting | |
|---|---|---|---|---|---|---|---|---|---|
| Activity | Number from Stage 1 | Number from Stage 2 | Adolescents | Disability | Paediatrics | Community | Private | Private practice | Community setting |
| Any | 3/3 | 1/4 | ✓ | ✓ | ✓ | ✓ | ✓ | ✓ | ✓ |
| Listening to natural sounds | 3/3 | 0/4 | | | ✓ | ✓ | ✓ | ✓ | ✓ |
| Having indoor pot plants | 0/3 | 0/4 | | | | | | | |
| Pictures of natural scenes | 1/3 | 0/4 | | | ✓ | | ✓ | | ✓ |
| Natural scents | 0/3 | 0/4 | | | | | | | |
| Travelling in a car or on public transport through natural environments | 0/3 | 1/4 | ✓ | ✓ | | | ✓ | ✓ | |
| Views of nature outside the window | 1/3 | 0/4 | | | ✓ | | ✓ | | ✓ |
| Open windows for fresh air and the smells and sounds of nature | 3/3 | 1/4 | ✓ | ✓ | ✓ | ✓ | ✓ | ✓ | ✓ |
| Nature-based virtual reality | 1/3 | 0/4 | | | | ✓ | ✓ | ✓ | |
| Other: Community cultural development | 1/3 | 0/4 | | | | ✓ | ✓ | ✓ | |

Notes: Only headings relevant to our sample have been reported. ^the main age groups worked with were not asked about in Stage 1.

### Enhancing outdoor natural environments

**Table S2.8: Locations that occupational therapists in our sample have advocated for the addition of or improvement to**

| | | | Role | | | Age groups^ | | | Scope | | | | | Sector | | Setting | | |
|---|---|---|---|---|---|---|---|---|---|---|---|---|---|---|---|---|---|---|
| Location | Number from Stage 1 | Number from Stage 2 | Teacher/ education | Clinician | Discharge planning | Children | Adolescents | Adults | Disability | Acute care | Mental health | Paediatrics | Community | Private | Public | Private practice | Hospital | Disability services |
| Any | 1/4 | 6/6 | ✓ | ✓ | ✓ | ✓ | ✓ | ✓ | ✓ | ✓ | ✓ | ✓ | ✓ | ✓ | ✓ | ✓ | ✓ | ✓ |
| Gardens or other natural element in care settings | 0/4 | 1/6 | ✓ | | | | | ✓ | | | ✓ | | | ✓ | | | ✓ | |
| Nature elements inside care settings | 0/4 | 1/6 | ✓ | | | | | ✓ | | | ✓ | | | ✓ | | | ✓ | |
| Nature elements in non-care, public settings | 0/4 | 1/6 | | | ✓ | | | ✓ | | ✓ | | | | ✓ | | | ✓ | |
| Community gardens | 0/4 | 2/6 | ✓ | ✓ | | ✓ | ✓ | | ✓ | | | ✓ | | ✓ | ✓ | ✓ | | |
| School/ childcare gardens | 0/4 | 0/6 | | | | | | | | | | | | | | | | |
| Public parks/ gardens | 0/4 | 3/6 | ✓ | ✓ | | ✓ | | ✓ | | | ✓ | ✓ | | ✓ | ✓ | ✓ | ✓ | ✓ |
| Green corridors | 0/4 | 0/6 | | | | | | | | | | | | | | | | |
| Blue spaces/ water bodies | 0/4 | 0/6 | | | | | | | | | | | | | | | | |
| Other: National parks | 0/4 | 1/6 | ✓ | | | | | ✓ | | | ✓ | | | ✓ | | | ✓ | |

Notes: Only headings relevant to our sample have been reported. ^the main age groups worked with were not asked about in Stage 1.

**Table S2.9: Locations that occupational therapists in our sample have directly engaged in the addition of or improvement to**

| | | | Role | | Age groups^ | | Scope | | | Sector | | Setting |
|---|---|---|---|---|---|---|---|---|---|---|---|---|
| **Location** | **Number from Stage 1** | **Number from Stage 2** | **Teacher/ education** | **Clinician** | **Children** | **Adolescents** | **Disability** | **Paediatrics** | **Community** | **Private** | **Public** | **Private practice** |
| Any | 1/4 | 3/6 | ✓ | ✓ | ✓ | ✓ | ✓ | ✓ | ✓ | ✓ | ✓ | ✓ |
| Gardens or other natural element in care settings | 0/4 | 0/6 | | | | | | | | | | |
| Nature elements inside care settings | 0/4 | 0/6 | | | | | | | | | | |
| Nature elements in non-care, public settings | 0/4 | 1/6 | | ✓ | ✓ | | | ✓ | | | ✓ | ✓ |
| Community gardens | 0/4 | 2/6 | ✓ | ✓ | ✓ | ✓ | ✓ | ✓ | | ✓ | ✓ | ✓ |
| School/ childcare gardens | 0/4 | 0/6 | | | | | | | | | | |
| Public parks/ gardens | 0/4 | 1/6 | | ✓ | ✓ | | | ✓ | | | ✓ | ✓ |
| Green corridors | 0/4 | 0/6 | | | | | | | | | | |
| Blue spaces/ water bodies | 0/4 | 0/6 | | | | | | | | | | |
| Other | 0/4 | 0/6 | | | | | | | | | | |

Notes: Only headings relevant to our sample have been reported. ^the main age groups worked with were not asked about in Stage 1.

### Physiotherapy/ exercise physiology

**Table S2.10: Physiotherapy/ exercise physiology respondent demographics**

| | Stage 2 (n=11) |
|---|---|
| Physiotherapy | 10/11 |
| Gender | |
| Woman | 7/11 |
| Man | 4/11 |
| Age group | |
| 20-29 years | 2/11 |
| 30-39 years | 4/11 |
| 40-49 years | 3/11 |
| 50-59 years | 2/11 |
| 60+ years | |
| Trained in Australia (pre-registration qualification) | 11/11 |
| Years worked in discipline, median (interquartile range) | 14 (7-23) |
| Main patient age group | |
| Children | 2/10 |
| Adolescents | 0/10 |
| Adults | 8/10 |
| Older adults | 0/10 |
| Role | |
| Clinician | 7/10 |
| Research | 2/10 |
| Health screener | 1/10 |
| Scope/ area | |
| Disability | 2/10 |
| Musculoskeletal | 3/10 |
| Neurological | 2/10 |
| Sports | 1/10 |
| Paediatrics | 1/10 |
| Mental health (exercise physiology) | 1/10 |
| Stream/ area | |
| Community-based care | 4/10 |
| Rehabilitation | 3/10 |
| Acute care | 1/10 |
| Pain | 1/10 |
| Mental health (exercise physiology) | 1/10 |
| Setting | |
| Private practice | 5/10 |
| Home and community based service | 1/10 |
| Hospital | 1/10 |
| Other community health care service | 1/10 |
| Domiciliary service | 1/10 |
| Private company | 1/10 |
| Private sector | 8/10 |
| Sees patients/ clients | 9/11 |
| Telehealth | 8/9 |
| Home visits only | 2/9 |
| Main practice in a major city | 3/6 |
| Hours worked in main practice setting, median (interquartile range) | 20 (12-32) |

### Recommendations

**Table S2.11: Recommendations for outdoor nature-based activities from the physiotherapists/ exercise physiologist in our sample**

| | | | Age groups | | Scope/ area | | | | | Stream | | | | | Sector | | Setting | | | | | |
|---|---|---|---|---|---|---|---|---|---|---|---|---|---|---|---|---|---|---|---|---|---|---|
| **Activity** | **Exercise physiologist** | **Physiotherapist** | **Children** | **Adults** | **Disability** | **Musculoskeletal** | **Neurological** | **Sports** | **Mental health** | **Community-based care** | **Rehabilitation** | **Acute care** | **Pain** | **Mental health** | **Private** | **Public** | **Private practice** | **Home and community-based service** | **Hospital** | **Other community health care service** | **Domiciliary service** | **Private company** |
| Any | 1/1 | 7/8 | ✓ | ✓ | ✓ | ✓ | ✓ | ✓ | ✓ | ✓ | ✓ | ✓ | ✓ | ✓ | ✓ | ✓ | ✓ | ✓ | ✓ | ✓ | ✓ | ✓ |
| Land-based gross motor activities | 1/1 | 7/8 | ✓ | ✓ | ✓ | ✓ | ✓ | ✓ | ✓ | ✓ | ✓ | ✓ | ✓ | ✓ | ✓ | ✓ | ✓ | ✓ | ✓ | ✓ | ✓ | ✓ |
| Land-based fine motor activities | 1/1 | 4/8 | ✓ | ✓ | ✓ | ✓ | ✓ | ✓ | ✓ | ✓ | ✓ | ✓ | ✓ | ✓ | ✓ | ✓ | ✓ | ✓ | | ✓ | ✓ | ✓ |
| Water-based physical activities | 1/1 | 6/8 | ✓ | ✓ | ✓ | ✓ | ✓ | ✓ | ✓ | ✓ | ✓ | ✓ | ✓ | ✓ | ✓ | ✓ | ✓ | ✓ | | | ✓ | ✓ |
| Active transport | 1/1 | 3/8 | ✓ | ✓ | ✓ | ✓ | | | ✓ | ✓ | | | ✓ | ✓ | ✓ | ✓ | ✓ | ✓ | | ✓ | | ✓ |
| Powered transport | 1/1 | 3/8 | ✓ | ✓ | ✓ | | ✓ | | ✓ | ✓ | | | | ✓ | ✓ | ✓ | | ✓ | | ✓ | ✓ | ✓ |
| Gardening | 1/1 | 6/8 | ✓ | ✓ | ✓ | ✓ | ✓ | | ✓ | ✓ | ✓ | ✓ | ✓ | ✓ | ✓ | ✓ | ✓ | ✓ | | ✓ | ✓ | ✓ |
| Ecological conservation or restoration | 0/1 | 1/8 | | ✓ | | ✓ | | | | | | | ✓ | | ✓ | | ✓ | | | | | |
| Engaging with/ tending to animals | 1/1 | 2/8 | | ✓ | | ✓ | | ✓ | ✓ | | ✓ | | ✓ | ✓ | ✓ | ✓ | ✓ | | | | | ✓ |
| Wilderness activities | 1/1 | 4/8 | ✓ | ✓ | ✓ | ✓ | | ✓ | ✓ | ✓ | ✓ | | ✓ | ✓ | ✓ | ✓ | ✓ | ✓ | | | | ✓ |
| Nature play | 1/1 | 3/8 | ✓ | ✓ | ✓ | ✓ | | | ✓ | ✓ | | ✓ | ✓ | ✓ | ✓ | ✓ | ✓ | ✓ | | | | ✓ |
| Nature observation activities | 1/1 | 2/8 | | ✓ | | ✓ | ✓ | | ✓ | ✓ | | | ✓ | ✓ | ✓ | ✓ | ✓ | | | | ✓ | ✓ |
| Mindfulness/ meditation/ relaxation | 1/1 | 7/8 | ✓ | ✓ | ✓ | ✓ | ✓ | ✓ | ✓ | ✓ | ✓ | ✓ | ✓ | ✓ | ✓ | ✓ | ✓ | ✓ | ✓ | ✓ | ✓ | ✓ |
| Forest bathing | 1/1 | 1/8 | | ✓ | | ✓ | | | ✓ | | | ✓ | | ✓ | ✓ | ✓ | ✓ | | | | | ✓ |
| Social activities | 1/1 | 4/8 | ✓ | ✓ | ✓ | ✓ | | | ✓ | ✓ | ✓ | | ✓ | ✓ | ✓ | ✓ | ✓ | ✓ | | ✓ | | ✓ |
| No specific activity | 1/1 | 6/8 | ✓ | ✓ | ✓ | ✓ | ✓ | | ✓ | ✓ | ✓ | ✓ | ✓ | ✓ | ✓ | ✓ | ✓ | ✓ | | ✓ | ✓ | ✓ |
| Other activities | 0/1 | 0/8 | | | | | | | | | | | | | | | | | | | | |

Notes: Only headings relevant to our sample have been reported

**Table S2.12: Locations in which outdoor nature-based activities were recommended by the physiotherapists/ exercise physiologist in our sample**

| | | | Age groups | | Scope/ area | | | | | Stream | | | | | Sector | | Setting | | | | |
|---|---|---|---|---|---|---|---|---|---|---|---|---|---|---|---|---|---|---|---|---|---|
| Location | Exercise physiologist | Physiotherapist | Children | Adults | Disability | Musculoskeletal | Neurological | Sports | Mental health | Community-based care | Rehabilitation | Acute care | Pain | Mental health | Private | Public | Private practice | Home and community-based service | Other community health care service | Domiciliary service | Private company |
| Private garden | 1/1 | 6/7 | ✓ | ✓ | ✓ | ✓ | ✓ | | ✓ | ✓ | ✓ | ✓ | ✓ | ✓ | ✓ | ✓ | ✓ | ✓ | ✓ | ✓ | ✓ |
| Community/ school garden | 0/1 | 2/7 | ✓ | ✓ | ✓ | ✓ | | | | ✓ | ✓ | | | | ✓ | | ✓ | ✓ | | | |
| Farm | 1/1 | 2/7 | ✓ | ✓ | ✓ | | | ✓ | ✓ | ✓ | ✓ | | | ✓ | ✓ | ✓ | ✓ | ✓ | | | ✓ |
| Public park | 1/1 | 5/7 | ✓ | ✓ | ✓ | ✓ | ✓ | | ✓ | ✓ | | ✓ | ✓ | ✓ | ✓ | ✓ | ✓ | ✓ | ✓ | ✓ | ✓ |
| National park | 1/1 | 6/7 | ✓ | ✓ | ✓ | ✓ | ✓ | | ✓ | ✓ | ✓ | ✓ | ✓ | ✓ | ✓ | ✓ | ✓ | ✓ | ✓ | ✓ | ✓ |
| Forest | 1/1 | 2/7 | | ✓ | | ✓ | | | ✓ | | ✓ | ✓ | | ✓ | ✓ | ✓ | ✓ | | | | ✓ |
| Oval/ sports field | 1/1 | 6/7 | ✓ | ✓ | ✓ | ✓ | ✓ | | ✓ | ✓ | ✓ | ✓ | ✓ | ✓ | ✓ | ✓ | ✓ | ✓ | ✓ | ✓ | ✓ |
| On the lake | 1/1 | 1/7 | | ✓ | | ✓ | | | ✓ | | | ✓ | | ✓ | ✓ | ✓ | ✓ | | | | ✓ |
| Near the lake | 1/1 | 2/7 | | ✓ | | ✓ | | | ✓ | | ✓ | ✓ | | ✓ | ✓ | ✓ | ✓ | | | | ✓ |
| On the river | 1/1 | 2/7 | | ✓ | ✓ | ✓ | | | ✓ | ✓ | | ✓ | | ✓ | ✓ | ✓ | ✓ | | ✓ | | ✓ |
| Near the river | 1/1 | 4/7 | | ✓ | ✓ | ✓ | ✓ | | ✓ | ✓ | ✓ | ✓ | | ✓ | ✓ | ✓ | ✓ | ✓ | ✓ | ✓ | ✓ |
| On the ocean | 1/1 | 3/7 | ✓ | ✓ | ✓ | ✓ | | | ✓ | ✓ | | ✓ | ✓ | ✓ | ✓ | ✓ | ✓ | ✓ | | | ✓ |
| Near the ocean | 1/1 | 4/7 | ✓ | ✓ | ✓ | ✓ | | | ✓ | ✓ | ✓ | ✓ | | ✓ | ✓ | ✓ | ✓ | ✓ | ✓ | | ✓ |
| Other | 0/1 | 0/7 | | | | | | | | | | | | | | | | | | | |

Notes: Only headings relevant to our sample have been reported

**Table S2.13: Recommendations for indoor nature-based activities from the physiotherapists/ exercise physiologist in our sample**

| Activity | Exercise physiologist | Physiotherapist | Age groups | Scope/ area | | | | Stream | | | | Sector | | Setting | | | | |
|---|---|---|---|---|---|---|---|---|---|---|---|---|---|---|---|---|---|---|
| | | | Adults | Disability | Musculoskeletal | Neurological | Mental health | Community-based care | Acute care | Pain | Mental health | Private | Public | Private practice | Home and community-based service | Other community health care service | Domiciliary service | Private company |
| Any | 1/1 | 4/7 | ✓ | ✓ | ✓ | ✓ | ✓ | ✓ | ✓ | ✓ | ✓ | ✓ | ✓ | ✓ | | ✓ | ✓ | ✓ |
| Listening to natural sounds | 1/1 | 2/7 | ✓ | | ✓ | ✓ | ✓ | ✓ | ✓ | | ✓ | ✓ | ✓ | ✓ | | | ✓ | ✓ |
| Having indoor pot plants | 1/1 | 1/7 | ✓ | | ✓ | | ✓ | | | ✓ | ✓ | ✓ | ✓ | | ✓ | | | ✓ |
| Pictures of natural scenes | 1/1 | 0/7 | ✓ | | | | ✓ | | | | ✓ | | ✓ | | | | | ✓ |
| Natural scents | 1/1 | 1/7 | ✓ | | | ✓ | ✓ | ✓ | | | ✓ | ✓ | ✓ | | | | ✓ | ✓ |
| Travelling in a car or on public transport through natural environments | 0/1 | 0/7 | | | | | | | | | | | | | | | | |
| Views of nature outside the window | 1/1 | 2/7 | ✓ | | ✓ | ✓ | ✓ | ✓ | | ✓ | ✓ | ✓ | ✓ | ✓ | | | ✓ | ✓ |
| Open windows for fresh air and the smells and sounds of nature | 0/1 | 3/7 | ✓ | ✓ | ✓ | ✓ | | ✓ | | ✓ | | ✓ | | ✓ | | ✓ | ✓ | |
| Nature-based virtual reality | 0/1 | 1/7 | ✓ | | | ✓ | | ✓ | | | | ✓ | | | | | ✓ | |
| Other: watching video of nature | 1/1 | 0/7 | ✓ | | | | ✓ | | | | ✓ | | ✓ | | | | | ✓ |

### Direct engagement

**Table S2.14: Direct engagement in outdoor nature-based activities by physiotherapists/ exercise physiologist in our sample with their patients/ clients**

| | | | Age groups | | Scope/ area | | | | | Stream | | | | Sector | | Setting | | | | |
|---|---|---|---|---|---|---|---|---|---|---|---|---|---|---|---|---|---|---|---|---|
| **Activity** | **Exercise physiologist** | **Physiotherapist** | **Children** | **Adults** | **Disability** | **Musculoskeletal** | **Neurological** | **Sports** | **Mental health** | **Community-based care** | **Rehabilitation** | **Pain** | **Mental health** | **Private** | **Public** | **Private practice** | **Home and community-based service** | **Other community health care service** | **Domiciliary service** | **Private company** |
| Any | 1/1 | 6/8 | ✓ | ✓ | ✓ | ✓ | ✓ | ✓ | ✓ | ✓ | ✓ | ✓ | ✓ | ✓ | ✓ | ✓ | ✓ | ✓ | ✓ | ✓ |
| Land-based gross motor activities | 1/1 | 6/8 | ✓ | ✓ | ✓ | ✓ | ✓ | ✓ | ✓ | ✓ | ✓ | ✓ | ✓ | ✓ | ✓ | ✓ | ✓ | ✓ | ✓ | ✓ |
| Land-based fine motor activities | 1/1 | 1/8 | | ✓ | ✓ | | | | ✓ | ✓ | | | ✓ | ✓ | ✓ | | | ✓ | | ✓ |
| Water-based physical activities | 0/1 | 3/8 | ✓ | ✓ | ✓ | | ✓ | | | ✓ | | | | ✓ | | | ✓ | ✓ | ✓ | |
| Active transport | 0/1 | 2/8 | ✓ | ✓ | ✓ | | | | | ✓ | | | | ✓ | | | ✓ | ✓ | | |
| Powered transport | 0/1 | 3/8 | ✓ | ✓ | ✓ | | ✓ | | | ✓ | | | | ✓ | | | ✓ | ✓ | ✓ | |
| Gardening | 1/1 | 2/8 | | ✓ | ✓ | | ✓ | | ✓ | ✓ | | | ✓ | ✓ | | | | ✓ | ✓ | ✓ |
| Ecological conservation or restoration | 0/1 | 0/8 | | | | | | | | | | | | | | | | | | |
| Engaging with/ tending to animals | 0/1 | 2/8 | | ✓ | ✓ | | | ✓ | | ✓ | ✓ | | | ✓ | | ✓ | | ✓ | | |
| Wilderness activities | 1/1 | 0/8 | | ✓ | | | | | ✓ | | | | ✓ | | ✓ | | | | | ✓ |
| Nature play | 0/1 | 1/8 | ✓ | | ✓ | | | | | ✓ | | | | ✓ | | | ✓ | | | |
| Nature observation activities | 1/1 | 1/8 | | ✓ | ✓ | | | | ✓ | ✓ | | | ✓ | ✓ | ✓ | | | ✓ | | ✓ |
| Mindfulness/ meditation/ relaxation | 1/1 | 1/8 | ✓ | ✓ | | | ✓ | | ✓ | ✓ | | | ✓ | ✓ | ✓ | | | | ✓ | ✓ |
| Forest bathing | 0/1 | 0/8 | | | | | | | | | | | | | | | | | | |
| Social activities | 0/1 | 2/8 | ✓ | ✓ | ✓ | | | | | ✓ | | | | ✓ | | | ✓ | ✓ | | |
| No specific activity | 1/1 | 4/8 | ✓ | ✓ | ✓ | ✓ | ✓ | | ✓ | ✓ | | ✓ | ✓ | ✓ | ✓ | ✓ | ✓ | ✓ | ✓ | ✓ |
| Other activities | 0/1 | 0/8 | | | | | | | | | | | | | | | | | | |

Notes: Only headings relevant to our sample have been reported.

**Table S2.15: Locations in which the physiotherapists/ exercise physiologist in our sample engaged directly with their patients/ clients in outdoor nature-based activities**

| | | | Age groups | | Scope/ area | | | | | Stream | | | | Sector | | Setting | | | | |
|---|---|---|---|---|---|---|---|---|---|---|---|---|---|---|---|---|---|---|---|---|
| **Locations** | **Exercise physiologist** | **Physiotherapist** | **Children** | **Adults** | **Disability** | **Musculoskeletal** | **Neurological** | **Sports** | **Mental health** | **Community-based care** | **Rehabilitation** | **Pain** | **Mental health** | **Private** | **Public** | **Private practice** | **Home and community-based service** | **Other community health care service** | **Domiciliary service** | **Private company** |
| Private garden | 1/1 | 4/8 | ✓ | ✓ | ✓ | ✓ | ✓ | | ✓ | ✓ | | ✓ | ✓ | ✓ | ✓ | ✓ | ✓ | ✓ | ✓ | ✓ |
| Community/ school garden | 0/1 | 1/8 | ✓ | | ✓ | | | | | ✓ | | | | ✓ | | | ✓ | | | |
| Farm | 1/1 | 1/8 | | ✓ | | | | ✓ | ✓ | | ✓ | | ✓ | ✓ | ✓ | ✓ | | | | ✓ |
| Public park | 1/1 | 4/8 | ✓ | ✓ | ✓ | ✓ | ✓ | | ✓ | ✓ | ✓ | | ✓ | ✓ | ✓ | ✓ | ✓ | ✓ | ✓ | ✓ |
| National park | 0/1 | 0/8 | | | | | | | | | | | | | | | | | | |
| Forest | 0/1 | 0/8 | | | | | | | | | | | | | | | | | | |
| Oval/ sports field | 1/1 | 3/8 | ✓ | ✓ | ✓ | | ✓ | | ✓ | ✓ | | | ✓ | ✓ | ✓ | | ✓ | ✓ | ✓ | ✓ |
| On the lake | 0/1 | 0/8 | | | | | | | | | | | | | | | | | | |
| Near the lake | 0/1 | 0/8 | | | | | | | | | | | | | | | | | | |
| On the river | 0/1 | 0/8 | | | | | | | | | | | | | | | | | | |
| Near the river | 1/1 | 2/8 | | ✓ | ✓ | | ✓ | | ✓ | ✓ | | | ✓ | ✓ | ✓ | | | ✓ | ✓ | ✓ |
| On the ocean | 0/1 | 0/8 | | | | | | | | | | | | | | | | | | |
| Near the ocean | 0/1 | 0/8 | | | | | | | | | | | | | | | | | | |
| Other | 0/1 | 0/8 | | | | | | | | | | | | | | | | | | |

**Table S2.16: Direct engagement in indoor nature-based activities by physiotherapists/ exercise physiologist in our sample with their patients/ clients**

| Activity | Exercise physiologist | Physiotherapist | Age groups | Scope/ area | | | | Stream | | | Sector | | Setting | | | | |
|---|---|---|---|---|---|---|---|---|---|---|---|---|---|---|---|---|---|
| | | | Adults | Disability | Musculoskeletal | Neurological | Mental health | Community-based care | Pain | Mental health | Private | Public | Private practice | Home and community-based service | Other community health care service | Domiciliary service | Private company |
| Any | 1/1 | 3/7 | ✓ | ✓ | ✓ | ✓ | ✓ | ✓ | ✓ | ✓ | ✓ | ✓ | ✓ | | ✓ | ✓ | ✓ |
| Listening to natural sounds | 1/1 | 2/7 | ✓ | | ✓ | ✓ | ✓ | ✓ | ✓ | ✓ | ✓ | ✓ | ✓ | | | ✓ | ✓ |
| Having indoor pot plants | 1/1 | 2/7 | ✓ | | ✓ | ✓ | ✓ | ✓ | ✓ | ✓ | ✓ | ✓ | ✓ | | | ✓ | ✓ |
| Pictures of natural scenes | 1/1 | 1/7 | ✓ | | ✓ | | ✓ | | ✓ | ✓ | ✓ | ✓ | ✓ | | | | ✓ |
| Natural scents | 0/1 | 2/7 | ✓ | | ✓ | ✓ | | ✓ | ✓ | | ✓ | | ✓ | | | ✓ | |
| Travelling in a car or on public transport through natural environments | 0/1 | 0/7 | | | | | | | | | | | | | | | |
| Views of nature outside the window | 1/1 | 2/7 | ✓ | | ✓ | ✓ | ✓ | ✓ | ✓ | ✓ | ✓ | ✓ | ✓ | | | ✓ | ✓ |
| Open windows for fresh air and the smells and sounds of nature | 1/1 | 3/7 | ✓ | ✓ | ✓ | ✓ | ✓ | ✓ | ✓ | ✓ | ✓ | ✓ | | ✓ | ✓ | ✓ | ✓ |
| Nature-based virtual reality | 0/1 | 1/7 | ✓ | | | ✓ | | ✓ | | | ✓ | | | | | ✓ | |
| Other: watching video of nature | 1/1 | 0/7 | ✓ | | | | ✓ | | | ✓ | | ✓ | | | | | ✓ |

Notes: Only headings relevant to our sample have been reported

### Enhancing outdoor natural environments

**Table S2.17: Locations that physiotherapists/ exercise physiologist in our sample have advocated for the addition of or improvement to**

| | | | Age groups | | Scope/ area | | | | | | Stream | | | | | Sector | | Setting | | | |
|---|---|---|---|---|---|---|---|---|---|---|---|---|---|---|---|---|---|---|---|---|---|
| **Locations** | **Exercise physiologist** | **Physiotherapist** | **Children** | **Adults** | **Disability** | **Musculoskeletal** | **Neurological** | **Sports** | **Paediatrics** | **Mental health** | **Community-based care** | **Rehabilitation** | **Acute care** | **Pain** | **Mental health** | **Private** | **Public** | **Private practice** | **Hospital** | **Domiciliary service** | **Private company** |
| Any | 1/1 | 6/9 | ✓ | ✓ | | ✓ | ✓ | ✓ | ✓ | ✓ | ✓ | ✓ | | ✓ | ✓ | ✓ | ✓ | ✓ | ✓ | ✓ | ✓ |
| Gardens or other natural element in care settings | 1/1 | 1/9 | ✓ | ✓ | | | | | ✓ | ✓ | | ✓ | | | ✓ | | ✓ | | ✓ | | ✓ |
| Nature elements inside care settings | 1/1 | 0/9 | | ✓ | | | | | | ✓ | | | | | ✓ | | ✓ | | | | ✓ |
| Nature elements in non-care, public settings | 0/1 | 2/9 | ✓ | ✓ | | | ✓ | | ✓ | | ✓ | ✓ | | | | ✓ | ✓ | | ✓ | ✓ | |
| Community gardens | 0/1 | 4/9 | ✓ | ✓ | | ✓ | ✓ | | ✓ | | ✓ | ✓ | | | | ✓ | ✓ | ✓ | ✓ | ✓ | |
| School/ childcare gardens | 0/1 | 2/9 | ✓ | ✓ | | | ✓ | | ✓ | | ✓ | ✓ | | | | ✓ | ✓ | | ✓ | ✓ | |
| Public parks/ gardens | 0/1 | 3/9 | ✓ | ✓ | | | ✓ | ✓ | ✓ | | ✓ | ✓ | | | | ✓ | ✓ | ✓ | ✓ | ✓ | |
| Green corridors | 0/1 | 3/9 | ✓ | ✓ | | | ✓ | ✓ | ✓ | | ✓ | ✓ | | | | ✓ | ✓ | ✓ | ✓ | ✓ | |
| Blue spaces/ water bodies | 0/1 | 1/9 | ✓ | | | | | | ✓ | | | ✓ | | | | | ✓ | | ✓ | | |
| Other: 'Bush' | 0/1 | 1/9 | | ✓ | | ✓ | | | | | | | | ✓ | | ✓ | | ✓ | | | |

Notes: Only headings relevant to our sample have been reported

**Table S2.18: Locations that physiotherapists/ exercise physiologist in our sample have directly engaged in the addition of or improvement to**

| | | Age groups | | Scope/ area | | | | Stream | | | | Sector | | Setting | | | | |
|---|---|---|---|---|---|---|---|---|---|---|---|---|---|---|---|---|---|---|
| **Locations** | **Physiotherapist** | **Children** | **Adults** | **Disability** | **Musculoskeletal** | **Neurological** | **Paediatrics** | **Community-based care** | **Rehabilitation** | **Acute care** | **Pain** | **Private** | **Public** | **Private practice** | **Home and community-based service** | **Hospital** | **Other community health care service** | **Domiciliary service** |
| Any | 5/8 | ✓ | ✓ | ✓ | ✓ | ✓ | ✓ | ✓ | ✓ | ✓ | ✓ | ✓ | ✓ | ✓ | ✓ | ✓ | ✓ | ✓ |
| Gardens or other natural element in care settings | 0/8 | | | | | | | | | | | | | | | | | |
| Nature elements inside care settings | 0/8 | | | | | | | | | | | | | | | | | |
| Nature elements in non-care, public settings | 0/8 | | | | | | | | | | | | | | | | | |
| Community gardens | 3/8 | | ✓ | ✓ | ✓ | ✓ | | ✓ | | ✓ | | ✓ | | ✓ | | | ✓ | ✓ |
| School/ childcare gardens | 1/8 | | ✓ | | | ✓ | | ✓ | | | | ✓ | | | | | | ✓ |
| Public parks/ gardens | 2/8 | | ✓ | | ✓ | ✓ | | ✓ | | ✓ | | ✓ | | ✓ | | | | ✓ |
| Green corridors | 3/8 | ✓ | ✓ | | ✓ | ✓ | ✓ | ✓ | ✓ | ✓ | | ✓ | ✓ | ✓ | | ✓ | | ✓ |
| Blue spaces/ water bodies | 3/8 | ✓ | ✓ | ✓ | ✓ | | ✓ | ✓ | ✓ | ✓ | | ✓ | ✓ | ✓ | | ✓ | ✓ | |
| Other: 'Bush' | 1/8 | | ✓ | | ✓ | | | | | | ✓ | ✓ | | ✓ | | | | |

Notes: Only headings relevant to our sample have been reported

### Psychology

**Table S2.19: Psychology respondent demographics**

| | Stage 2 (n=22) |
|---|---|
| Gender | |
| Woman | 19/22 |
| Man | 3/22 |
| Age group | |
| 20-29 years | 1/22 |
| 30-39 years | 7/22 |
| 40-49 years | 3/22 |
| 50-59 years | 5/22 |
| 60+ years | 6/22 |
| Trained in Australia (pre-registration qualification) | 19/22 |
| Years worked in discipline, median (interquartile range) | 15 (7-29) |
| Main patient age group | |
| Children | 1/22 |
| Adolescents | 3/22 |
| Adults | 17/22 |
| Older adults | 1/22 |
| Role | |
| Teacher/ educator | 1/22 |
| Clinician | 20/22 |
| Administration | 1/22 |
| Area of endorsement | |
| None | 10/22 |
| Clinical psychology | 10/22 |
| Clinical psychology & counselling psychology | 1/22 |
| Health psychology | 1/22 |
| Area of practice | |
| Psychology management/ administration | 1/22 |
| Mental health intervention | 10/22 |
| Mental health rehabilitation | 1/22 |
| Counselling | 6/22 |
| Personal development/ coaching | 1/22 |
| Organisational practice | 1/22 |
| Teaching/ supervision | 1/22 |
| Perinatal | 1/22 |
| Setting | |
| Private practice | 17/21 |
| Government department/ agency | 2/21 |
| Community mental health service | 2/21 |
| Private sector | 16/22 |
| Sees patients/ clients | 21/22 |
| Telehealth | 20/21 |
| Home visits only | 1/21 |
| Main practice in a major city | 16/20 |
| Hours worked in main practice setting, median (interquartile range) | 20 (15-32) |

### Recommendations

**Table S2.20: Recommendations for outdoor nature-based activities from the psychologists in our sample**

| | | Age groups | | | | Endorsement area | | | | Practice area | | | | | | Sector | | Setting | | |
|---|---|---|---|---|---|---|---|---|---|---|---|---|---|---|---|---|---|---|---|---|
| **Activity** | **Psychologists** | **Children** | **Adolescents** | **Adults** | **Older adults** | **None** | **Clinical psychology** | **Clinical psychology & counselling psychology** | **Health psychology** | **Psychology management/ administration** | **Mental health intervention** | **Counselling** | **Personal development/ coaching** | **Organisational practice** | **Perinatal** | **Private** | **Public** | **Private practice** | **Government department or agency** | **Community mental health service** |
| Any | 19/19 | ✓ | ✓ | ✓ | ✓ | ✓ | ✓ | ✓ | ✓ | ✓ | ✓ | ✓ | ✓ | ✓ | ✓ | ✓ | ✓ | ✓ | ✓ | ✓ |
| Land-based gross motor activities | 9/19 | | ✓ | ✓ | ✓ | ✓ | ✓ | | ✓ | | ✓ | ✓ | | ✓ | ✓ | ✓ | ✓ | ✓ | ✓ | |
| Land-based fine motor activities | 8/19 | ✓ | ✓ | ✓ | ✓ | ✓ | ✓ | | ✓ | | ✓ | ✓ | | | ✓ | ✓ | ✓ | ✓ | | |
| Water-based physical activities | 7/19 | | ✓ | ✓ | ✓ | ✓ | ✓ | | ✓ | | ✓ | ✓ | | | ✓ | ✓ | | ✓ | | |
| Active transport | 6/19 | ✓ | ✓ | ✓ | ✓ | ✓ | ✓ | | ✓ | | ✓ | ✓ | | | ✓ | ✓ | ✓ | ✓ | | |
| Powered transport | 3/19 | | | ✓ | ✓ | ✓ | ✓ | | | | ✓ | | | | | ✓ | | ✓ | | |
| Gardening | 14/19 | | ✓ | ✓ | ✓ | ✓ | ✓ | | ✓ | ✓ | ✓ | ✓ | ✓ | ✓ | ✓ | ✓ | ✓ | ✓ | ✓ | ✓ |
| Ecological conservation or restoration | 7/19 | | | ✓ | ✓ | ✓ | ✓ | | ✓ | | ✓ | | ✓ | ✓ | ✓ | ✓ | ✓ | ✓ | ✓ | |
| Engaging with/ tending to animals | 12/19 | ✓ | ✓ | ✓ | ✓ | ✓ | ✓ | | ✓ | | ✓ | ✓ | | ✓ | ✓ | ✓ | ✓ | ✓ | ✓ | ✓ |
| Wilderness activities | 9/19 | | | ✓ | | ✓ | ✓ | | ✓ | ✓ | | ✓ | ✓ | ✓ | ✓ | ✓ | ✓ | ✓ | ✓ | |
| Nature play | 10/19 | ✓ | ✓ | ✓ | ✓ | ✓ | ✓ | | ✓ | | ✓ | ✓ | ✓ | ✓ | ✓ | ✓ | ✓ | ✓ | ✓ | |
| Nature observation activities | 12/19 | | ✓ | ✓ | ✓ | ✓ | ✓ | | ✓ | | ✓ | ✓ | ✓ | | ✓ | ✓ | ✓ | ✓ | | ✓ |
| Mindfulness/ meditation/ relaxation | 19/19 | ✓ | ✓ | ✓ | ✓ | ✓ | ✓ | ✓ | ✓ | ✓ | ✓ | ✓ | ✓ | ✓ | ✓ | ✓ | ✓ | ✓ | ✓ | ✓ |
| Forest bathing | 7/19 | | | ✓ | ✓ | ✓ | ✓ | | | | ✓ | ✓ | ✓ | | | ✓ | | ✓ | | |
| Social activities | 11/19 | ✓ | ✓ | ✓ | ✓ | ✓ | ✓ | ✓ | ✓ | | ✓ | ✓ | | | ✓ | ✓ | ✓ | ✓ | | ✓ |
| No specific activity | 18/19 | ✓ | ✓ | ✓ | ✓ | ✓ | ✓ | ✓ | ✓ | ✓ | ✓ | ✓ | ✓ | ✓ | ✓ | ✓ | ✓ | ✓ | ✓ | ✓ |
| Other: reading | 1/19 | | | ✓ | | ✓ | | | | | | ✓ | | | | ✓ | | ✓ | | |
| Other: art | | | | ✓ | | | | | ✓ | | | | | | ✓ | ✓ | | ✓ | | |

Notes: Only headings relevant to our sample have been reported

**Table S2.21: Locations in which outdoor nature-based activities were recommended by the psychologists in our sample**

| | | Age groups | | | | Endorsement area | | | | Practice area | | | | | | Sector | | Setting | | |
|---|---|---|---|---|---|---|---|---|---|---|---|---|---|---|---|---|---|---|---|---|
| **Location** | **Psychologists** | **Children** | **Adolescents** | **Adults** | **Older adults** | **None** | **Clinical psychology** | **Clinical psychology & counselling psychology** | **Health psychology** | **Psychology management/ administration** | **Mental health intervention** | **Counselling** | **Personal development/ coaching** | **Organisational practice** | **Perinatal** | **Private** | **Public** | **Private practice** | **Government department or agency** | **Community mental health service** |
| Private garden | 17/19 | ✓ | ✓ | ✓ | ✓ | ✓ | ✓ | ✓ | ✓ | ✓ | ✓ | ✓ | ✓ | | ✓ | ✓ | ✓ | ✓ | | ✓ |
| Community/ school garden | 8/19 | ✓ | ✓ | ✓ | ✓ | ✓ | ✓ | | ✓ | | ✓ | ✓ | | | ✓ | ✓ | ✓ | ✓ | | |
| Farm | 6/19 | ✓ | ✓ | ✓ | ✓ | ✓ | ✓ | | ✓ | | ✓ | ✓ | | | ✓ | ✓ | ✓ | ✓ | | |
| Public park | 15/19 | | ✓ | ✓ | ✓ | ✓ | ✓ | ✓ | ✓ | ✓ | ✓ | ✓ | ✓ | ✓ | ✓ | ✓ | ✓ | ✓ | ✓ | ✓ |
| National park | 12/19 | | ✓ | ✓ | ✓ | ✓ | ✓ | ✓ | ✓ | | ✓ | ✓ | ✓ | ✓ | ✓ | ✓ | ✓ | ✓ | ✓ | |
| Forest | 11/19 | ✓ | ✓ | ✓ | ✓ | ✓ | ✓ | | ✓ | | ✓ | ✓ | ✓ | ✓ | ✓ | ✓ | ✓ | ✓ | ✓ | |
| Oval/ sports field | 5/19 | | ✓ | ✓ | | ✓ | ✓ | | ✓ | | ✓ | ✓ | | ✓ | ✓ | ✓ | ✓ | ✓ | ✓ | |
| On the lake | 3/19 | | | ✓ | | | ✓ | | ✓ | | ✓ | | | | ✓ | ✓ | | ✓ | | |
| Near the lake | 7/19 | | ✓ | ✓ | | ✓ | ✓ | | ✓ | | ✓ | ✓ | | ✓ | ✓ | ✓ | ✓ | ✓ | ✓ | |
| On the river | 5/19 | | ✓ | ✓ | | ✓ | ✓ | | ✓ | | ✓ | ✓ | | | ✓ | ✓ | | ✓ | | |
| Near the river | 10/19 | | ✓ | ✓ | | ✓ | ✓ | ✓ | ✓ | | ✓ | ✓ | ✓ | | ✓ | ✓ | | ✓ | | |
| On the ocean | 7/19 | | ✓ | ✓ | ✓ | ✓ | ✓ | | ✓ | | ✓ | ✓ | ✓ | | ✓ | ✓ | ✓ | ✓ | | |
| Near the ocean | 14/19 | ✓ | ✓ | ✓ | | ✓ | ✓ | ✓ | ✓ | | ✓ | ✓ | ✓ | | ✓ | ✓ | ✓ | ✓ | | |
| Other | 0/19 | | | | | | | | | | | | | | | | | | | |

Notes: Only headings relevant to our sample have been reported

**Table S2.22: Recommendations for indoor nature-based activities from the psychologists in our sample**

| Activity | Psychologists | Age groups: Children | Age groups: Adolescents | Age groups: Adults | Age groups: Older adults | Endorsement area: None | Endorsement area: Clinical psychology | Endorsement area: Health psychology | Practice area: Psychology management/ administration | Practice area: Mental health intervention | Practice area: Counselling | Practice area: Organisational practice | Practice area: Perinatal | Sector: Private | Sector: Public | Setting: Private practice | Setting: Government department or agency | Setting: Community mental health service |
|---|---|---|---|---|---|---|---|---|---|---|---|---|---|---|---|---|---|---|
| Any | 14/18 | ✓ | ✓ | ✓ | ✓ | ✓ | ✓ | ✓ | ✓ | ✓ | ✓ | ✓ | ✓ | ✓ | ✓ | ✓ | ✓ | ✓ |
| Listening to natural sounds | 9/18 | ✓ | ✓ | ✓ | ✓ | ✓ | ✓ | ✓ | | ✓ | ✓ | | ✓ | ✓ | ✓ | ✓ | | |
| Having indoor pot plants | 10/18 | | ✓ | ✓ | ✓ | ✓ | ✓ | ✓ | ✓ | ✓ | ✓ | ✓ | ✓ | ✓ | ✓ | ✓ | ✓ | ✓ |
| Pictures of natural scenes | 8/18 | | ✓ | ✓ | ✓ | ✓ | ✓ | ✓ | ✓ | ✓ | ✓ | | ✓ | ✓ | ✓ | ✓ | | |
| Natural scents | 7/18 | ✓ | | ✓ | ✓ | ✓ | ✓ | ✓ | | ✓ | ✓ | | ✓ | ✓ | ✓ | ✓ | | |
| Travelling in a car or on public transport through natural environments | 8/18 | | ✓ | ✓ | ✓ | ✓ | ✓ | ✓ | | ✓ | ✓ | ✓ | ✓ | ✓ | ✓ | ✓ | ✓ | ✓ |
| Views of nature outside the window | 11/18 | | ✓ | ✓ | ✓ | ✓ | ✓ | ✓ | | ✓ | ✓ | ✓ | ✓ | ✓ | ✓ | ✓ | ✓ | ✓ |
| Open windows for fresh air and the smells and sounds of nature | 8/18 | | ✓ | ✓ | ✓ | ✓ | ✓ | ✓ | | ✓ | ✓ | ✓ | ✓ | ✓ | ✓ | ✓ | ✓ | ✓ |
| Nature-based virtual reality | 2/18 | | | ✓ | ✓ | ✓ | ✓ | | | ✓ | | | | ✓ | | ✓ | | |
| Other: sleep stories | 1/18 | | ✓ | | | | ✓ | | | ✓ | | | | ✓ | | ✓ | | |
| Other: discussions | 1/18 | | | ✓ | | | ✓ | | | ✓ | | | | ✓ | | ✓ | | |
| Other: displays of natural elements | 1/18 | | | ✓ | | | ✓ | | | ✓ | | | | ✓ | | ✓ | | |
| Other: videos | 1/18 | | | ✓ | | | ✓ | | | ✓ | | | | ✓ | | ✓ | | |
| Other: cut flowers | 1/18 | | | ✓ | | | ✓ | | | ✓ | | | | ✓ | | ✓ | | |
| Other: animal visits | 1/18 | | | ✓ | | | ✓ | | | ✓ | | | | ✓ | | ✓ | | |

Notes: Only headings relevant to our sample have been reported

### Direct engagement

**Table S2.23: Direct engagement in outdoor nature-based activities by psychologists in our sample with their patients/ clients**

| | | Age groups | | | | Endorsement area | | | Practice area | | | | | | Sector | | Setting | |
|---|---|---|---|---|---|---|---|---|---|---|---|---|---|---|---|---|---|---|
| Activity | Psychologists | Children | Adolescents | Adults | Older adults | None | Clinical psychology | Health psychology | Psychology management/ administration | Mental health intervention | Counselling | Personal development/ coaching | Organisational practice | Perinatal | Private | Public | Private practice | Government department or agency |
| Any | 14/18 | ✓ | ✓ | ✓ | ✓ | ✓ | ✓ | ✓ | ✓ | ✓ | ✓ | ✓ | ✓ | ✓ | ✓ | ✓ | ✓ | ✓ |
| Land-based gross motor activities | 2/18 | | | ✓ | ✓ | ✓ | ✓ | | | ✓ | | | | | ✓ | | ✓ | |
| Land-based fine motor activities | 3/18 | | | ✓ | ✓ | ✓ | ✓ | | | ✓ | | | | | ✓ | | ✓ | |
| Water-based physical activities | 1/18 | | | ✓ | | | ✓ | | | ✓ | | | | | ✓ | | ✓ | |
| Active transport | 0/18 | | | | | | | | | | | | | | | | | |
| Powered transport | 1/18 | | | | ✓ | ✓ | | | | ✓ | | | | | ✓ | | ✓ | |
| Gardening | 4/18 | | ✓ | ✓ | ✓ | ✓ | ✓ | | | ✓ | | ✓ | | | ✓ | | ✓ | |
| Ecological conservation or restoration | 1/18 | | | ✓ | | | ✓ | | | ✓ | | | | | ✓ | | ✓ | |
| Engaging with/ tending to animals | 1/18 | | | ✓ | | | ✓ | | | ✓ | | | | | ✓ | | ✓ | |
| Wilderness activities | 3/18 | | | ✓ | | ✓ | ✓ | | | ✓ | | | ✓ | | ✓ | ✓ | ✓ | ✓ |
| Nature play | 2/18 | ✓ | | ✓ | | ✓ | ✓ | | | ✓ | ✓ | | | | ✓ | ✓ | ✓ | |
| Nature observation activities | 6/18 | | | ✓ | | ✓ | ✓ | | | ✓ | ✓ | ✓ | ✓ | | ✓ | ✓ | ✓ | ✓ |
| Mindfulness/ meditation/ relaxation | 11/18 | ✓ | | ✓ | ✓ | ✓ | ✓ | | ✓ | ✓ | ✓ | ✓ | ✓ | | ✓ | ✓ | ✓ | ✓ |
| Forest bathing | 3/18 | | | ✓ | ✓ | ✓ | ✓ | | | ✓ | | ✓ | | | ✓ | | ✓ | |
| Social activities | 6/18 | ✓ | | ✓ | | ✓ | ✓ | | | ✓ | ✓ | | ✓ | | ✓ | ✓ | ✓ | ✓ |
| No specific activity | 7/18 | ✓ | | ✓ | | ✓ | ✓ | | ✓ | ✓ | ✓ | ✓ | ✓ | | ✓ | ✓ | ✓ | ✓ |
| Other activities: wildlife cruises | 1/18 | | | ✓ | | | ✓ | | | ✓ | | | | | ✓ | | ✓ | |

Notes: Only headings relevant to our sample have been reported.

**Table S2.24: Locations in which the psychologists in our sample engaged directly with their patients/ clients in outdoor nature-based activities**

| | | Age groups | | | | Endorsement area | | | Practice area | | | | | | Sector | | Setting | |
|---|---|---|---|---|---|---|---|---|---|---|---|---|---|---|---|---|---|---|
| **Location** | **Psychologists** | **Children** | **Adolescents** | **Adults** | **Older adults** | **None** | **Clinical psychology** | **Health psychology** | **Psychology management/ administration** | **Mental health intervention** | **Counselling** | **Personal development/ coaching** | **Organisational practice** | **Perinatal** | **Private** | **Public** | **Private practice** | **Government department or agency** |
| Private garden | 5/18 | ✓ | | ✓ | ✓ | ✓ | ✓ | | | ✓ | ✓ | ✓ | | | ✓ | ✓ | ✓ | |
| Community/ school garden | 4/18 | | | ✓ | ✓ | ✓ | | ✓ | | ✓ | | | ✓ | ✓ | ✓ | ✓ | ✓ | ✓ |
| Farm | 1/18 | ✓ | | | | ✓ | | | | | ✓ | | | | | ✓ | | ✓ |
| Public park | 7/18 | | | ✓ | ✓ | ✓ | ✓ | ✓ | ✓ | ✓ | | | ✓ | ✓ | ✓ | ✓ | ✓ | ✓ |
| National park | 3/18 | | | ✓ | | | ✓ | | | ✓ | | | | | ✓ | | ✓ | |
| Forest | 4/18 | | | ✓ | | ✓ | ✓ | | | ✓ | | ✓ | | | ✓ | | ✓ | |
| Oval/ sports field | 2/18 | | | ✓ | | | ✓ | ✓ | | ✓ | | | | ✓ | ✓ | | ✓ | |
| On the lake | 1/18 | | | ✓ | | | ✓ | | | ✓ | | | | | ✓ | | ✓ | |
| Near the lake | 3/18 | | | ✓ | | ✓ | ✓ | | | ✓ | | | ✓ | | ✓ | ✓ | ✓ | ✓ |
| On the river | 0/18 | | | | | | | | | | | | | | | | | |
| Near the river | 2/18 | | | ✓ | | | ✓ | | | ✓ | | | | | ✓ | | ✓ | |
| On the ocean | 1/18 | | | ✓ | | | ✓ | | | ✓ | | | | | ✓ | | ✓ | |
| Near the ocean | 3/18 | | | ✓ | ✓ | ✓ | ✓ | | | ✓ | | | | | ✓ | | ✓ | |
| Wetlands | 1/18 | | | ✓ | | ✓ | | | | ✓ | | | | | ✓ | | ✓ | |
| Café garden | 1/18 | | | ✓ | | ✓ | | | | ✓ | | | | | ✓ | | ✓ | |
| Carpark with garden supplies | 1/18 | | ✓ | | | ✓ | | | | ✓ | | | | | ✓ | | ✓ | |

Notes: Only headings relevant to our sample have been reported

**Table S2.25: Direct engagement in indoor nature-based activities by psychologists in our sample with their patients/ clients**

| | | Age groups | | | | Endorsement area | | | Practice area | | | | | Sector | | Setting | | |
|---|---|---|---|---|---|---|---|---|---|---|---|---|---|---|---|---|---|---|
| **Activity** | **Psychologists** | **Children** | **Adolescents** | **Adults** | **Older adults** | **None** | **Clinical psychology** | **Health psychology** | **Psychology management/ administration** | **Mental health intervention** | **Counselling** | **Organisational practice** | **Perinatal** | **Private** | **Public** | **Private practice** | **Government department or agency** | **Community mental health service** |
| Any | 15/18 | ✓ | ✓ | ✓ | ✓ | ✓ | ✓ | ✓ | ✓ | ✓ | ✓ | ✓ | ✓ | ✓ | ✓ | ✓ | ✓ | ✓ |
| Listening to natural sounds | 11/18 | ✓ | ✓ | ✓ | ✓ | ✓ | ✓ | ✓ | | ✓ | ✓ | | ✓ | ✓ | ✓ | ✓ | | ✓ |
| Having indoor pot plants | 14/18 | ✓ | ✓ | ✓ | ✓ | ✓ | ✓ | ✓ | ✓ | ✓ | ✓ | ✓ | ✓ | ✓ | ✓ | ✓ | ✓ | ✓ |
| Pictures of natural scenes | 8/18 | | ✓ | ✓ | ✓ | ✓ | ✓ | ✓ | ✓ | ✓ | ✓ | | ✓ | ✓ | ✓ | ✓ | | |
| Natural scents | 7/18 | ✓ | ✓ | ✓ | ✓ | ✓ | ✓ | ✓ | | ✓ | ✓ | | ✓ | ✓ | ✓ | ✓ | | |
| Travelling in a car or on public transport through natural environments | 2/18 | | | ✓ | | ✓ | ✓ | | | ✓ | ✓ | | | ✓ | | ✓ | | |
| Views of nature outside the window | 11/18 | | ✓ | ✓ | ✓ | ✓ | ✓ | ✓ | | ✓ | ✓ | ✓ | ✓ | ✓ | ✓ | ✓ | ✓ | ✓ |
| Open windows for fresh air and the smells and sounds of nature | 6/18 | | | ✓ | ✓ | ✓ | ✓ | | | ✓ | ✓ | | | ✓ | | ✓ | | |
| Nature-based virtual reality | 3/18 | | | ✓ | ✓ | ✓ | ✓ | | | ✓ | | | | ✓ | | ✓ | | |
| Other: discussions | 1/18 | | | ✓ | | | ✓ | | | ✓ | | | | ✓ | | ✓ | | |
| Other: documentaries | 1/18 | | | ✓ | | | ✓ | | | ✓ | | | | ✓ | | ✓ | | |
| Other: natural items | 1/18 | | | ✓ | | | ✓ | | | ✓ | | | | ✓ | | ✓ | | |
| Other: animal visits | 1/18 | | | ✓ | | | ✓ | | | ✓ | | | | ✓ | | ✓ | | |
| Other: cut flowers | 1/18 | | | ✓ | | | ✓ | | | ✓ | | | | ✓ | | ✓ | | |
| Other: gardening | 1/18 | | ✓ | | | | ✓ | | | ✓ | | | | ✓ | | ✓ | | |
| Other: art | 1/18 | | ✓ | | | | ✓ | | | ✓ | | | | ✓ | | ✓ | | |
| Other: guided imagery | 1/18 | | ✓ | | | | ✓ | | | ✓ | | | | ✓ | | ✓ | | |
| Other: metaphors | 1/18 | | ✓ | | | | ✓ | | | ✓ | | | | ✓ | | ✓ | | |

Notes: Only headings relevant to our sample have been reported

### Enhancing outdoor natural environments

**Table S2.26: Locations that psychologists in our sample have advocated for the addition of or improvement to**

| | | Age groups | | | Endorsement area | | | | Practice area | | | | | Sector | | Setting | | |
|---|---|---|---|---|---|---|---|---|---|---|---|---|---|---|---|---|---|---|
| **Activity** | **Psychologists** | **Adolescents** | **Adults** | **Older adults** | **None** | **Clinical psychology** | **Clinical psychology & counselling psychology** | **Health psychology** | **Mental health intervention** | **Counselling** | **Organisational practice** | **Teaching/ supervisory** | **Perinatal** | **Private** | **Public** | **Private practice** | **Government department or agency** | **Community mental health service** |
| Any | 13/18 | ✓ | ✓ | ✓ | ✓ | ✓ | ✓ | ✓ | ✓ | ✓ | ✓ | ✓ | ✓ | ✓ | ✓ | ✓ | ✓ | ✓ |
| Gardens or other natural element in care settings | 8/18 | ✓ | ✓ | ✓ | ✓ | ✓ | ✓ | ✓ | ✓ | ✓ | | ✓ | ✓ | ✓ | ✓ | ✓ | | ✓ |
| Nature elements inside care settings | 7/18 | ✓ | ✓ | ✓ | ✓ | ✓ | | ✓ | ✓ | ✓ | | | ✓ | ✓ | ✓ | ✓ | | ✓ |
| Nature elements in non-care, public settings | 8/18 | ✓ | ✓ | ✓ | ✓ | ✓ | | ✓ | ✓ | ✓ | ✓ | | ✓ | ✓ | ✓ | ✓ | ✓ | ✓ |
| Community gardens | 7/18 | | ✓ | ✓ | ✓ | ✓ | | ✓ | ✓ | ✓ | | ✓ | ✓ | ✓ | | ✓ | | |
| School/ childcare gardens | 6/18 | ✓ | ✓ | ✓ | ✓ | ✓ | | ✓ | ✓ | ✓ | | | ✓ | ✓ | | ✓ | | |
| Public parks/ gardens | 6/18 | | ✓ | | ✓ | ✓ | ✓ | ✓ | ✓ | ✓ | | ✓ | ✓ | ✓ | | ✓ | | |
| Green corridors | 4/18 | | ✓ | ✓ | ✓ | ✓ | | | ✓ | ✓ | | | | ✓ | | ✓ | | |
| Blue spaces/ water bodies | 3/18 | | ✓ | | | ✓ | ✓ | | ✓ | ✓ | | ✓ | | ✓ | | ✓ | | |
| Other: Coastal park | 1/18 | | ✓ | | | ✓ | | | ✓ | | | ✓ | | ✓ | | ✓ | | |

Notes: Only headings relevant to our sample have been reported

**Table S2.27: Locations that psychologists in our sample have directly engaged in the addition of or improvement to**

| Activity | Psychologists | Age groups: Children | Adolescents | Adults | Older adults | Endorsement area: None | Clinical psychology | Clinical psychology & counselling psychology | Health psychology | Practice area: Psychology management/ administration | Mental health intervention | Counselling | Personal development/ coaching | Organisational practice | Teaching/ supervisory | Perinatal | Sector: Private | Public | Setting: Private practice | Government department or agency |
|---|---|---|---|---|---|---|---|---|---|---|---|---|---|---|---|---|---|---|---|---|
| Any | 14/19 | ✓ | ✓ | ✓ | ✓ | ✓ | ✓ | ✓ | ✓ | ✓ | ✓ | ✓ | ✓ | ✓ | ✓ | ✓ | ✓ | ✓ | ✓ | ✓ |
| Gardens or other natural element in care settings | 5/19 | | | ✓ | ✓ | ✓ | ✓ | ✓ | | ✓ | ✓ | ✓ | | | ✓ | | ✓ | ✓ | ✓ | |
| Nature elements inside care settings | 4/19 | ✓ | ✓ | ✓ | ✓ | ✓ | ✓ | | | | ✓ | ✓ | | | | | ✓ | ✓ | ✓ | |
| Nature elements in non-care, public settings | 8/19 | ✓ | ✓ | ✓ | ✓ | ✓ | ✓ | | | | ✓ | ✓ | ✓ | | ✓ | | ✓ | ✓ | ✓ | |
| Community gardens | 6/19 | | | ✓ | ✓ | ✓ | ✓ | | | | ✓ | | ✓ | | ✓ | | ✓ | | ✓ | |
| School/ childcare gardens | 4/19 | | ✓ | ✓ | ✓ | ✓ | ✓ | | | | ✓ | | | ✓ | | | ✓ | ✓ | ✓ | ✓ |
| Public parks/ gardens | 5/19 | | | ✓ | | ✓ | ✓ | | | | ✓ | | ✓ | | ✓ | | ✓ | | ✓ | |
| Green corridors | 5/19 | | | ✓ | ✓ | ✓ | ✓ | | | | ✓ | | ✓ | | | | ✓ | | ✓ | |
| Blue spaces/ water bodies | 3/19 | | | ✓ | | ✓ | ✓ | | | | ✓ | | ✓ | | ✓ | | ✓ | | ✓ | |
| Other | 0/19 | | | | | | | | | | | | | | | | | | | |

Notes: Only headings relevant to our sample have been reported

### Social work

**Table S2.28: Social worker respondent demographics**

| | Stage 2 (n=15) |
|---|---|
| Gender | |
| Woman | 14/15 |
| Man | 1/15 |
| Age group | |
| 20-29 years | 0/15 |
| 30-39 years | 4/15 |
| 40-49 years | 5/15 |
| 50-59 years | 3/15 |
| 60+ years | 3/15 |
| Trained in Australia (pre-registration qualification) | 15/15 |
| Years worked in discipline, median (interquartile range) | 12 (6-20) |
| Main patient age group | |
| Children | 0/15 |
| Adolescents | 3/15 |
| Adults | 11/15 |
| Older adults | 1/15 |
| Role | |
| Clinician | 13/15 |
| Community worker | 1/15 |
| Manager | 1/15 |
| Field | |
| Education | 1/15 |
| Family violence | 1/15 |
| Family support | 1/15 |
| Mental health | 8/15 |
| Health | 2/15 |
| Community work | 1/15 |
| Youth | 1/15 |
| Setting | |
| Private practice | 5/15 |
| Hospital | 1/15 |
| Other community health care service | 4/15 |
| Education facility | 1/15 |
| Community development | 1/15 |
| Youth-based NFP | 1/15 |
| Other government department | 1/15 |
| Community garden community worker | 1/15 |
| Private sector | 7/15 |
| Sees patients/ clients | 15/15 |
| Telehealth | 15/15 |
| Home visits only | 2/15 |
| Main practice in a major city | 7/13 |
| Hours worked in main practice setting, median (interquartile range) | 37 (22-38) |

### Recommendations

**Table S2.29: Recommendations for outdoor nature-based activities from the social workers in our sample**

| | | Age groups | | | Field | | | | | | Sector | | Setting | | | | | | |
|---|---|---|---|---|---|---|---|---|---|---|---|---|---|---|---|---|---|---|---|
| **Activity** | **Social workers** | **Adolescents** | **Adults** | **Older adults** | **Education** | **Family support** | **Mental health** | **Health** | **Community work** | **Youth** | **Private** | **Public** | **Private practice** | **Hospital** | **Other community health care service** | **Education facility** | **Community development** | **Youth-based not for profit** | **Community garden as a community worker** |
| Any | 14/15 | ✓ | ✓ | ✓ | ✓ | ✓ | ✓ | ✓ | ✓ | ✓ | ✓ | ✓ | ✓ | ✓ | ✓ | ✓ | ✓ | ✓ | ✓ |
| Land-based gross motor activities | 9/15 | ✓ | ✓ | ✓ | ✓ | | ✓ | ✓ | ✓ | ✓ | ✓ | ✓ | ✓ | | ✓ | ✓ | ✓ | ✓ | ✓ |
| Land-based fine motor activities | 7/15 | ✓ | ✓ | ✓ | | | ✓ | ✓ | ✓ | ✓ | ✓ | ✓ | ✓ | | ✓ | | ✓ | ✓ | ✓ |
| Water-based physical activities | 6/15 | | ✓ | ✓ | | | ✓ | ✓ | ✓ | | ✓ | ✓ | ✓ | | ✓ | | ✓ | | ✓ |
| Active transport | 4/15 | ✓ | ✓ | | | | ✓ | | ✓ | ✓ | ✓ | ✓ | ✓ | | ✓ | | | ✓ | ✓ |
| Powered transport | 1/15 | | | ✓ | | | | ✓ | | | ✓ | | | | | | ✓ | | |
| Gardening | 11/15 | ✓ | ✓ | ✓ | ✓ | | ✓ | ✓ | ✓ | ✓ | ✓ | ✓ | ✓ | ✓ | ✓ | ✓ | ✓ | ✓ | ✓ |
| Ecological conservation or restoration | 4/15 | | ✓ | ✓ | ✓ | | ✓ | ✓ | ✓ | | ✓ | ✓ | ✓ | | | ✓ | ✓ | | ✓ |
| Engaging with/ tending to animals | 11/15 | ✓ | ✓ | ✓ | ✓ | | ✓ | ✓ | ✓ | ✓ | ✓ | ✓ | ✓ | | ✓ | ✓ | ✓ | ✓ | ✓ |
| Wilderness activities | 2/15 | | ✓ | ✓ | | | | ✓ | ✓ | | ✓ | ✓ | | | | | ✓ | | ✓ |
| Nature play | 5/15 | | ✓ | | | ✓ | ✓ | | ✓ | | ✓ | ✓ | ✓ | | ✓ | | | | ✓ |
| Nature observation activities | 8/15 | ✓ | ✓ | ✓ | | | ✓ | ✓ | ✓ | ✓ | ✓ | ✓ | ✓ | | ✓ | | ✓ | ✓ | ✓ |
| Mindfulness/ meditation/ relaxation | 13/15 | ✓ | ✓ | ✓ | | ✓ | ✓ | ✓ | ✓ | ✓ | ✓ | ✓ | ✓ | ✓ | ✓ | | ✓ | ✓ | ✓ |
| Forest bathing | 2/15 | | ✓ | | | | ✓ | | | | ✓ | | ✓ | | | | | | |
| Social activities | 9/15 | ✓ | ✓ | ✓ | | ✓ | ✓ | ✓ | ✓ | | ✓ | ✓ | ✓ | | ✓ | | ✓ | | ✓ |
| No specific activity | 13/15 | ✓ | ✓ | ✓ | ✓ | ✓ | ✓ | ✓ | ✓ | ✓ | ✓ | ✓ | ✓ | ✓ | ✓ | ✓ | ✓ | ✓ | ✓ |
| Other activities | 0/15 | | | | | | | | | | | | | | | | | | |

Notes: Only headings relevant to our sample have been reported

**Table S2.30: Locations in which outdoor nature-based activities were recommended by the social workers in our sample**

| | | Age groups | | | Field | | | | | | Sector | | Setting | | | | | | |
|---|---|---|---|---|---|---|---|---|---|---|---|---|---|---|---|---|---|---|---|
| Location | Social workers | Adolescents | Adults | Older adults | Education | Family support | Mental health | Health | Community work | Youth | Private | Public | Private practice | Hospital | Other community health care service | Education facility | Community development | Youth-based not for profit | Community garden as a community worker |
| Private garden | 12/15 | ✓ | ✓ | ✓ | | ✓ | ✓ | ✓ | ✓ | ✓ | ✓ | ✓ | ✓ | | ✓ | | ✓ | ✓ | ✓ |
| Community/ school garden | 9/15 | | ✓ | ✓ | ✓ | | ✓ | ✓ | ✓ | | ✓ | ✓ | ✓ | | ✓ | ✓ | ✓ | | ✓ |
| Farm | 3/15 | | ✓ | ✓ | | | ✓ | ✓ | | | ✓ | ✓ | | ✓ | ✓ | | ✓ | | |
| Public park | 13/15 | ✓ | ✓ | ✓ | ✓ | ✓ | ✓ | ✓ | ✓ | ✓ | ✓ | ✓ | ✓ | | ✓ | ✓ | ✓ | ✓ | ✓ |
| National park | 5/15 | | ✓ | ✓ | | | ✓ | ✓ | | | ✓ | ✓ | ✓ | | ✓ | | ✓ | | |
| Forest | 5/15 | | ✓ | ✓ | | | ✓ | ✓ | | | ✓ | ✓ | ✓ | | ✓ | | ✓ | | |
| Oval/ sports field | 1/15 | | | ✓ | | | | ✓ | | | ✓ | | | | | | ✓ | | |
| On the lake | 2/15 | | ✓ | ✓ | | | ✓ | ✓ | | | ✓ | | ✓ | | | | ✓ | | |
| Near the lake | 5/15 | ✓ | ✓ | | | | ✓ | | | ✓ | ✓ | ✓ | ✓ | ✓ | ✓ | | | ✓ | |
| On the river | 3/15 | | ✓ | ✓ | | | ✓ | ✓ | | | ✓ | ✓ | ✓ | | ✓ | | ✓ | | |
| Near the river | 6/15 | ✓ | ✓ | ✓ | | | ✓ | ✓ | | ✓ | ✓ | ✓ | ✓ | ✓ | ✓ | | ✓ | ✓ | |
| On the ocean | 6/15 | | ✓ | ✓ | | | ✓ | ✓ | ✓ | | ✓ | ✓ | ✓ | | ✓ | | ✓ | | ✓ |
| Near the ocean | 9/15 | ✓ | ✓ | ✓ | | | ✓ | ✓ | | | ✓ | ✓ | ✓ | | ✓ | | ✓ | | |
| Other | 0/15 | | | | | | | | | | | | | | | | | | |

**Table S2.31: Recommendations for indoor nature-based activities from the social workers in our sample**

| | | Age groups | | | Field | | | | | | | Sector | | Setting | | | | | | | |
|---|---|---|---|---|---|---|---|---|---|---|---|---|---|---|---|---|---|---|---|---|---|
| Activity | Social workers | Adolescents | Adults | Older adults | Education | Family violence | Family support | Mental health | Health | Community work | Youth | Private | Public | Private practice | Hospital | Other community health care service | Education facility | Community development | Youth-based not for profit | Other government department or agency | Community garden as a community worker |
| Any | 14/15 | ✓ | ✓ | ✓ | ✓ | ✓ | ✓ | ✓ | ✓ | ✓ | ✓ | ✓ | ✓ | ✓ | ✓ | ✓ | ✓ | ✓ | ✓ | ✓ | ✓ |
| Listening to natural sounds | 8/15 | ✓ | ✓ | | ✓ | | ✓ | ✓ | ✓ | | ✓ | ✓ | ✓ | ✓ | | ✓ | ✓ | | ✓ | | ✓ |
| Having indoor pot plants | 8/15 | ✓ | ✓ | | ✓ | ✓ | ✓ | ✓ | ✓ | ✓ | | ✓ | ✓ | ✓ | | ✓ | ✓ | | | ✓ | ✓ |
| Pictures of natural scenes | 3/15 | | ✓ | | | | ✓ | ✓ | | | | ✓ | | ✓ | | ✓ | | | | | |
| Natural scents | 6/15 | ✓ | ✓ | | | | ✓ | ✓ | ✓ | | | ✓ | ✓ | ✓ | ✓ | ✓ | | | | | |
| Travelling in a car or on public transport through natural environments | 3/15 | | ✓ | | | | | ✓ | | | | ✓ | ✓ | ✓ | | ✓ | | | | | |
| Views of nature outside the window | 6/15 | ✓ | ✓ | ✓ | | | | ✓ | ✓ | | ✓ | ✓ | ✓ | ✓ | | ✓ | | ✓ | ✓ | | |
| Open windows for fresh air and the smells and sounds of nature | 7/15 | ✓ | ✓ | ✓ | | | ✓ | ✓ | ✓ | ✓ | | ✓ | ✓ | ✓ | | ✓ | | ✓ | | | ✓ |
| Nature-based virtual reality | 0/15 | | | | | | | | | | | | | | | | | | | | |
| Other: sand tray | 1/15 | | ✓ | | | | | ✓ | | | | ✓ | | ✓ | | | | | | | |
| Other: Cut flowers | 2/15 | | ✓ | | | | | ✓ | | ✓ | | ✓ | ✓ | ✓ | | | | | | | ✓ |
| Other: Herbs | 1/15 | | ✓ | | | | | ✓ | | | | ✓ | | ✓ | | | | | | | |
| Other: Sharing pot plants with local café | 1/15 | | ✓ | | | | | | | ✓ | | | ✓ | | | | | | | | ✓ |

Notes: Only headings relevant to our sample have been reported

### Direct engagement

**Table S2.32: Direct engagement in outdoor nature-based activities by social workers in our sample with their patients/ clients**

| | | Age groups | | | Field | | | | | | Sector | | Setting | | | | | | | |
|---|---|---|---|---|---|---|---|---|---|---|---|---|---|---|---|---|---|---|---|---|
| **Activity** | **Social workers** | **Adolescents** | **Adults** | **Older adults** | **Education** | **Family violence** | **Mental health** | **Health** | **Community work** | **Youth** | **Private** | **Public** | **Private practice** | **Hospital** | **Other community health care service** | **Education facility** | **Community development** | **Youth-based not for profit** | **Other government department or agency** | **Community garden as a community worker** |
| Any | 13/15 | ✓ | ✓ | ✓ | ✓ | ✓ | ✓ | ✓ | ✓ | ✓ | ✓ | ✓ | ✓ | ✓ | ✓ | ✓ | ✓ | ✓ | ✓ | ✓ |
| Land-based gross motor activities | 3/15 | | ✓ | | | | ✓ | | ✓ | | ✓ | ✓ | ✓ | | ✓ | | | | | ✓ |
| Land-based fine motor activities | 4/15 | ✓ | ✓ | | | ✓ | ✓ | | ✓ | ✓ | ✓ | ✓ | ✓ | | | | | ✓ | ✓ | |
| Water-based physical activities | 0/15 | | | | | | | | | | | | | | | | | | | |
| Active transport | 1/15 | | ✓ | | | | | | ✓ | | | ✓ | | | | | | | | ✓ |
| Powered transport | 0/15 | | | | | | | | | | | | | | | | | | | |
| Gardening | 1/15 | | ✓ | | | | | | ✓ | | | ✓ | | | | | | | | ✓ |
| Ecological conservation or restoration | 1/15 | | ✓ | | | | | | ✓ | | | ✓ | | | | | | | | ✓ |
| Engaging with/ tending to animals | 2/15 | ✓ | ✓ | | | | | ✓ | ✓ | | | ✓ | | | ✓ | | | | | ✓ |
| Wilderness activities | 0/15 | | | | | | | | | | | | | | | | | | | |
| Nature play | 2/15 | | ✓ | | | | ✓ | | ✓ | | ✓ | ✓ | ✓ | | | | | | | ✓ |
| Nature observation activities | 2/15 | ✓ | | ✓ | | | | ✓ | | ✓ | ✓ | ✓ | | | | | ✓ | ✓ | | |
| Mindfulness/ meditation/ relaxation | 11/15 | ✓ | ✓ | ✓ | | ✓ | ✓ | ✓ | ✓ | ✓ | ✓ | ✓ | ✓ | ✓ | ✓ | | ✓ | ✓ | ✓ | ✓ |
| Forest bathing | 0/15 | | | | | | | | | | | | | | | | | | | |
| Social activities | 3/15 | | ✓ | | | | ✓ | | ✓ | | ✓ | ✓ | ✓ | | ✓ | | | | | ✓ |
| No specific activity | 11/15 | ✓ | ✓ | ✓ | ✓ | ✓ | ✓ | ✓ | ✓ | ✓ | ✓ | ✓ | ✓ | ✓ | ✓ | ✓ | ✓ | ✓ | ✓ | ✓ |
| Other activities | 0/15 | | | | | | | | | | | | | | | | | | | |

Notes: Only headings relevant to our sample have been reported.

**Table S2.33: Locations in which the social workers in our sample engaged directly with their patients/ clients in outdoor nature-based activities**

| | | Age groups | | | Field | | | | | Sector | | Setting | | | | | | |
|---|---|---|---|---|---|---|---|---|---|---|---|---|---|---|---|---|---|---|
| **Location** | **Social workers** | **Adolescents** | **Adults** | **Older adults** | **Family violence** | **Mental health** | **Health** | **Community work** | **Youth** | **Private** | **Public** | **Private practice** | **Hospital** | **Other community health care service** | **Community development** | **Youth-based not for profit** | **Other government department or agency** | **Community garden as a community worker** |
| Private garden | 4/15 | ✓ | ✓ | ✓ | | ✓ | ✓ | | ✓ | ✓ | ✓ | ✓ | | ✓ | ✓ | ✓ | | |
| Community/ school garden | 5/15 | ✓ | ✓ | | ✓ | ✓ | | ✓ | ✓ | ✓ | ✓ | ✓ | | ✓ | | ✓ | ✓ | ✓ |
| Farm | 0/15 | | | | | | | | | | | | | | | | | |
| Public park | 9/15 | ✓ | ✓ | ✓ | ✓ | ✓ | ✓ | ✓ | ✓ | ✓ | ✓ | ✓ | ✓ | ✓ | ✓ | ✓ | ✓ | ✓ |
| National park | 0/15 | | | | | | | | | | | | | | | | | |
| Forest | 0/15 | | | | | | | | | | | | | | | | | |
| Oval/ sports field | 4/15 | ✓ | ✓ | | ✓ | ✓ | | | ✓ | ✓ | ✓ | ✓ | | ✓ | | ✓ | ✓ | |
| On the lake | 0/15 | | | | | | | | | | | | | | | | | |
| Near the lake | 3/15 | ✓ | ✓ | | | ✓ | | | ✓ | ✓ | ✓ | ✓ | ✓ | | | ✓ | | |
| On the river | 0/15 | | | | | | | | | | | | | | | | | |
| Near the river | 1/15 | | ✓ | | | ✓ | | | | | ✓ | | ✓ | | | | | |
| On the ocean | 0/15 | | | | | | | | | | | | | | | | | |
| Near the ocean | 1/15 | | ✓ | | | ✓ | | | | ✓ | | ✓ | | | | | | |
| Other | 0/15 | | | | | | | | | | | | | | | | | |

Notes: Only headings relevant to our sample have been reported

**Table S2.34: Direct engagement in indoor nature-based activities by social workers in our sample with their patients/ clients**

| | | Age groups | | | Field | | | | | | | Sector | | Setting | | | | | | | |
|---|---|---|---|---|---|---|---|---|---|---|---|---|---|---|---|---|---|---|---|---|---|
| **Activity** | **Social workers** | **Adolescents** | **Adults** | **Older adults** | **Education** | **Family violence** | **Family support** | **Mental health** | **Health** | **Community work** | **Youth** | **Private** | **Public** | **Private practice** | **Hospital** | **Other community health care service** | **Education facility** | **Community development** | **Youth-based not for profit** | **Other government department or agency** | **Community garden as a community worker** |
| Any | 13/15 | ✓ | ✓ | ✓ | ✓ | ✓ | ✓ | ✓ | ✓ | ✓ | ✓ | ✓ | ✓ | ✓ | ✓ | ✓ | ✓ | ✓ | ✓ | ✓ | ✓ |
| Listening to natural sounds | 4/15 | ✓ | ✓ | | | | | ✓ | ✓ | ✓ | ✓ | ✓ | ✓ | ✓ | | ✓ | | | ✓ | | ✓ |
| Having indoor pot plants | 9/15 | ✓ | ✓ | | ✓ | ✓ | | ✓ | ✓ | ✓ | | ✓ | ✓ | ✓ | | ✓ | ✓ | | | ✓ | ✓ |
| Pictures of natural scenes | 7/15 | ✓ | ✓ | | ✓ | ✓ | ✓ | ✓ | | | | ✓ | ✓ | ✓ | | ✓ | ✓ | | | ✓ | |
| Natural scents | 4/15 | ✓ | ✓ | | | | | ✓ | ✓ | | | ✓ | ✓ | ✓ | ✓ | ✓ | | | | | |
| Travelling in a car or on public transport through natural environments | 1/15 | ✓ | | | | | | | | | ✓ | | ✓ | | | | | | ✓ | | |
| Views of nature outside the window | 4/15 | ✓ | ✓ | | ✓ | ✓ | | ✓ | | | ✓ | ✓ | ✓ | ✓ | | | ✓ | | ✓ | ✓ | |
| Open windows for fresh air and the smells and sounds of nature | 4/15 | ✓ | ✓ | ✓ | | ✓ | | ✓ | ✓ | | ✓ | ✓ | ✓ | ✓ | | | | ✓ | ✓ | ✓ | |
| Nature-based virtual reality | 1/15 | | ✓ | | | | ✓ | | | | | ✓ | | | | ✓ | | | | | |
| Other: salt lamp | 1/15 | | ✓ | | | | | ✓ | | | | ✓ | | ✓ | | | | | | | |
| Other: gardening | 1/15 | | ✓ | | | | | | | ✓ | | | ✓ | | | | | | | | ✓ |

Notes: Only headings relevant to our sample have been reported

### Enhancing outdoor natural environments

**Table S2.35: Locations that social workers in our sample have advocated for the addition of or improvement to**

| Location | Social workers | Age groups | | | Field | | | | | | | Sector | | Setting | | | | | | | |
|---|---|---|---|---|---|---|---|---|---|---|---|---|---|---|---|---|---|---|---|---|---|
| | | Adolescents | Adults | Older adults | Education | Family violence | Family support | Mental health | Health | Community work | Youth | Private | Public | Private practice | Hospital | Other community health care service | Education facility | Community development | Youth-based not for profit | Other government department or agency | Community garden as a community worker |
| Any | 11/15 | ✓ | ✓ | ✓ | ✓ | ✓ | ✓ | ✓ | ✓ | ✓ | ✓ | ✓ | ✓ | ✓ | ✓ | ✓ | ✓ | ✓ | ✓ | ✓ | ✓ |
| Gardens or other natural element in care settings | 8/15 | ✓ | ✓ | ✓ | ✓ | ✓ | | ✓ | ✓ | ✓ | ✓ | ✓ | ✓ | ✓ | ✓ | ✓ | ✓ | ✓ | ✓ | ✓ | ✓ |
| Nature elements inside care settings | 3/15 | ✓ | ✓ | ✓ | | | | | ✓ | ✓ | ✓ | ✓ | ✓ | | | | | ✓ | ✓ | | ✓ |
| Nature elements in non-care, public settings | 7/15 | ✓ | ✓ | ✓ | ✓ | ✓ | ✓ | ✓ | ✓ | ✓ | ✓ | ✓ | ✓ | | | ✓ | ✓ | ✓ | ✓ | ✓ | ✓ |
| Community gardens | 4/15 | | ✓ | | ✓ | | | ✓ | | ✓ | | ✓ | ✓ | ✓ | | | ✓ | | | | ✓ |
| School/ childcare gardens | 3/15 | ✓ | ✓ | | | ✓ | | ✓ | | ✓ | | ✓ | ✓ | ✓ | | | | | | ✓ | ✓ |
| Public parks/ gardens | 2/15 | | ✓ | | | | | ✓ | | ✓ | | ✓ | ✓ | ✓ | | | | | | | ✓ |
| Green corridors | 1/15 | | ✓ | | | | | | | ✓ | | | ✓ | | | | | | | | ✓ |
| Blue spaces/ water bodies | 2/15 | | ✓ | | | | | ✓ | | ✓ | | ✓ | ✓ | ✓ | | | | | | | ✓ |
| Other: street trees | 1/15 | | ✓ | | | | | | | ✓ | | | ✓ | | | | | | | | ✓ |

Notes: Only headings relevant to our sample have been reported

**Table S2.36: Locations that social workers in our sample have directly engaged in the addition of or improvement to**

| | | Age groups | | | Field | | | | | Sector | | Setting | | | | |
|---|---|---|---|---|---|---|---|---|---|---|---|---|---|---|---|---|
| **Location** | **Social workers** | **Adolescents** | **Adults** | **Older adults** | **Education** | **Family violence** | **Mental health** | **Community work** | **Youth** | **Private** | **Public** | **Private practice** | **Education facility** | **Youth-based not for profit** | **Other government department or agency** | **Community garden as a community worker** |
| Any | 6/15 | ✓ | ✓ | | ✓ | ✓ | ✓ | ✓ | ✓ | ✓ | ✓ | ✓ | ✓ | ✓ | ✓ | ✓ |
| Gardens or other natural element in care settings | 2/15 | | ✓ | | | | ✓ | ✓ | | ✓ | ✓ | ✓ | | | | ✓ |
| Nature elements inside care settings | 1/15 | | ✓ | | | | | ✓ | | | ✓ | | | | | ✓ |
| Nature elements in non-care, public settings | 4/15 | ✓ | ✓ | | ✓ | ✓ | | ✓ | ✓ | | ✓ | | ✓ | ✓ | ✓ | ✓ |
| Community gardens | 2/15 | | ✓ | | | | ✓ | ✓ | | ✓ | ✓ | ✓ | | | | ✓ |
| School/ childcare gardens | 2/15 | ✓ | ✓ | | | ✓ | | ✓ | | | ✓ | | | | ✓ | ✓ |
| Public parks/ gardens | 1/15 | | ✓ | | | | | ✓ | | | ✓ | | | | | ✓ |
| Green corridors | 1/15 | | ✓ | | | | | ✓ | | | ✓ | | | | | ✓ |
| Blue spaces/ water bodies | 1/15 | | ✓ | | | | | ✓ | | | ✓ | | | | | ✓ |
| Other: Aboriginal outdoor space in hospital | 1/15 | | ✓ | | | | ✓ | | | ✓ | | ✓ | | | | |
| Other: urban tree canopy campaigns | 1/15 | | ✓ | | | | | ✓ | | | ✓ | | | | | ✓ |

Notes: Only headings relevant to our sample have been reported

### Speech pathology

**Table S2.37: Speech pathology respondent demographics**

| | Stage 1 (n=12) | Stage 2 (n=4) |
|---|---|---|
| Gender | | |
| Woman | 11/12 | 4/4 |
| Man | 1/12 | 0/4 |
| Age group | | |
| 20-29 years | 2/12 | 0/4 |
| 30-39 years | 7/12 | 2/4 |
| 40-49 years | 3/12 | 2/4 |
| 50-59 years | 0/12 | 0/4 |
| 60+ years | 0/12 | 0/4 |
| Trained in Australia (pre-registration qualification) | 12/12 | 4/4 |
| Years worked in discipline, median (interquartile range) | 7 (4-13) | 11 (10-14) |
| Main patient age group | | |
| Children | NA | 4/4 |
| Adolescents | NA | 0/4 |
| Adults | NA | 0/4 |
| Older adults | NA | 0/4 |
| Role | | |
| Teacher/ educator | NA | 0/4 |
| Clinician | NA | 4/4 |
| Administration | NA | 0/4 |
| Discharge planning | NA | 0/4 |
| Area | | |
| Speech and language | 6/12 | 3/4 |
| Mental health and pragmatic language | 0/12 | 1/4 |
| Communication | 1/12 | 0/4 |
| Leadership/ management | 2/12 | 0/4 |
| Voice | 1/12 | 0/4 |
| Swallowing and mealtimes | 1/12 | 0/4 |
| Research | 1/12 | 0/4 |
| Setting | | |
| Private practice | 4/11 | 3/4 |
| Home and community based service | 1/11 | 0/4 |
| Hospital | 1/11 | 0/4 |
| Disability service | 0/11 | 1/4 |
| Educational facility | 1/11 | 0/4 |
| Community | 3/11 | 0/4 |
| Residential care | 1/11 | 0/4 |
| Private sector | 8/11 | 4/4 |
| Sees patients/ clients | 11/12 | 4/4 |
| Telehealth | 10/12 | 3/4 |
| Home visits only | NA | 0/4 |
| Main practice in a major city^ | 5/11 | 3/4 |
| Hours worked in main practice setting (range) | 24 (9-38) | 17 (13-23) |

Note: NA: Not asked. ^Stage 2 respondents were only asked for the postcode of their main practice if they did not solely conduct home visits.

### Recommendations

**Table S2.38: Recommendations for outdoor nature-based activities from the speech pathologists in our sample**

| | | | Age groups^ | Area | | | | Sector | | Setting | | |
|---|---|---|---|---|---|---|---|---|---|---|---|---|
| **Activity** | **Number from Stage 1** | **Number from Stage 2** | **Children** | **Speech and language** | **Mental health and pragmatic language** | **Communication** | **Swallowing and mealtimes** | **Private** | **Public** | **Private practice** | **Community** | **Residential care** |
| Any | 3/11 | 2/3 | ✓ | ✓ | ✓ | ✓ | ✓ | ✓ | ✓ | ✓ | ✓ | ✓ |
| Land-based gross motor activities | 1/11 | 0/3 | | | | ✓ | | ✓ | | ✓ | | |
| Land-based fine motor activities | 0/11 | 0/3 | | | | | | | | | | |
| Water-based physical activities | 1/11 | 0/3 | | | | ✓ | | ✓ | | ✓ | | |
| Active transport | 0/11 | 1/3 | ✓ | | ✓ | | | ✓ | | ✓ | | |
| Powered transport | 0/11 | 0/3 | | | | | | | | | | |
| Gardening | 0/11 | 1/3 | ✓ | ✓ | | | | ✓ | | ✓ | | |
| Ecological conservation or restoration | 0/11 | 0/3 | | | | | | | | | | |
| Engaging with/ tending to animals | 2/11 | 1/3 | ✓ | ✓ | | ✓ | | ✓ | ✓ | ✓ | ✓ | |
| Wilderness activities | 0/11 | 1/3 | ✓ | | ✓ | | | ✓ | | ✓ | | |
| Nature play | 2/11 | 1/3 | ✓ | ✓ | | ✓ | | ✓ | ✓ | ✓ | ✓ | |
| Nature observation activities | 1/11 | 0/3 | | | | | ✓ | ✓ | | | | ✓ |
| Mindfulness/ meditation/ relaxation | 1/11 | 1/3 | ✓ | | ✓ | | ✓ | ✓ | | ✓ | | ✓ |
| Forest bathing | 0/11 | 0/3 | | | | | | | | | | |
| Social activities | 2/11 | 1/3 | ✓ | | ✓ | ✓ | ✓ | ✓ | | ✓ | | ✓ |
| No specific activity | 0/11 | 1/3 | ✓ | | ✓ | | | ✓ | | ✓ | | |
| Other activities | 0/11 | 0/3 | | | | | | | | | | |

Notes: Only headings relevant to our sample have been reported. ^the main age groups worked with were not asked about in Stage 1.

**Table S2.39: Locations in which outdoor nature-based activities were recommended by the speech pathologists in our sample**

| | | | Age groups^ | Area | | | | Sector | | Setting | | |
|---|---|---|---|---|---|---|---|---|---|---|---|---|
| **Location** | **Number from Stage 1** | **Number from Stage 2** | **Children** | **Speech and language** | **Mental health and pragmatic language** | **Communication** | **Swallowing and mealtimes** | **Private** | **Public** | **Private practice** | **Community** | **Residential care** |
| Private garden | 1/11 | 1/3 | ✓ | ✓ | | | ✓ | ✓ | | ✓ | | ✓ |
| Community/ school garden | 0/11 | 0/3 | | | | | | | | | | |
| Farm | 1/11 | 1/3 | ✓ | ✓ | | | | ✓ | ✓ | ✓ | ✓ | |
| Public park | 1/11 | 1/3 | ✓ | ✓ | | ✓ | | ✓ | | ✓ | | |
| National park | 0/11 | 1/3 | ✓ | | ✓ | | | ✓ | | ✓ | | |
| Forest | 0/11 | 1/3 | ✓ | | ✓ | | | ✓ | | ✓ | | |
| Oval/ sports field | 0/11 | 0/3 | | | | | | | | | | |
| On the lake | 0/11 | 0/3 | | | | | | | | | | |
| Near the lake | 0/11 | 1/3 | ✓ | | ✓ | | | ✓ | | ✓ | | |
| On the river | 0/11 | 0/3 | | | | | | | | | | |
| Near the river | 0/11 | 0/3 | | | | | | | | | | |
| On the ocean | 0/11 | 0/3 | | | | | | | | | | |
| Near the ocean | 0/11 | 1/3 | ✓ | | ✓ | | | ✓ | | ✓ | | |
| Other | 0/11 | 0/3 | | | | | | | | | | |

Notes: Only headings relevant to our sample have been reported. ^the main age groups worked with were not asked about in Stage 1.

**Table S2.40: Recommendations for indoor nature-based activities from the speech pathologists in our sample**

| | | | Age groups^ | Area | | Sector | | Setting | | |
|---|---|---|---|---|---|---|---|---|---|---|
| **Activity** | **Number from Stage 1** | **Number from Stage 2** | **Children** | **Speech and language** | **Mental health and pragmatic language** | **Private** | **Public** | **Private practice** | **Home and community based service** | **Community** |
| Any | 2/5 | 1/2 | ✓ | ✓ | ✓ | ✓ | ✓ | ✓ | ✓ | ✓ |
| Listening to natural sounds | 0/5 | 0/2 | | | | | | | | |
| Having indoor pot plants | 0/5 | 0/2 | | | | | | | | |
| Pictures of natural scenes | 1/5 | 0/2 | ✓ | ✓ | | | ✓ | | | ✓ |
| Natural scents | 0/5 | 0/2 | | | | | | | | |
| Travelling in a car or on public transport through natural environments | 0/5 | 0/2 | | | | | | | | |
| Views of nature outside the window | 2/5 | 0/2 | | ✓ | | ✓ | ✓ | | ✓ | ✓ |
| Open windows for fresh air and the smells and sounds of nature | 1/5 | 1/2 | ✓ | ✓ | ✓ | ✓ | | ✓ | ✓ | |
| Nature-based virtual reality | 0/5 | 0/2 | | | | | | | | |
| Other | 0/5 | 0/2 | | | | | | | | |

Notes: Only headings relevant to our sample have been reported. ^the main age groups worked with were not asked about in Stage 1.

### Direct engagement

**Table S2.41: Direct engagement in outdoor nature-based activities by social workers in our sample with their patients/ clients**

| | | | Age groups^ | Area | | | Sector | | Setting | | | |
|---|---|---|---|---|---|---|---|---|---|---|---|---|
| **Activity** | **Number from Stage 1** | **Number from Stage 2** | **Children** | **Speech and language** | **Mental health and pragmatic language** | **Swallowing and mealtimes** | **Private** | **Public** | **Private practice** | **Home and community based service** | **Community** | **Residential care** |
| Any | 3/11 | 1/2 | ✓ | ✓ | ✓ | ✓ | ✓ | ✓ | ✓ | ✓ | ✓ | ✓ |
| Land-based gross motor activities | 1/11 | 0/2 | ✓ | ✓ | | | | ✓ | | | ✓ | |
| Land-based fine motor activities | 0/11 | 0/2 | | | | | | | | | | |
| Water-based physical activities | 0/11 | 0/2 | | | | | | | | | | |
| Active transport | 0/11 | 0/2 | | | | | | | | | | |
| Powered transport | 0/11 | 0/2 | | | | | | | | | | |
| Gardening | 0/11 | 0/2 | | | | | | | | | | |
| Ecological conservation or restoration | 0/11 | 0/2 | | | | | | | | | | |
| Engaging with/ tending to animals | 1/11 | 0/2 | ✓ | ✓ | | | | ✓ | | | ✓ | |
| Wilderness activities | 0/11 | 1/2 | ✓ | | ✓ | | ✓ | | ✓ | | | |
| Nature play | 1/11 | 0/2 | ✓ | ✓ | | | | ✓ | | | ✓ | |
| Nature observation activities | 1/11 | 0/2 | ✓ | ✓ | | | | ✓ | | | ✓ | |
| Mindfulness/ meditation/ relaxation | 0/11 | 1/2 | ✓ | | ✓ | | ✓ | | ✓ | | | |
| Forest bathing | 0/11 | 0/2 | | | | | | | | | | |
| Social activities | 1/11 | 1/2 | ✓ | | ✓ | ✓ | ✓ | | ✓ | | | ✓ |
| No specific activity | 2/11 | 1/2 | ✓ | ✓ | ✓ | ✓ | ✓ | ✓ | ✓ | | ✓ | ✓ |
| Other activities: speech/ language/ communication therapy in parks | 1/11 | 0/2 | ✓ | ✓ | | | ✓ | | | ✓ | | |

Notes: Only headings relevant to our sample have been reported. ^the main age groups worked with were not asked about in Stage 1.

**Table S2.42: Locations in which the occupational therapists in our sample engaged directly with their patients/ clients in outdoor nature-based activities**

| | | | Age groups^ | Area | | | Sector | | Setting | | | |
|---|---|---|---|---|---|---|---|---|---|---|---|---|
| Location | Number from Stage 1 | Number from Stage 2 | Children | Speech and language | Mental health and pragmatic language | Swallowing and mealtimes | Private | Public | Private practice | Home and community based service | Community | Residential care |
| Private garden | 2/11 | 0/2 | | ✓ | | ✓ | ✓ | ✓ | | ✓ | ✓ | |
| Community/ school garden | 2/11 | 0/2 | | ✓ | | | ✓ | ✓ | | ✓ | ✓ | |
| Farm | 0/11 | 0/2 | | | | | | | | | | |
| Public park | 2/11 | 1/2 | ✓ | ✓ | ✓ | ✓ | ✓ | | ✓ | ✓ | | ✓ |
| National park | 0/11 | 1/2 | ✓ | | ✓ | | ✓ | | ✓ | | | |
| Forest | 0/11 | 1/2 | ✓ | | ✓ | | ✓ | | ✓ | | | |
| Oval/ sports field | 0/11 | 0/2 | | | | | | | | | | |
| On the lake | 0/11 | 0/2 | | | | | | | | | | |
| Near the lake | 1/11 | 1/2 | ✓ | ✓ | ✓ | | ✓ | | ✓ | ✓ | | |
| On the river | 0/11 | 0/2 | | | | | | | | | | |
| Near the river | 0/11 | 0/2 | | | | | | | | | | |
| On the ocean | 0/11 | 0/2 | | | | | | | | | | |
| Near the ocean | 0/11 | 0/2 | | | | | | | | | | |
| Other | 0/11 | 0/2 | | | | | | | | | | |

Notes: Only headings relevant to our sample have been reported. ^the main age groups worked with were not asked about in Stage 1.

**Table S2.43: Direct engagement in indoor nature-based activities by social workers in our sample with their patients/ clients**

| | | | Age groups^ | Area | | Sector | | Setting | | |
|---|---|---|---|---|---|---|---|---|---|---|
| **Activity** | **Number from Stage 1** | **Number from Stage 2** | **Children** | **Speech and language** | **Mental health and pragmatic language** | **Private** | **Public** | **Private practice** | **Home and community based service** | **Community** |
| Any | 2/5 | 1/2 | ✓ | ✓ | ✓ | ✓ | ✓ | ✓ | ✓ | ✓ |
| Listening to natural sounds | 0/5 | 0/2 | | | | | | | | |
| Having indoor pot plants | 0/5 | 0/2 | | | | | | | | |
| Pictures of natural scenes | 1/5 | 0/2 | | ✓ | | | ✓ | | | ✓ |
| Natural scents | 0/5 | 0/2 | | | | | | | | |
| Travelling in a car or on public transport through natural environments | 0/5 | 0/2 | | | | | | | | |
| Views of nature outside the window | 2/5 | 0/2 | | ✓ | | ✓ | ✓ | | ✓ | ✓ |
| Open windows for fresh air and the smells and sounds of nature | 1/5 | 1/2 | ✓ | ✓ | ✓ | ✓ | | ✓ | ✓ | |
| Nature-based virtual reality | 0/5 | 0/2 | | | | | | | | |
| Other | 0/5 | 0/2 | | | | | | | | |

Notes: Only headings relevant to our sample have been reported. ^the main age groups worked with were not asked about in Stage 1.

### Enhancing outdoor natural environments

Only one social worker in our sample engaged in advocacy for the addition of or improvement to outdoor natural spaces. They reported advocating for the organisation to move their premises to a location with more outdoor space. This social worker worked in the speech and language area, in the public sector, in a community setting.

No social workers in our sample engaged in the provision of the addition of or improvement to outdoor natural spaces.
